# Daily Forecasting of New Cases for Regional Epidemics of Coronavirus Disease 2019 with Bayesian Uncertainty Quantification

**DOI:** 10.1101/2020.07.20.20151506

**Authors:** Yen Ting Lin, Jacob Neumann, Ely F. Miller, Richard G. Posner, Abhishek Mallela, Cosmin Safta, Jaideep Ray, Gautam Thakur, Supriya Chinthavali, William S. Hlavacek

**Affiliations:** Los Alamos National Laboratory, Los Alamos, New Mexico, USA; Northern Arizona University, Flagstaff, Arizona, USA; University of California, Davis, California, USA; Sandia National Laboratories, Livermore, California, USA; Oak Ridge National Laboratory, Oak Ridge, Tennessee, USA

**Keywords:** Mathematical Model, Statistics, Uncertainty, Epidemics, Coronavirus Disease 2019 (COVID-19), Severe Acute Respiratory Syndrome Coronavirus 2 (SARS-CoV-2)

## Abstract

To increase situational awareness and support evidence-based policy-making, we formulated a mathematical model for COVID-19 transmission within a regional population. This compartmental model accounts for quarantine, self-isolation, social distancing, a non-exponentially distributed incubation period, asymptomatic individuals, and mild and severe forms of symptomatic disease. Using Bayesian inference, we have been calibrating region-specific models daily for consistency with new reports of confirmed cases from the 15 most populous metropolitan statistical areas in the United States and quantifying uncertainty in parameter estimates and predictions of future case reports. This online learning approach allows for early identification of new trends despite considerable variability in case reporting.

**Article Summary Line:** We report models for regional COVID-19 epidemics and use of Bayesian inference to quantify uncertainty in daily predictions of expected reporting of new cases, enabling identification of new trends in surveillance data.

## Text

Coronavirus disease 2019 (COVID-19), caused by severe acute respiratory syndrome coronavirus 2 (SARS-CoV-2) *(1)*, was detected in the United States (US) in January 2020 *(2)*. In February, COVID-19-caused deaths were detected *(3)*. Thereafter, surveillance testing expanded nationwide *(4)*. These and other efforts revealed community spread across the US and exponential growth of new COVID-19 cases throughout most of March with a doubling time of 2 to 3 d *(5)*, similar to that of the initial outbreak in China *(6)*. This situation led to government mandates prohibiting, for example, public gatherings, as well as broad adoption of social-distancing practices, such as working-from-home, curtailing of travel, and face mask-wearing *(7)*. Although the US became a hot spot of the COVID-19 pandemic, detection of new cases peaked in late-April and steadily declined until mid-June *(4)*, suggesting that mandates and social distancing were effective at slowing COVID-19 transmission. Attempts to quantify the impacts of these measures suggest substantial reduction of disease burden *(8, 9)*.

In mid-June and mid-September, the number of new daily cases in the US began increasing a second and third time *(4)*. It is imperative that we effectively monitor ongoing COVID-19 transmission, so that dangerous upticks in cases can be responded to as quickly as possible.

To contribute to situational awareness of COVID-19 transmission dynamics, we developed a mathematical model for the regional COVID-19 epidemic in each of the 15 most populous US metropolitan statistical areas (MSAs) *(10)*. Each model is composed of ordinary differential equations (ODEs), which characterize the dynamics of various populations, including mixing and protected-by-social-distancing subpopulations.

In an ongoing online-learning effort, we have been calibrating our models for regional COVID-19 epidemics on a daily basis for consistency with historical case reports. We have also been applying Bayesian methods to quantify uncertainties in predicted detection of new cases. In the face of variability in case detection, this approach allows for identification of new epidemic trends, enabling an evidence-based response by policy makers.

## Methods

### Data Used in Online Learning

The COVID-19 surveillance data used to parameterize models—reports of new confirmed cases—are obtained daily (at variable times of day) from the GitHub repository maintained by *The New York Times* newspaper *(11)*. We aggregate county-level case counts to obtain case counts for each of the 15 most populous US MSAs. These MSAs encompass the following cities: New York City; Los Angeles; Chicago; Dallas; Houston; Washington, DC; Miami; Philadelphia; Atlanta; Phoenix; Boston; San Francisco; Riverside, CA; Detroit; and Seattle. The political entities comprising each MSA, which are almost always counties^1^, are those delineated by the federal government *(10)*.

### Models for COVID-19 Transmission and Model Parameters

In ongoing work, each day, for the regional COVID-19 epidemic in each of the 15 MSAs of interest, we parameterize a compartmental model for consistency with all daily reports of new confirmed cases that are available at the time^2^.

Each MSA-specific model accounts for 25 populations (Figure 1 and Appendix Figure 1). Infectious individuals are taken to be exposed and incubating virus (e.g., presymptomatic), asymptomatic while clearing virus, or symptomatic. The parameters *ρ*_*E*_ and *ρ*_*A*_ characterize the relative infectiousness of exposed and asymptomatic individuals compared to symptomatic individuals. Infected individuals move to quarantine with rate constant *k*_*Q*_. Symptomatic individuals with mild disease quarantine themselves with rate constant *j*_*Q*_. Social distancing is modeled by allowing susceptible and infectious individuals to transition between mixing and protected populations. The size of the protected population is determined by two parameters: *λ*_*i*_, a rate constant, and *p*_*i*_, a steady-state population setpoint, where index *i* refers to the current social-distancing period. The model allows for *n* distinct social-distancing periods beyond an initial period of social distancing. Individuals in the protected population are less likely to be infected and less likely to transmit disease by a factor *m*_*b*_ < 1. Within the mixing population, disease is transmitted with rate constant *β*. The model reproduces a non-exponentially distributed incubation period by dividing the incubation period into five sequential stages of equal mean duration, 1/*k*_*L*_. Individuals in the first incubation period are taken to be non-infectious and non-detectable as infected. A fraction of exposed individuals exiting the incubation period, *f*_*A*_, never develop symptoms. The remaining individuals develop mild disease. A fraction of these individuals, *f*_*H*_, progress to severe disease/hospitalization; the others recover. A fraction of individuals with severe disease, *f*_*R*_, recover; the others die. Hospitalized individuals (or at home with severe disease) are taken to be quarantined. Individuals leave the asymptomatic state with rate constant *c*_*A*_, leave the mild disease state with rate constant *c*_*I*_, and leave the severe disease/hospitalized state with rate constant *c*_*H*_.

**Figure 1.**
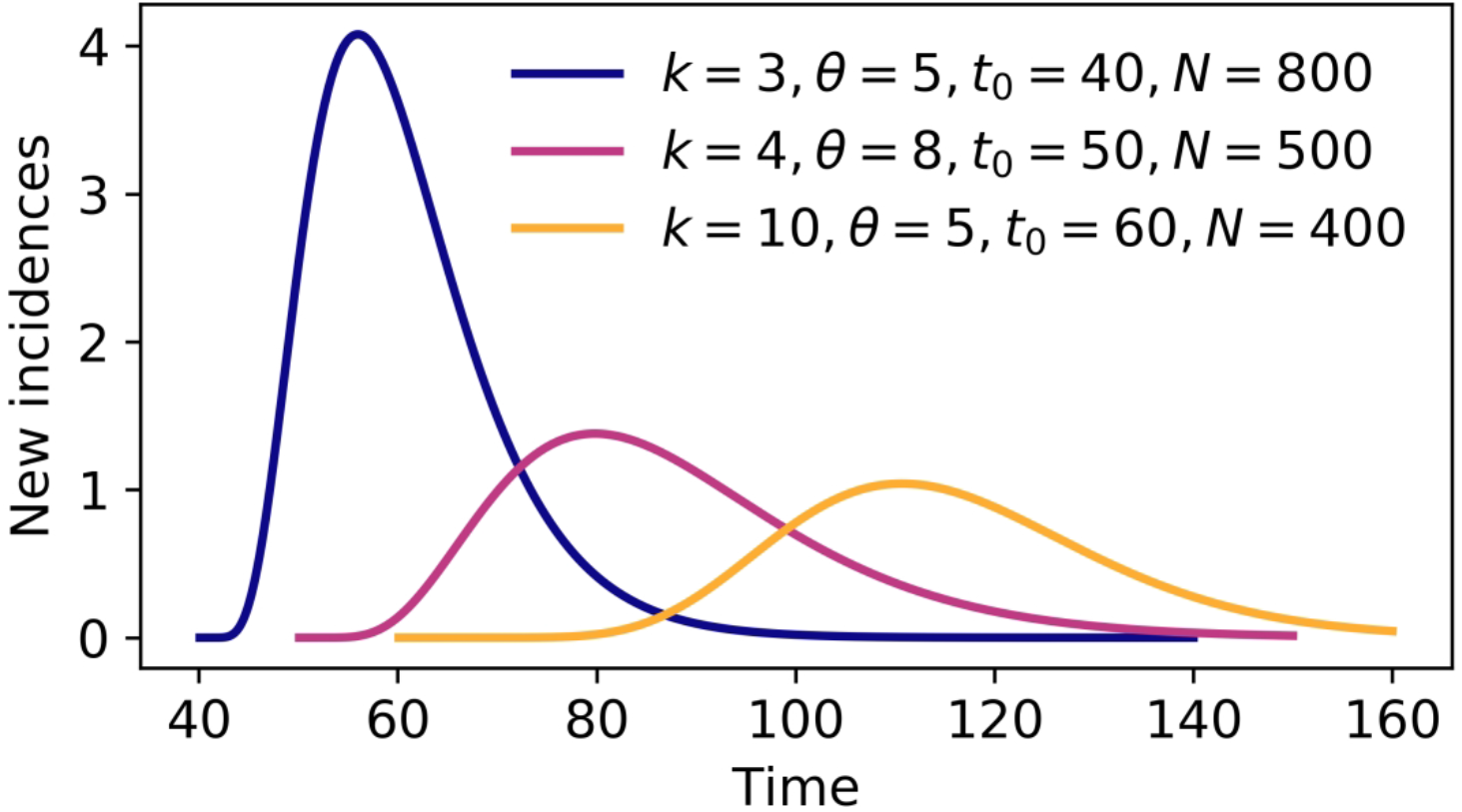
Illustration of the populations and processes considered in a mechanistic compartmental model for the dynamics of COVID-19 transmission. The model accounts for susceptible individuals (*S*), exposed individuals without symptoms in the incubation phase of disease (*E*), asymptomatic individuals in the immune clearance phase of disease (*A*), mildly ill symptomatic individuals (*I*), severely ill individuals in hospital or at home (*H*), recovered individuals (*R*), and deceased individuals (*D*). The model also accounts for social distancing, which establishes mixing and protected subpopulations denoted by M and P subscripts, respectively; quarantine driven by testing and contact tracing, which establishes quarantined subpopulations denoted by a Q subscript; and self-isolation spurred by symptom awareness. Individuals who are self-isolating because of symptoms are taken to be members of the *I*_*Q*_ population. The incubation period is divided into 5 stages (*E*_1_ through *E*_5_), which allows the model to reproduce an empirically determined (non-exponential) Erlang distribution of waiting times for the onset of symptoms after infection *(12)*. The exposed population (consisting of individuals incubating virus) includes both presymptomatic and asymptomatic individuals. The *A*-populations consist of true asymptomatic individuals in the immune clearance phase. The gray background indicates the populations that contribute to disease transmission. An auxiliary measurement model (Equations (23) and (24) in the Appendix) accounts for imperfect detection and reporting of new cases. Only symptomatic cases are assumed to be detectable in surveillance testing.

The model consists of 25 ordinary differential equations (ODEs), defined by Equations (1)–(22) in the Appendix. Each state variable of the model represents the size of a population. In addition to the 25 ODEs, we consider an auxiliary 1-parameter measurement model that relates state variables to expected case reporting (see Equations (23) and (24) in the Appendix) and a negative binomial model for variability in new case detection (see Equations (25)–(27) in the Appendix). The model is formulated so as to allow consideration of multiple periods of social distancing with distinct setpoints for the protected population size. In the model, there is always an initial period of social distancing. The number of additional social-distancing periods is indicated by *n*. Here, we only consider two cases: *n* = 0 and *n* = 1. The best value of *n* is determined using a model selection procedure described in the Appendix.

For *n* = 0, the compartmental model and auxiliary measurement model have a total of 20 parameters. We take six of these parameters to have adjustable values (Table 1) and 14 to have fixed values (Tables 2 and 3). In the Appendix, we describe each parameter and explain the fixed parameter settings, which are based on information in Refs. *(12)–(20)* and assumptions. The adjustable model parameters are *t*_0_, the start time of the local epidemic; *σ* > *t*_0_, the time at which social distancing begins; *p*_0_, which establishes a setpoint for the quasi-stationary fraction of the total population practicing social distancing; *λ*_0_, which characterizes the rate of movement between the mixing and protected subpopulations and establishes a timescale for population-level adoption of social-distancing practices; and *β*, which characterizes the rate of disease transmission in the absence of social distancing. The measurement-model parameter, *f*_*D*_, represents the time-averaged fraction of new cases detected. Inference of adjustable parameter values is based on a negative binomial likelihood function (Equation (27) in the Appendix). The dispersal parameter *r* of the likelihood is taken to be adjustable; its value is jointly inferred with those of *t*_0_, *σ, p*_0_, *λ*_0_, *β*, and *f*_*D*_.

**Table 1.** Inferred values of the adjustable parameters of the compartmental model (*t*_0_, *σ, p*_0_, *λ*_0_, and *β*), the auxiliary measurement model (*f*_*D*_), and the associated statistical model for noise in case detection and reporting (*r*).

| Parameter | Estimate* (Units) | Comment |
| --- | --- | --- |
| $t_0$ | 33 (d) | Start of COVID-19 transmission |
| $\sigma$ | 33 (d) | Start of social distancing |
| $p_0$ | 0.87 | Social-distancing setpoint |
| $\lambda_0$ | 0.10 (/d) | Social-distancing rate parameter |
| $\beta$ | 2.0 (/d) | Disease-transmission rate parameter |
| $f_D$ | 0.12 | Fraction of active cases reported |
| $r$ | 12 | Dispersal parameter of NB($r, p$)** |

\*All estimates are region-specific and inference-time-dependent. Inferences are performed daily. Here, we report the maximum a posteriori (MAP) estimates inferred for the New York City MSA using all confirmed COVID-19 case-count data available in the GitHub repository maintained by *The New York Times* newspaper (11) for the period starting on 21-January-2020 and ending on 21-June-2020 (inclusive dates). Time $t = 0$ corresponds to 0000 hours on 21-January-2020. \*\*The probability parameter of NB( $r, p$ ) is constrained, i.e., its reporting-time-dependent value is determined by a formula, which is given by Equation (26) in the Appendix.

**Table 2.** Estimates for the fixed parameters of the compartmental model.

| Parameter | Estimate (Units) | Source |
| --- | --- | --- |
| $S_0$ | 19,216,182* | (13) |
| $I_0$ | 1 | Assumption |
| $n$ | 0** | Assumption |
| $m_b$ | 0.1 | Assumption |
| $\rho_E$ | 1.1 | Arons et al. (14) |
| $\rho_A$ | 0.9 | Nguyen et al. (15) |
| $k_L$ | 0.94 (/d) | Lauer et al. (12) |
| $k_Q$ | 0.0038 (/d) | Assumption |
| $j_Q$ | 0.4 (/d) | Assumption |
| $f_A$ | 0.44 | (16, 17) |
| $f_H$ | 0.054 | Perez-Saez et al. (18) |
| $f_R$ | 0.79 | Richardson et al. (19) |
| $c_A$ | 0.26 (/d) | Sakurai et al. (17) |
| $c_I$ | 0.12 (/d) | Wölfel et al. (20) |
| $c_H$ | 0.17 (/d) | Richardson et al. (19) |

\*All estimates listed in this table are taken to apply to all regions of interest except for $n$ , the number of distinct social-distancing periods that follow an initial social-distancing period, and $S_0$ , the region-specific initial number of susceptible individuals. The value given here for $S_0$ is the US Census Bureau-estimated total population of the New York City metropolitan statistical area. \*\*We assume $n = 0$ unless stated otherwise.

**Table 3.**
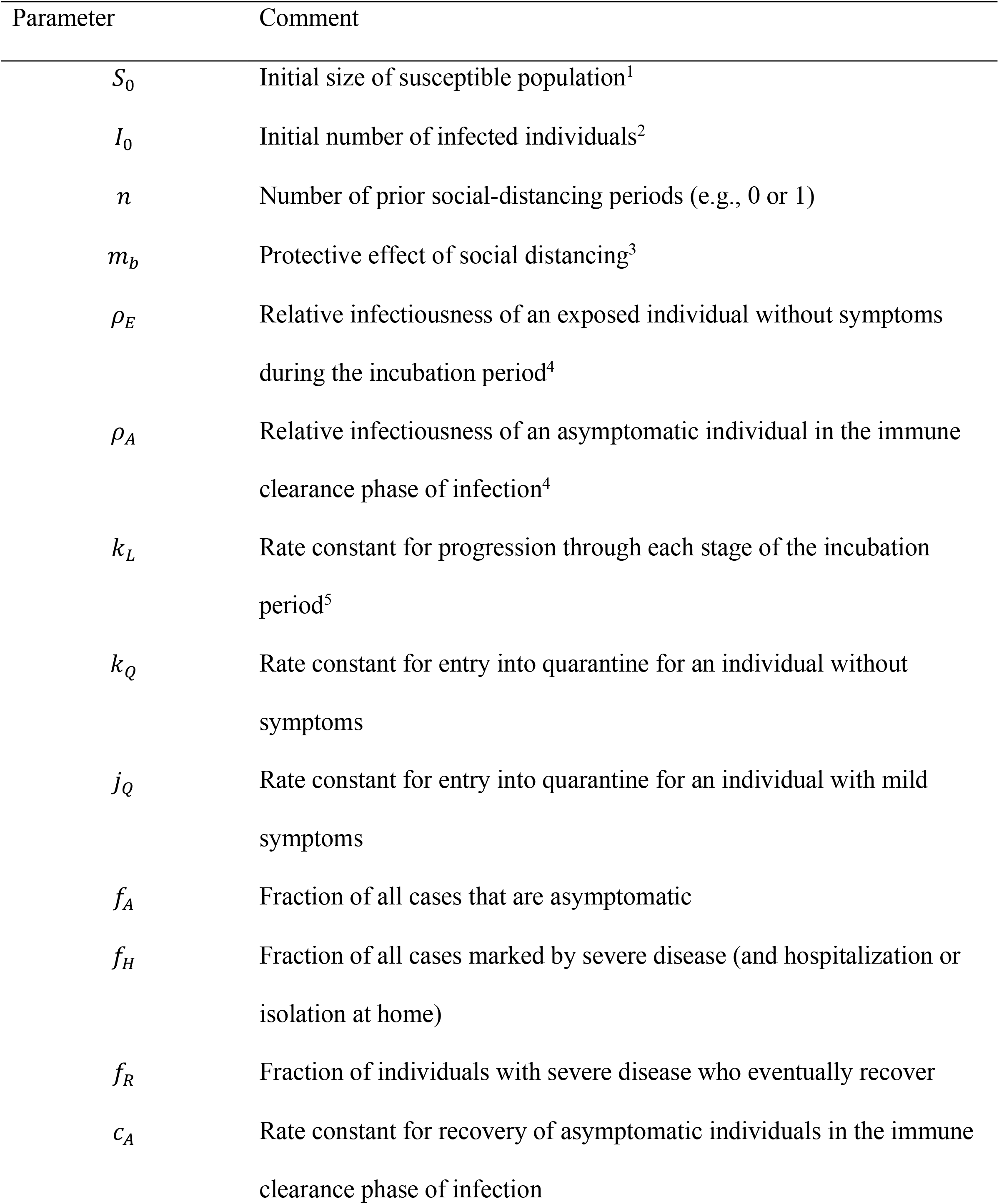
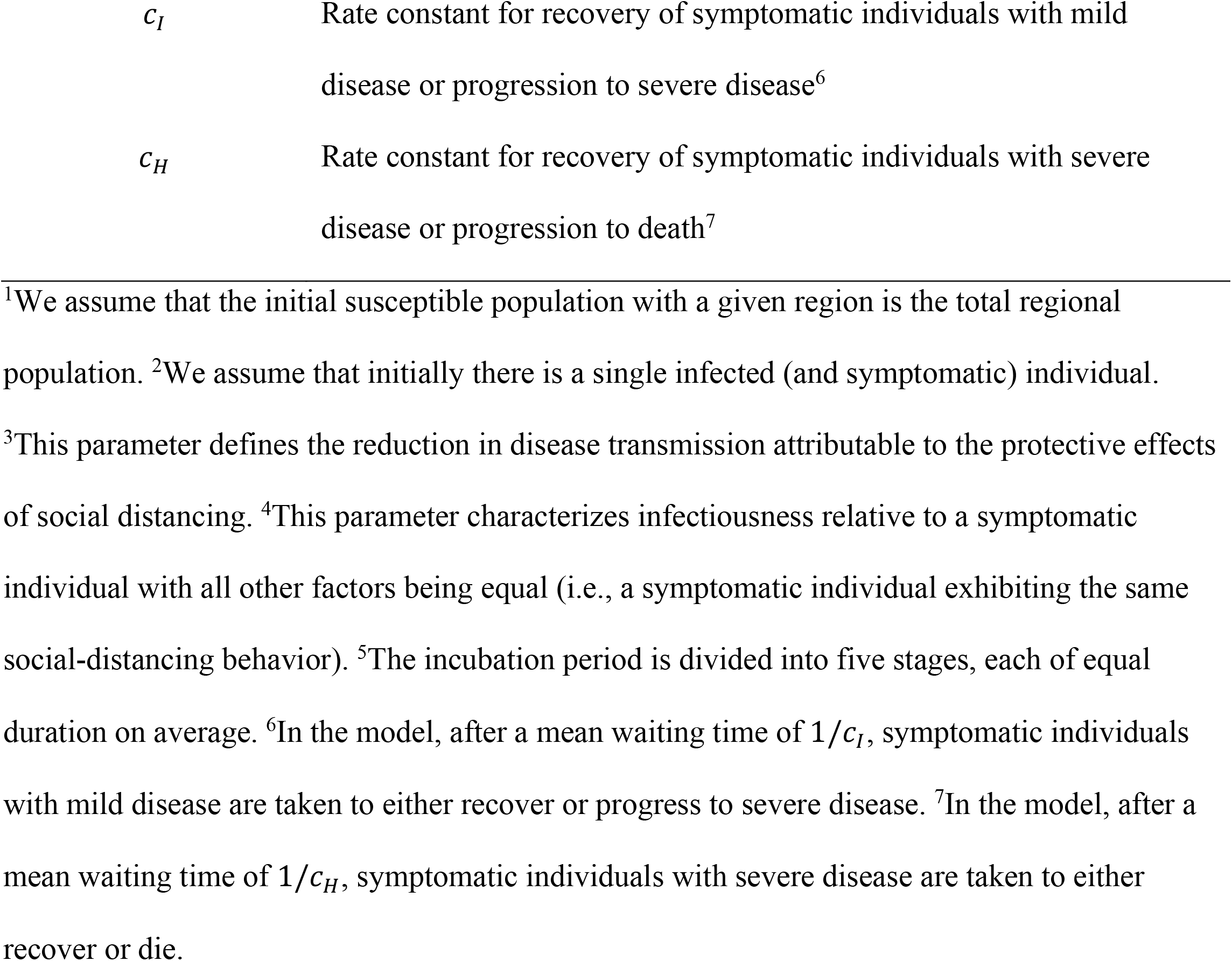
Description of the fixed parameters of the compartmental model.

| Parameter | Comment |
| --- | --- |
| $S_0$ | Initial size of susceptible population<sup>1</sup> |
| $I_0$ | Initial number of infected individuals<sup>2</sup> |
| $n$ | Number of prior social-distancing periods (e.g., 0 or 1) |
| $m_b$ | Protective effect of social distancing<sup>3</sup> |
| $\rho_E$ | Relative infectiousness of an exposed individual without symptoms during the incubation period<sup>4</sup> |
| $\rho_A$ | Relative infectiousness of an asymptomatic individual in the immune clearance phase of infection<sup>4</sup> |
| $k_L$ | Rate constant for progression through each stage of the incubation period<sup>5</sup> |
| $k_Q$ | Rate constant for entry into quarantine for an individual without symptoms |
| $j_Q$ | Rate constant for entry into quarantine for an individual with mild symptoms |
| $f_A$ | Fraction of all cases that are asymptomatic |
| $f_H$ | Fraction of all cases marked by severe disease (and hospitalization or isolation at home) |
| $f_R$ | Fraction of individuals with severe disease who eventually recover |
| $c_A$ | Rate constant for recovery of asymptomatic individuals in the immune clearance phase of infection |
| $c_I$ | Rate constant for recovery of symptomatic individuals with mild disease or progression to severe disease<sup>6</sup> |
| $c_H$ | Rate constant for recovery of symptomatic individuals with severe disease or progression to death<sup>7</sup> |

<sup>1</sup>We assume that the initial susceptible population with a given region is the total regional population. <sup>2</sup>We assume that initially there is a single infected (and symptomatic) individual. <sup>3</sup>This parameter defines the reduction in disease transmission attributable to the protective effects of social distancing. <sup>4</sup>This parameter characterizes infectiousness relative to a symptomatic individual with all other factors being equal (i.e., a symptomatic individual exhibiting the same social-distancing behavior). <sup>5</sup>The incubation period is divided into five stages, each of equal duration on average. <sup>6</sup>In the model, after a mean waiting time of $1/c_I$ , symptomatic individuals with mild disease are taken to either recover or progress to severe disease. <sup>7</sup>In the model, after a mean waiting time of $1/c_H$ , symptomatic individuals with severe disease are taken to either recover or die.

For *n* > 0 distinct social-distancing periods after an initial social-distancing period, the compartmental model has three additional adjustable parameters for each additional period of social distancing. For one additional period of social distancing (*n* = 1), the additional adjustable parameters are *τ*_1_ > *σ*, the onset time of second-phase social-distancing; *p*_1_, the second-phase quasi-stationary setpoint parameter; and *λ*_1_, which determines the timescale for transition from first-to second-phase social-distancing behavior. A second social-distancing period is considered by replacing *p*_0_ with *p*_1_ and *λ*_0_ with *λ*_1_ at time *t* = *τ*_1_. If adherence to effective social-distancing practices begins to relax at time *t* = *τ*_1_, then *p*_1_ < *p*_0_.

### Statistical Model for Noisy Case Reporting

The compartmental model is deterministic. We interpret the model to predict the expected number of new confirmed COVID-19 cases reported daily. In other words, we assume that the number of new cases reported over a 1-d period is a random variable and its expected value follows a deterministic trajectory. We further assume that day-to-day fluctuations in the random variable are independent and characterized by a negative binomial distribution, which we will denote as N*B*(*r, p*). We use N*B*(*r, p*) to statistically model noise in reporting (and case detection) because its support—the non-negative integers—is natural for populations and its shape is flexible enough to recapitulate an array of unimodal distributions. With these assumptions, we obtain a likelihood function (Equation (27) in the Appendix) taking the form of a product of probability mass functions of N*B*(*r, p*). Formulation of a likelihood is a prerequisite for standard Bayesian inference^3^.

### Online Learning of Model Parameter Values through Bayesian Inference

We use Bayesian inference to learn adjustable model parameter values. Inferences are performed daily for each MSA of interest. In each inference, we assume a uniform prior and use an adaptive Markov chain Monte Carlo (MCMC) algorithm *(21)* to generate samples of the posterior distribution for the adjustable parameters. A full description of our inference procedure is provided in the Appendix.

The maximum a posteriori (MAP) estimate of a parameter value is the value of the parameter corresponding to the mode of its marginal posterior, where probability mass is highest. MAP estimates are maximum likelihood estimates because we assume a uniform prior.

### Forecasting with Quantification of Prediction Uncertainty: Bayesian Predictive Inference

In addition to inferring parameter values, we quantify uncertainty in predicted trajectories of daily case reports. A predictive inference of the expected number of new cases detected on a given day is derived from a model by parameterizing it using a randomly chosen parameter posterior sample generated in MCMC sampling. The number of cases detected is then predicted by adding a noise term, drawn from N*B*(*r, p*), where *r* is set at the randomly sampled value and *p* is set using Equation (26) in the Appendix.

For any given (1-day) surveillance period and specified settings for parameter values, a prediction of the compartmental model is obtained by using LSODA *(22)* to numerically integrate Equations (1)–(17) and (23) in the Appendix; the initial condition is defined by the inferred value of *t*_*_ (Table 1) and the fixed settings for *S*_*_ and *I*_*_ (Tables 2 and 3). A prediction of the actual number of new cases detected is obtained by entering the predicted expected number of new cases into Equation (29) of the Appendix.

The 95% credible interval for the predicted number of new case reports on a given day is the central part of the marginal predictive posterior capturing 95% of the probability mass. This region is bounded above by the 97.5 percentile and below by the 2.5 percentile

## Results

The objective of our ongoing study is to detect significant new trends in new COVID-19 cases as early as possible by 1) systematically and regularly updating mathematical models capturing historical trends in regional COVID-19 epidemics through Bayesian inference and 2) making forecasts with rigorously quantified uncertainties through Bayesian predictive inference.

An important aspect of how we are analyzing COVID-19 data is our focus on the populations of US cities and their surrounding areas (i.e., MSAs) vs. the regional populations within other political boundaries, such as those of the US States. The boundaries of MSAs are defined on the basis of social and economic interactions *(10)*, which suggests that the population of an MSA is likely to be more uniformly affected by the COVID-19 pandemic than, for example, the population of a State. In accordance with this expectation, daily reports of new COVID-19 cases for New York City (Figure 2, panel A) are more temporally correlated than for the US State of New York (Figure 2, panel B), New Jersey (Figure 2, panel C), or Pennsylvania (Figure 2, panel D). New York, New Jersey, and Pennsylvania are the three States encompassing the New York City MSA.

**Figure 2.**
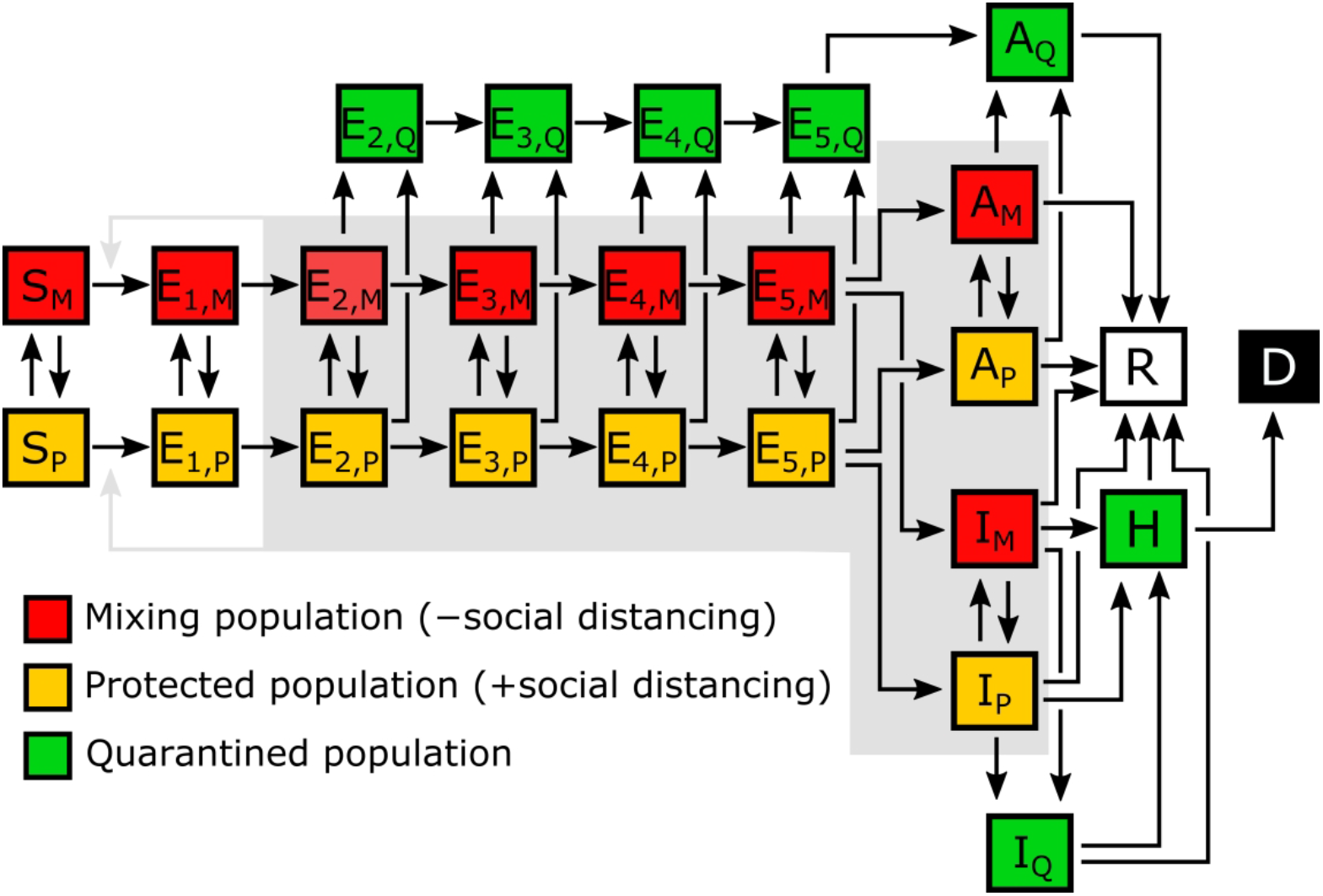
Temporal correlations in surveillance data. Shown here are time-series of fractional (i.e., normalized) case counts. We define the fractional case count for a county on a given date to be the reported number of cases on that date divided by the total reported number of cases in the county over the entire period of interest. The panels in this figure show fractional case counts for **(A)** the 23 counties comprising the New York City metropolitan statistical area (MSA), **(B)** the 62 counties comprising the State of New York, **(C)** the 21 counties comprising the State of New Jersey, and **(D)** the 67 counties comprising the State of Pennsylvania. Within each plot, a different color is used for the data points from each distinct county. As can be seen, time-series for the counties of the New York City MSA are more temporally correlated than for the State-level time-series. Daily case counts for New Jersey are similar to those for New York City because the two populations overlap considerably: ∼74% of New Jersey’s population is part of the New York City MSA and ∼32% of the population of the New York City MSA is part of the State of New Jersey. Time-averaged Fano factors for the four regions are as follows: 0.0026 (New York City), 0.021 (State of New York), 1.2 (State of New Jersey), and 0.028 (State of Pennsylvania). A smaller Fano factor indicates less county-to-county variability.

For each of the 15 most populous MSAs in the US, we parameterized a compartmental model using MSA-specific surveillance data, namely, aggregated county-level reports indicating the number of new confirmed COVID-19 cases within a given MSA each day. Bayesian parameterization and forecasting with uncertainty quantification (UQ)—predictive inference— were performed daily for each of the 15 MSAs.

Results of Bayesian predictive inference are exemplified in Figure 3. Predictions are obtained in the form of a predictive posterior distribution. In Figure 3, the entire shaded region indicates the 95% credible interval of the predictive posterior as a function of time. In other words, this band indicates where 95% of predictions of daily case reports fall at the times indicated. Predictions vary because of the uncertainties in adjustable model parameter estimates, which are characterized quantitatively through Bayesian inference. The colors within the shaded band indicate other credible intervals (10%, 20%, etc.) and also the median of all predictions as a function of time. The dataset used in inference is the complete time series of available daily new case counts for the region of interest.

**Figure 3.**
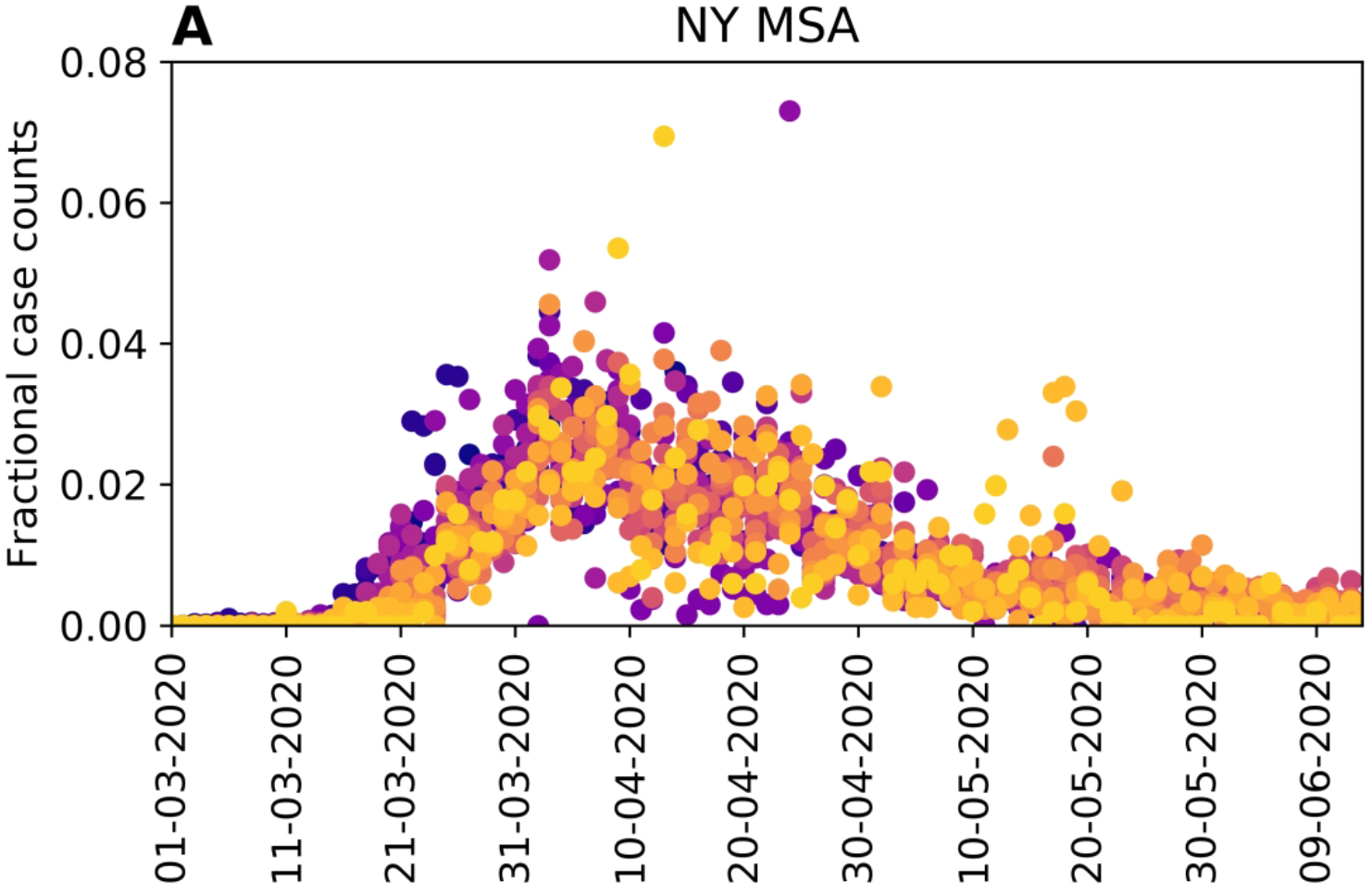
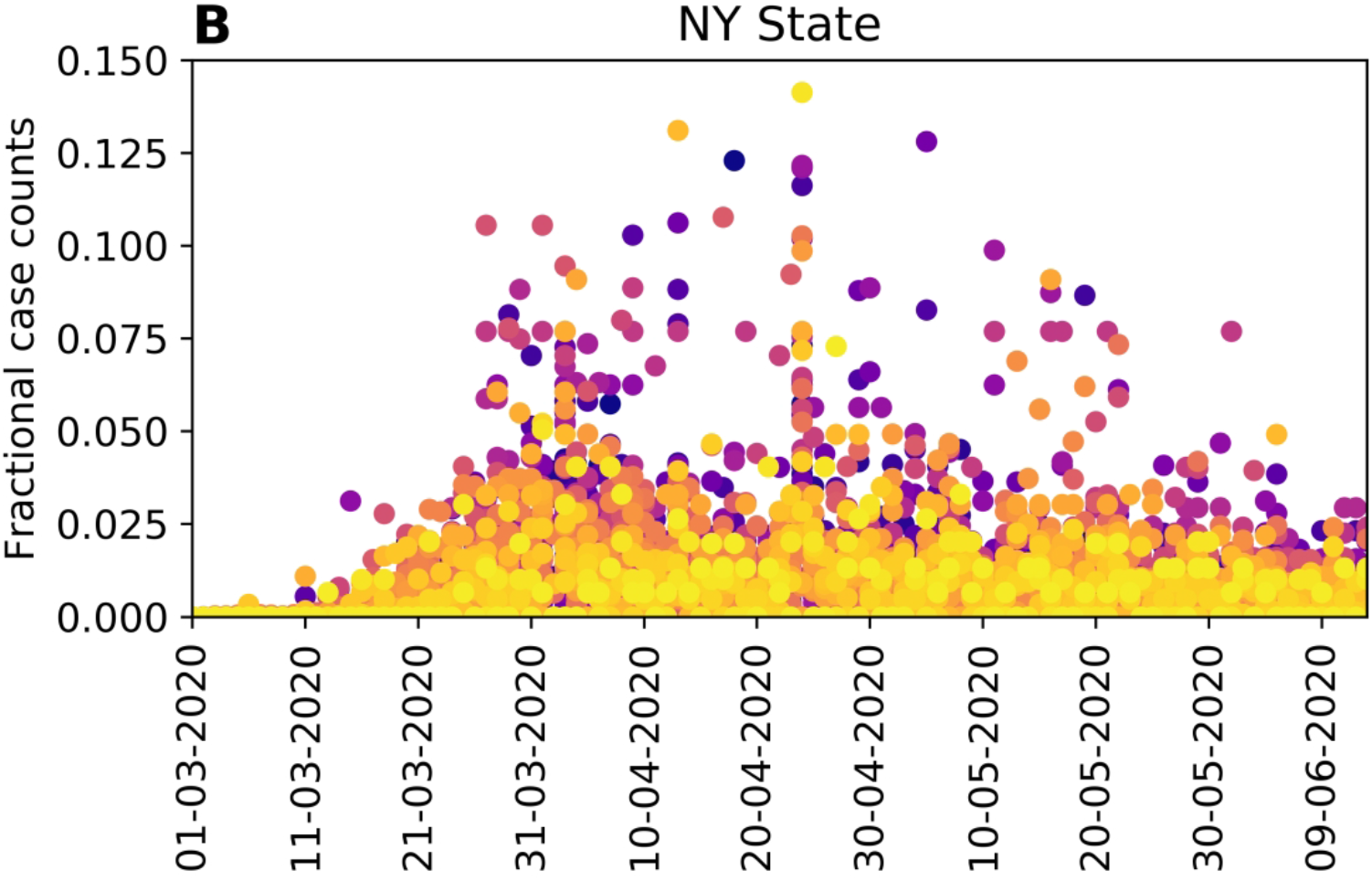
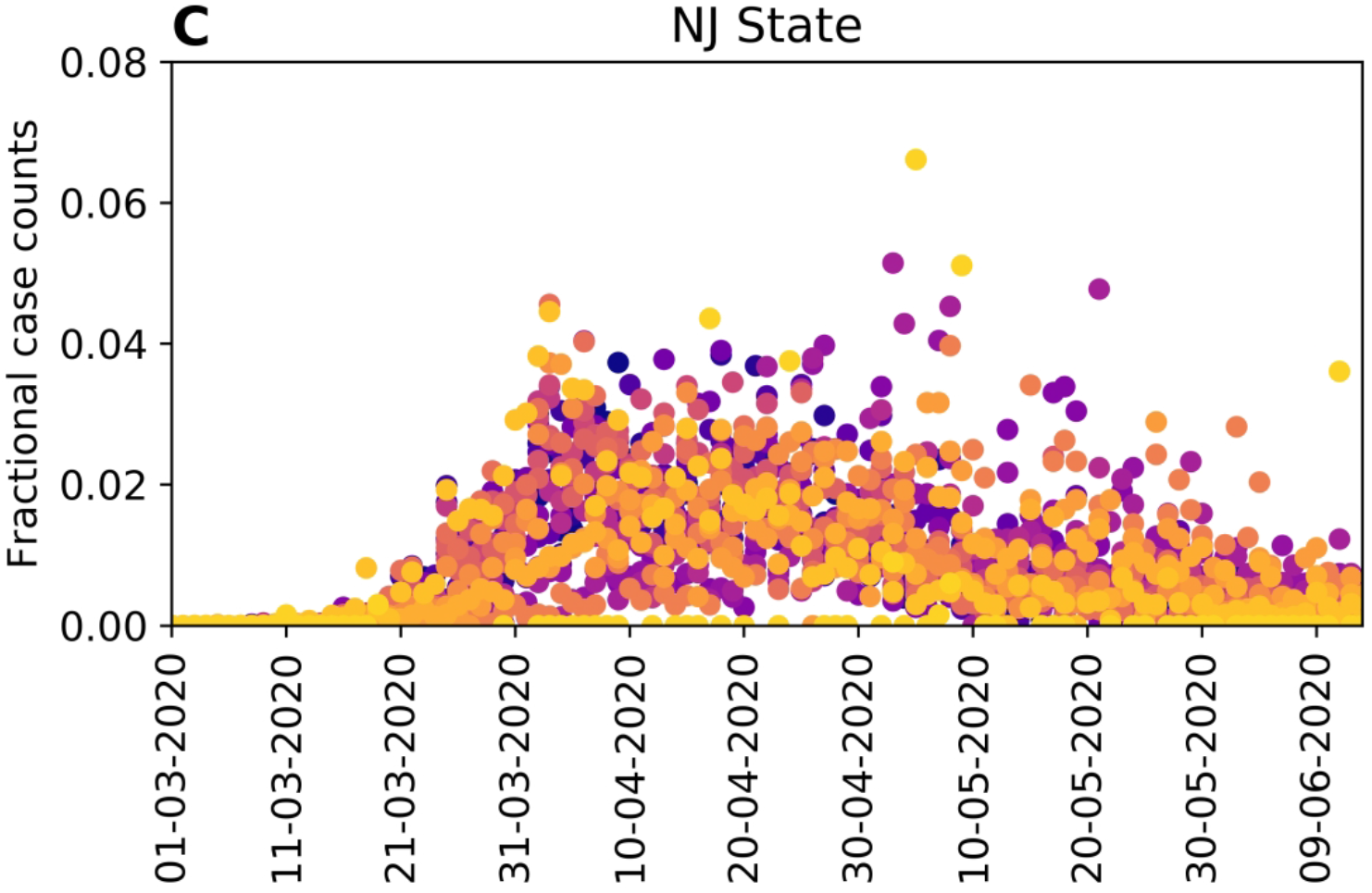
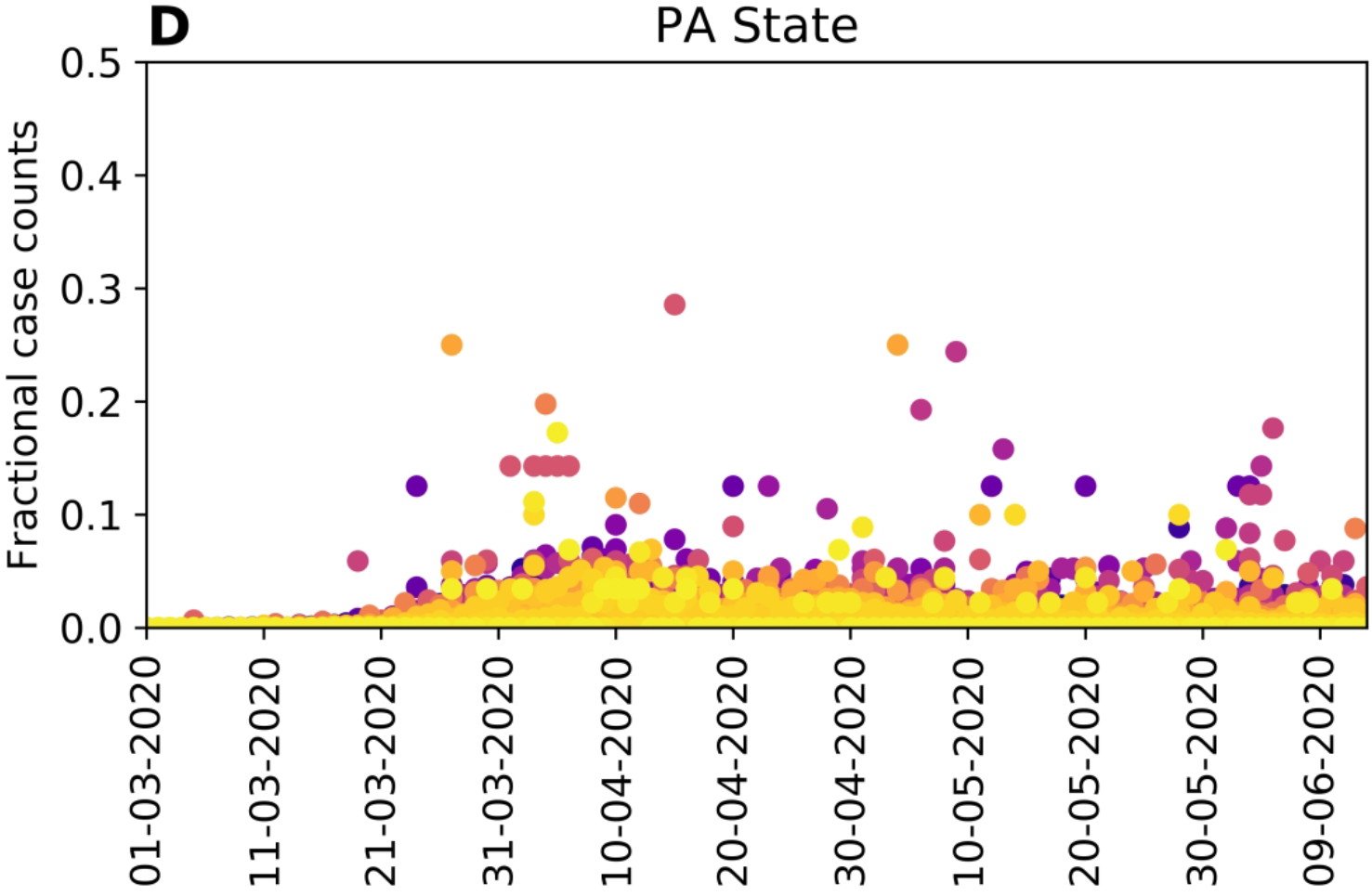
Illustration of Bayesian predictive inference using daily new case counts in the New York Metropolitan Statistical Area. We forecast future daily reports of new COVID-19 cases with rigorous uncertainty quantification (UQ) through online Bayesian learning of model parameters. Each day, using all daily case-reporting data available up to that point, we perform Markov chain Monte Carlo (MCMC) sampling of the posterior distribution for a set of adjustable parameters. Subsampling of the posterior samples then allows us to use the relevant model to generate trajectories of the epidemic curve that account for both parametric and observation uncertainty. The entire shaded region indicates the 95% credible interval for predictions of daily case reports. In other words, the central 95% of all predictions lie within the shaded region. The color-coded bands within the shaded region indicate other credible intervals, as indicated in the legend.

Predictive inferences for all 15 MSAs of interest are shown in Figure 4. The predictions of Figure 4 are conditioned on the compartmental model with *n* = 0. These results demonstrate that, for the timeframe of interest, the compartmental model with *n* = 0 is capable of reproducing many but not all of the empirical epidemic curves for the MSAs of interest, which vary in shape.

**Figure 4.**
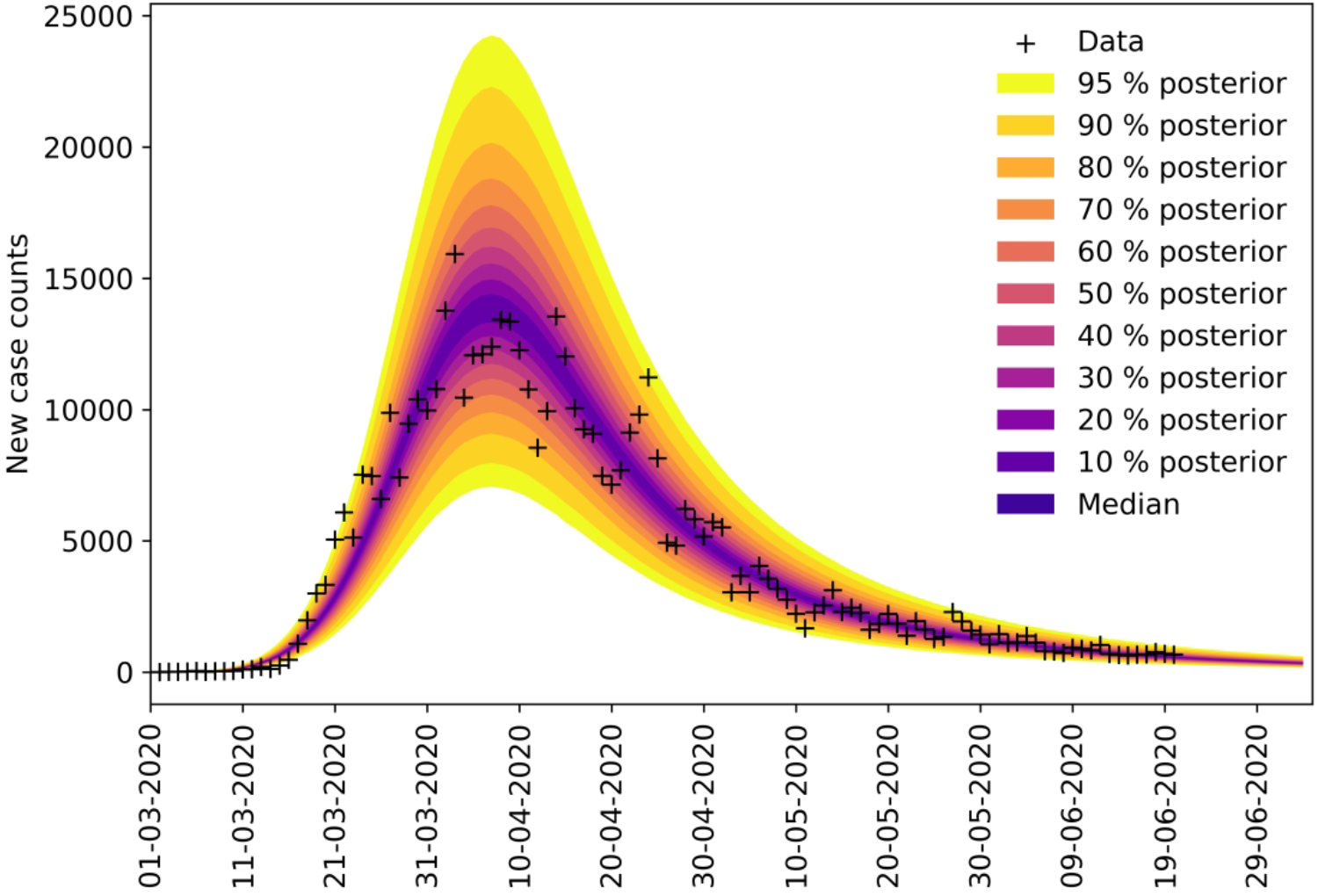
Bayesian predictive inferences for the 15 most populous metropolitan statistical areas (MSAs) in the United States. Predictions are conditioned on the compartmental model with structure defined by *n* = 0, i.e., the simplest version of the model, which accounts for only a single period of social distancing.

Recall that predictive inferences are made daily. In Figure 5, we show predictive inferences for New York City and Phoenix made over a series of progressively later dates. These results illustrate that accurate short-term predictions are possible but continual updating of parameter estimates is required to maintain accuracy. In Videos 1 and 2, daily predictions for New York City and Phoenix, respectively, are shown as animations.

**Figure 5.**
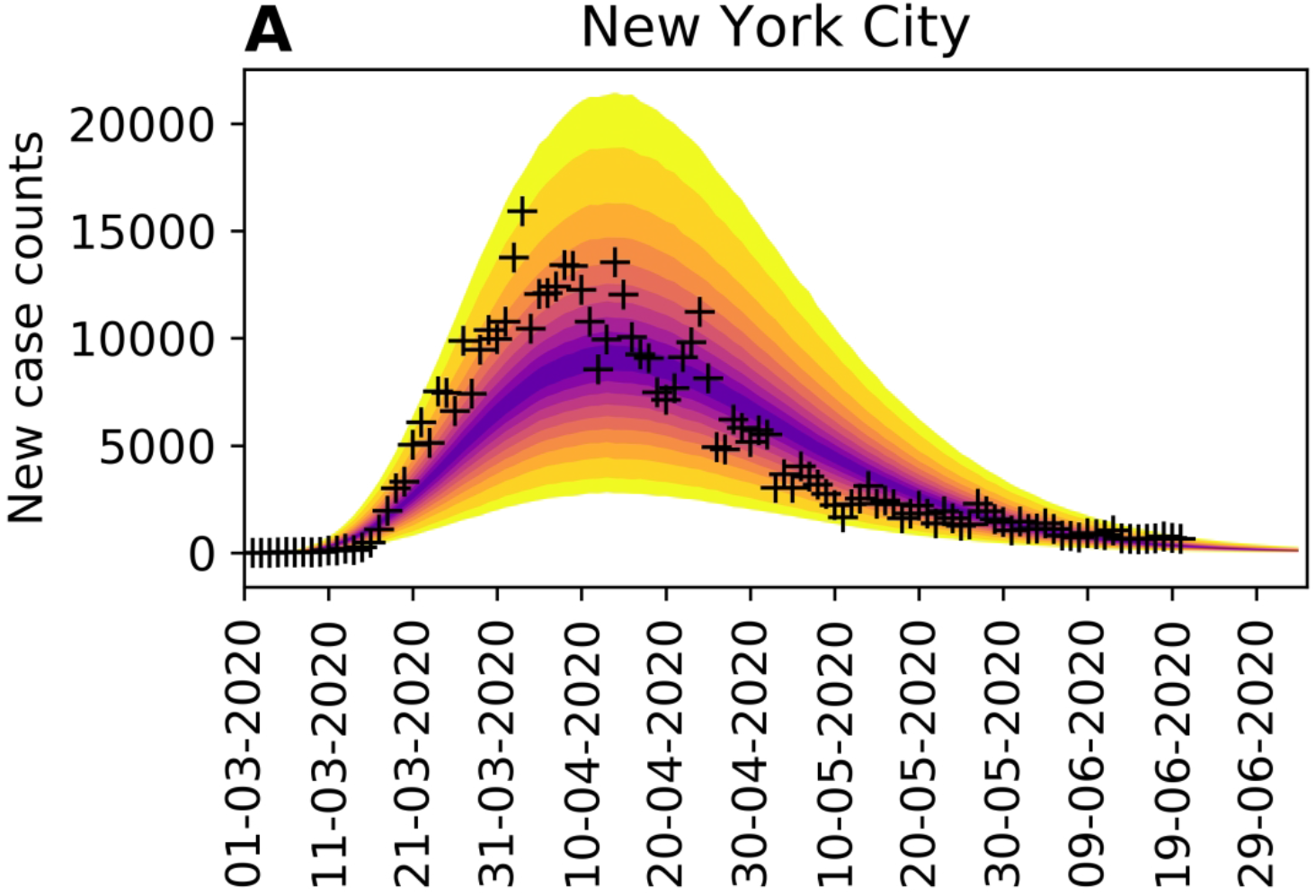
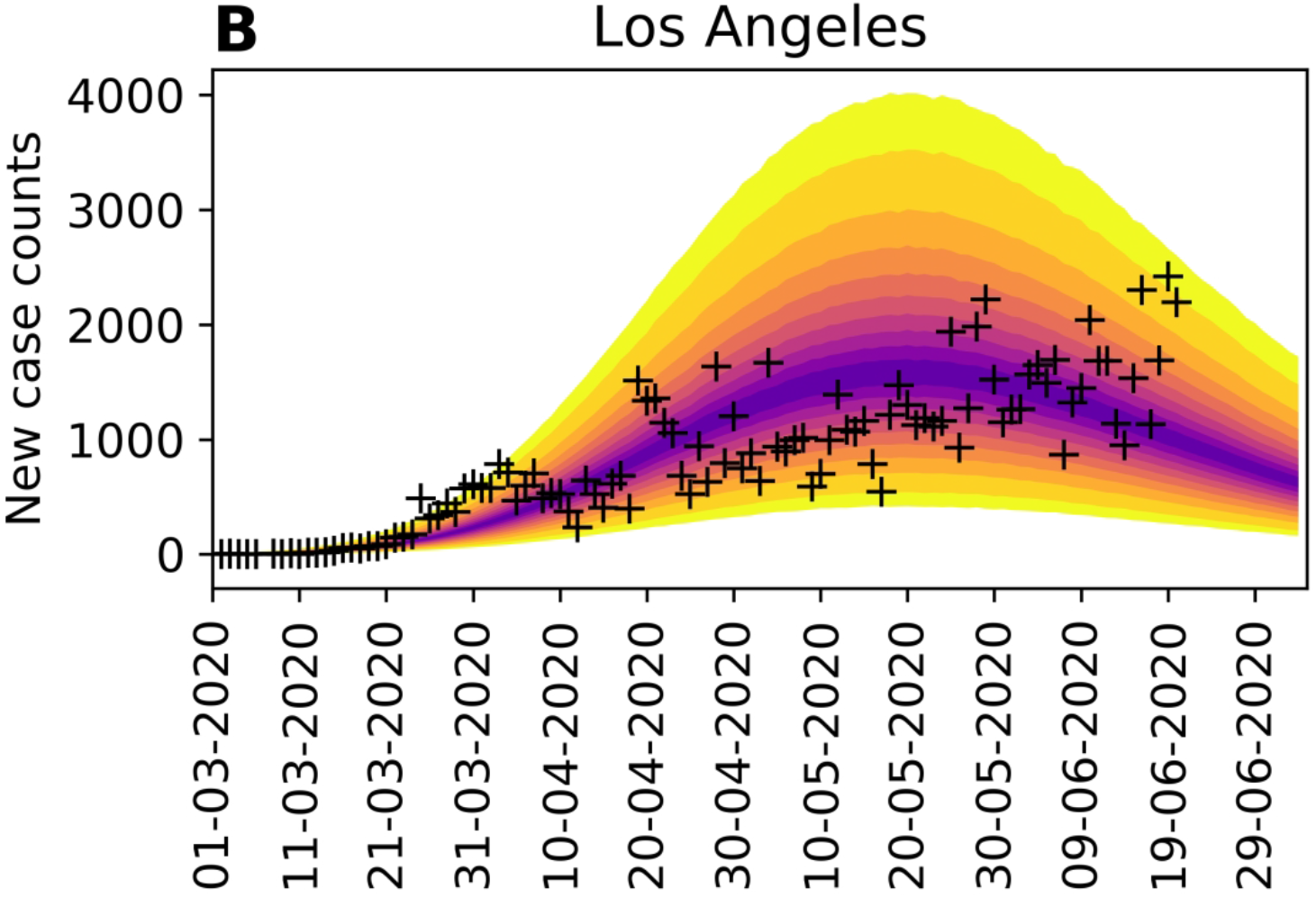
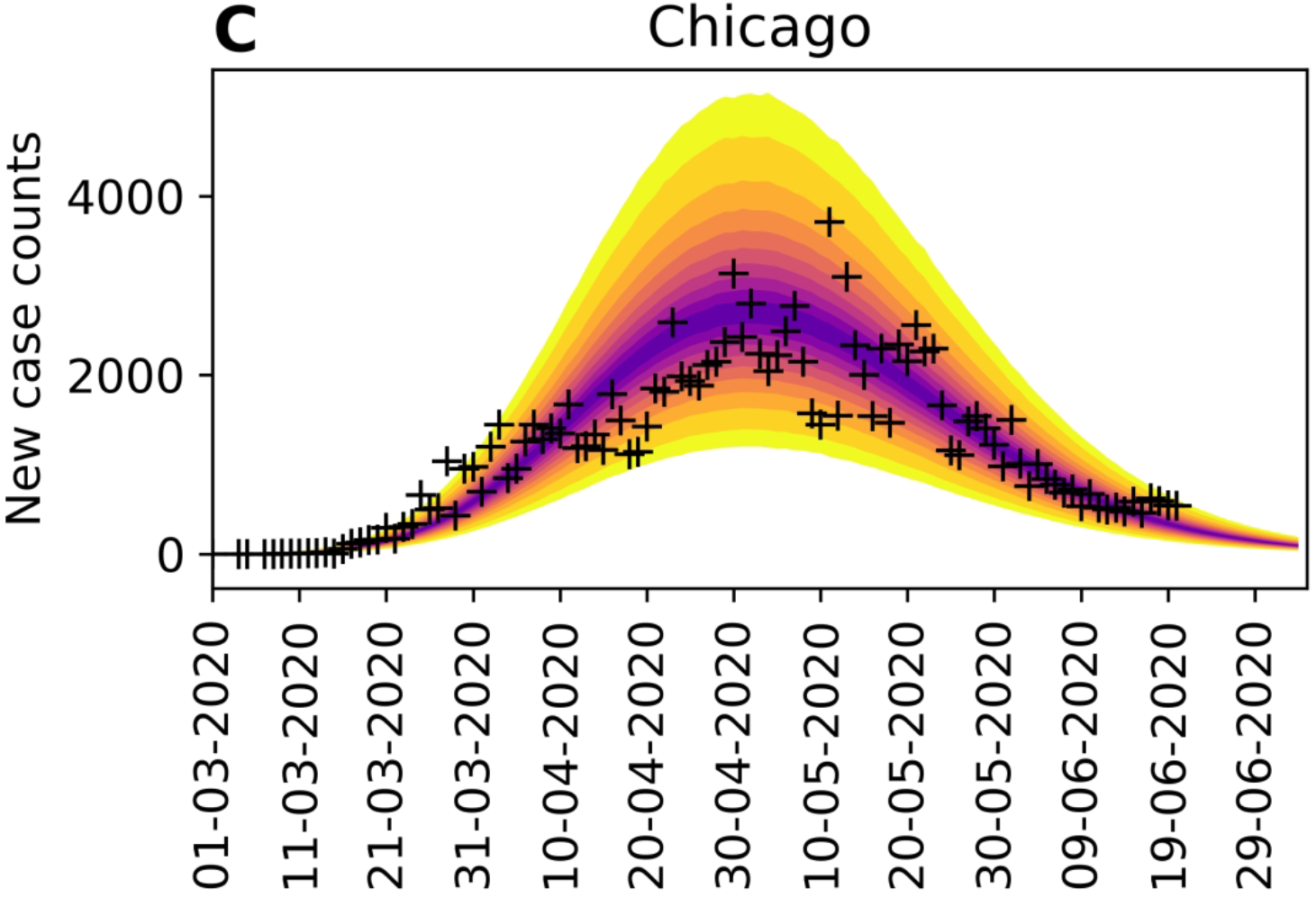
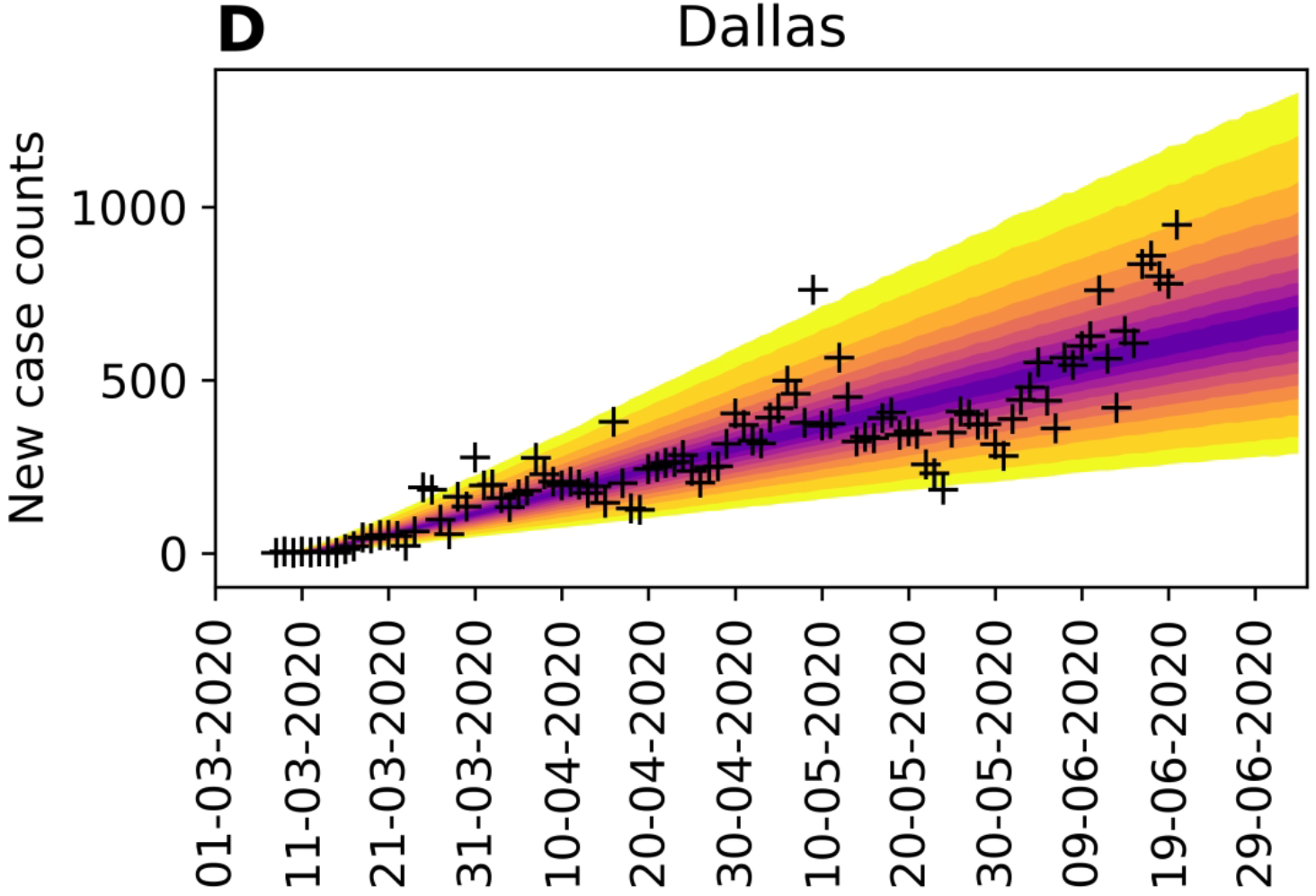
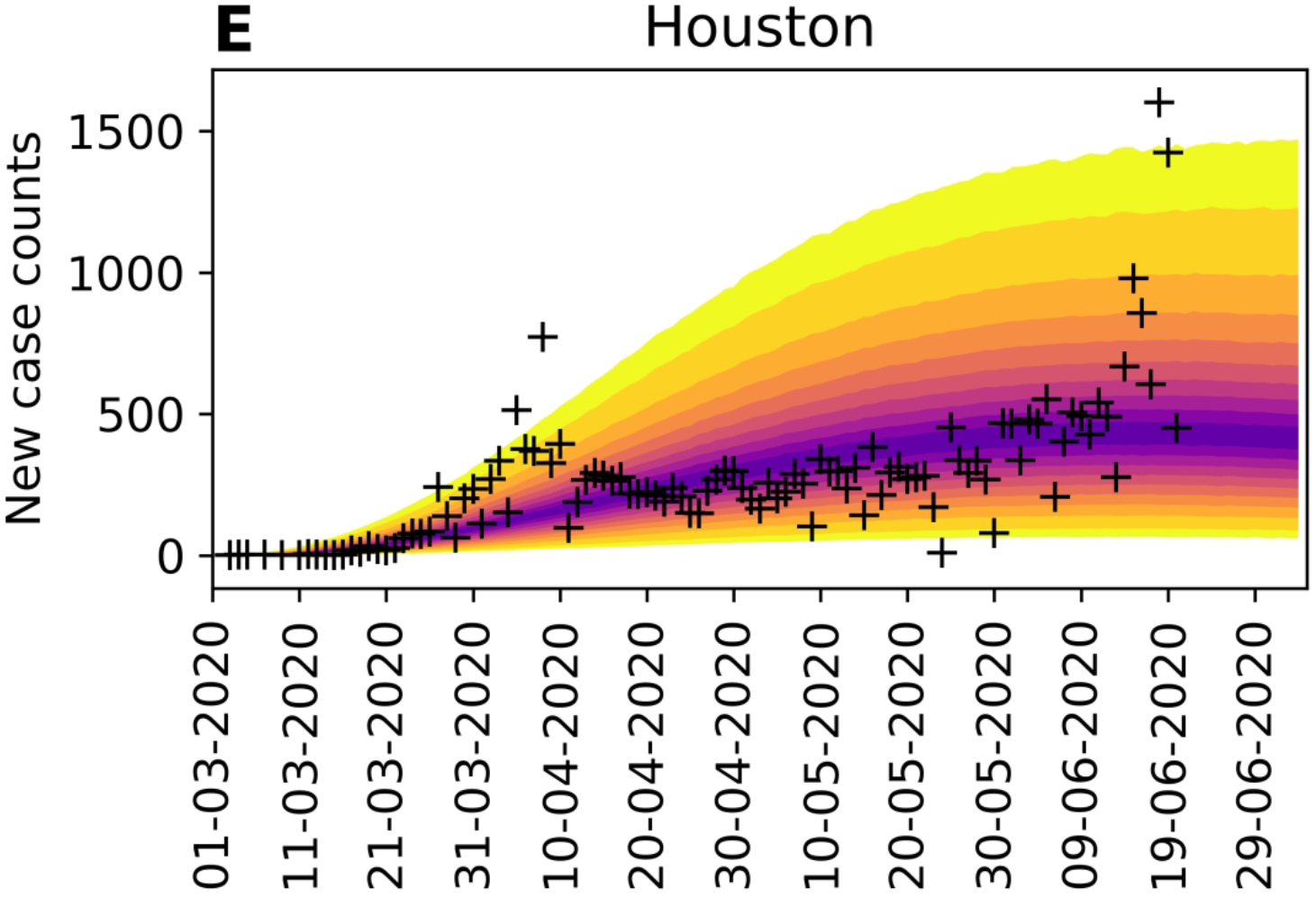
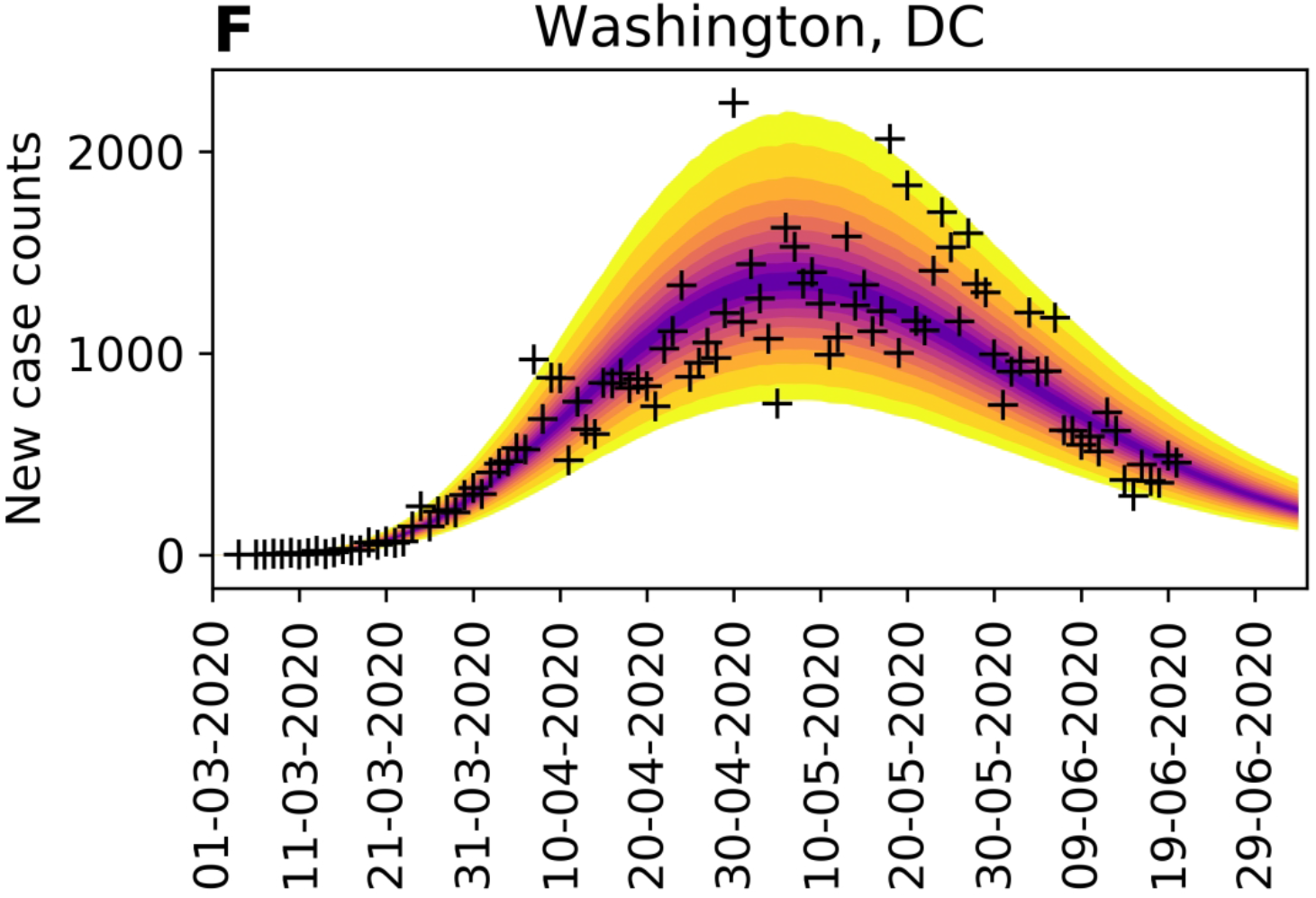
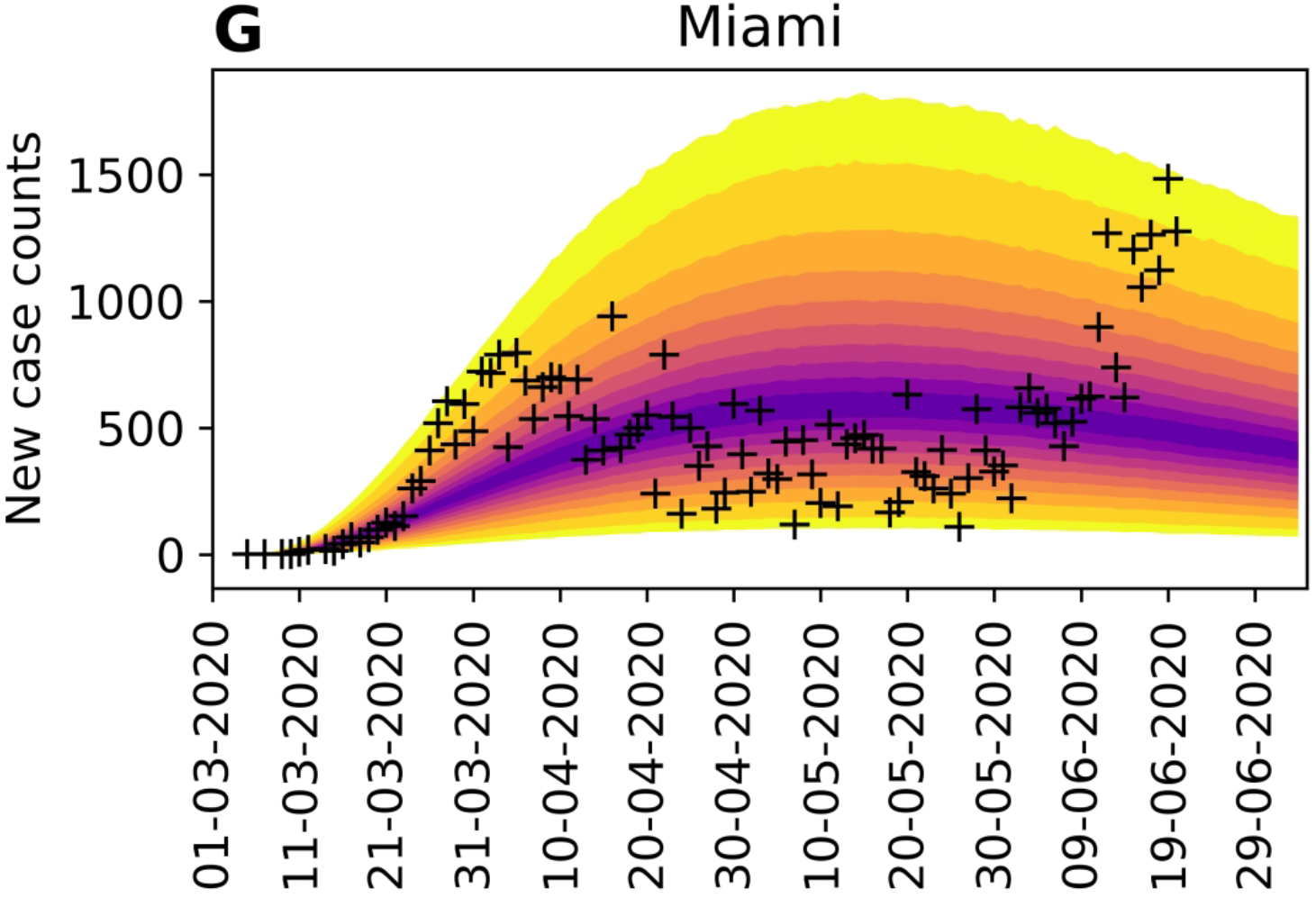
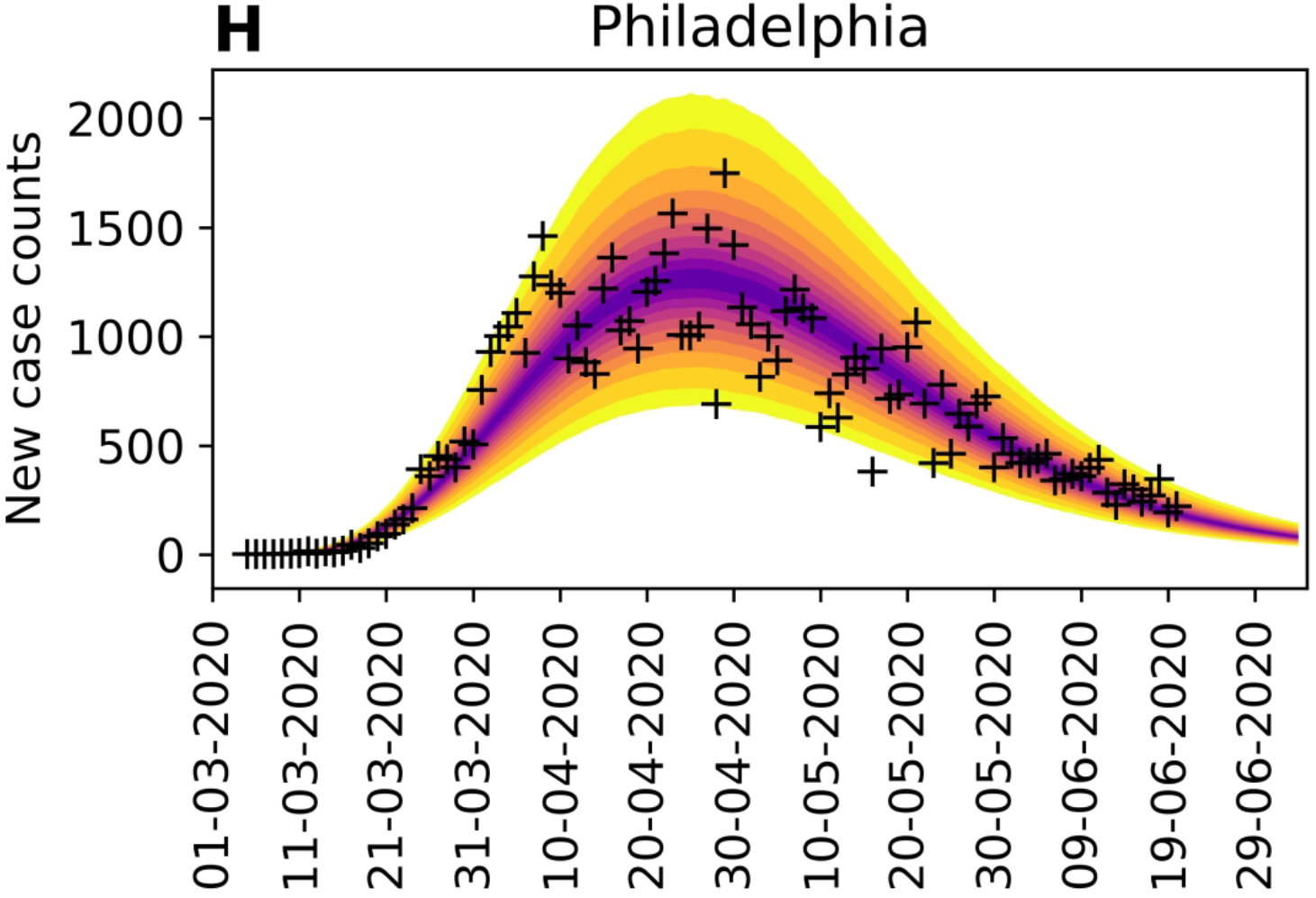
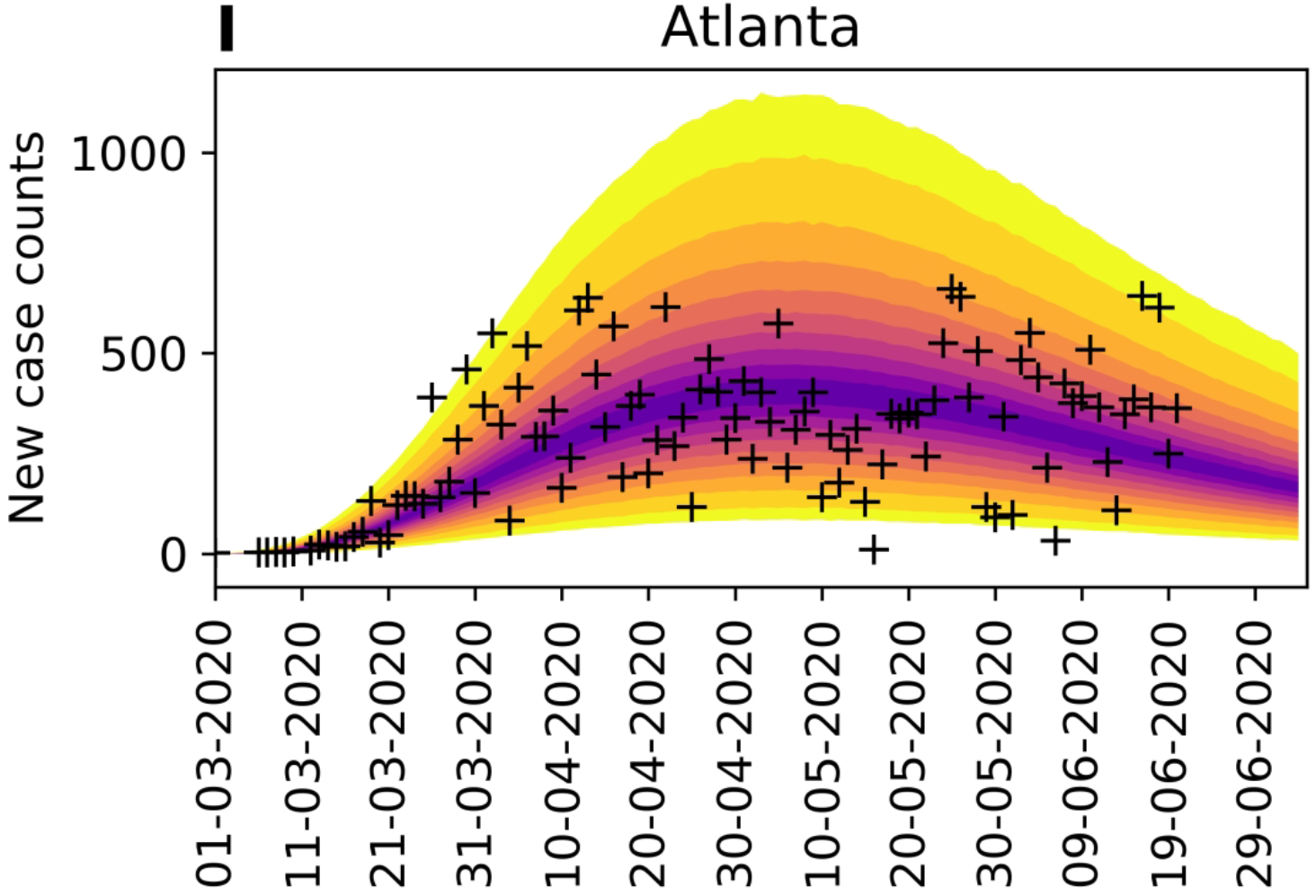
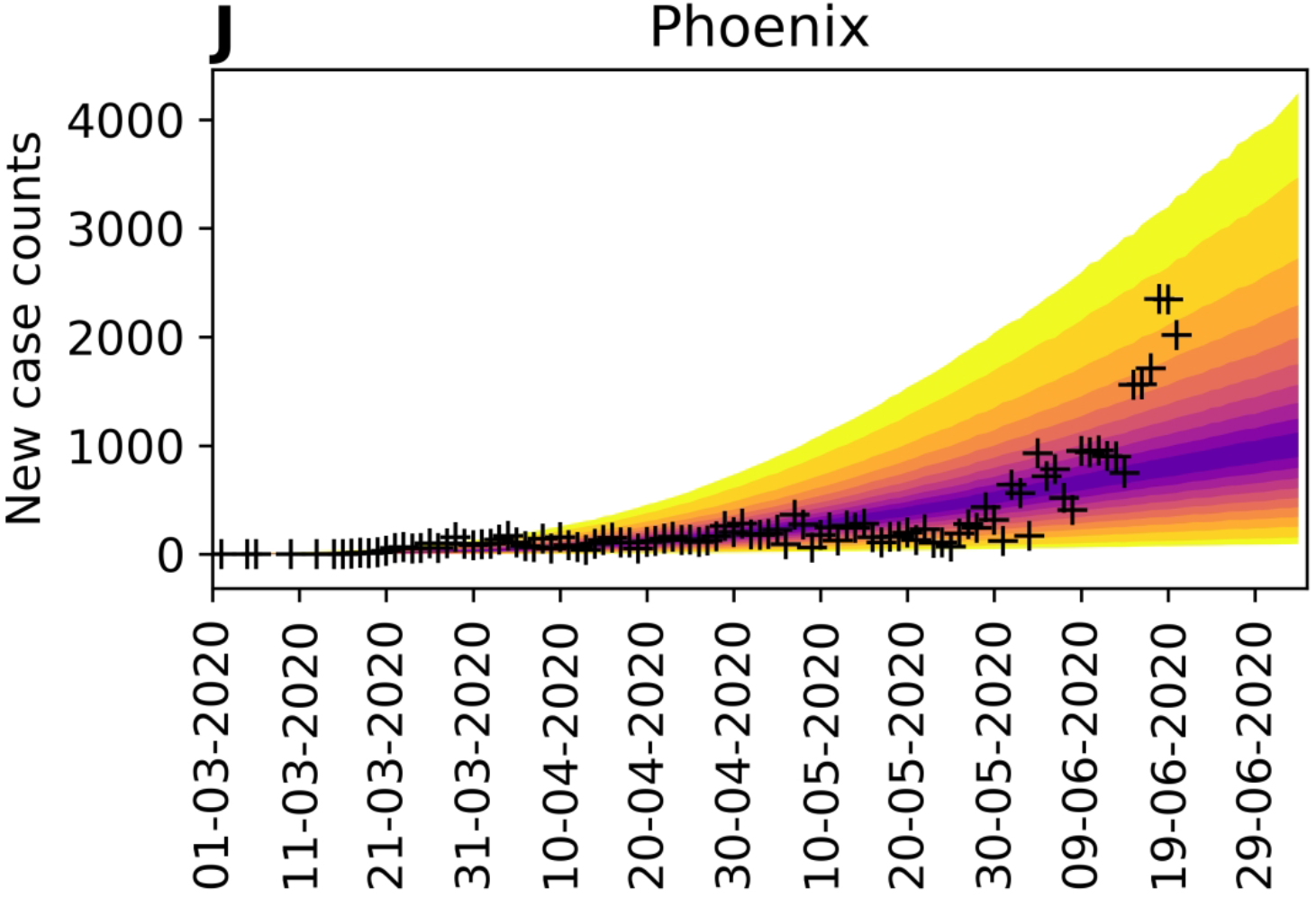
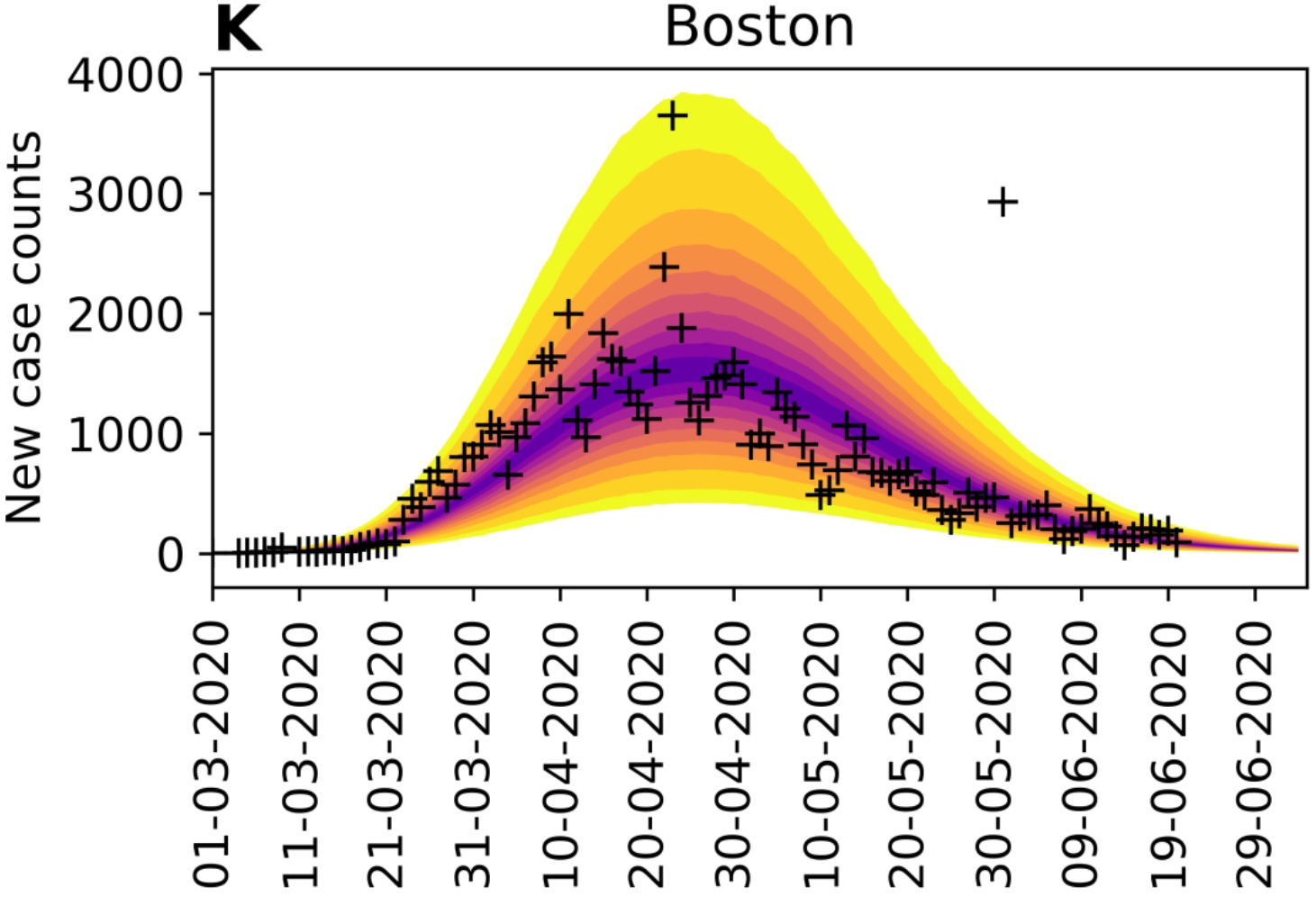
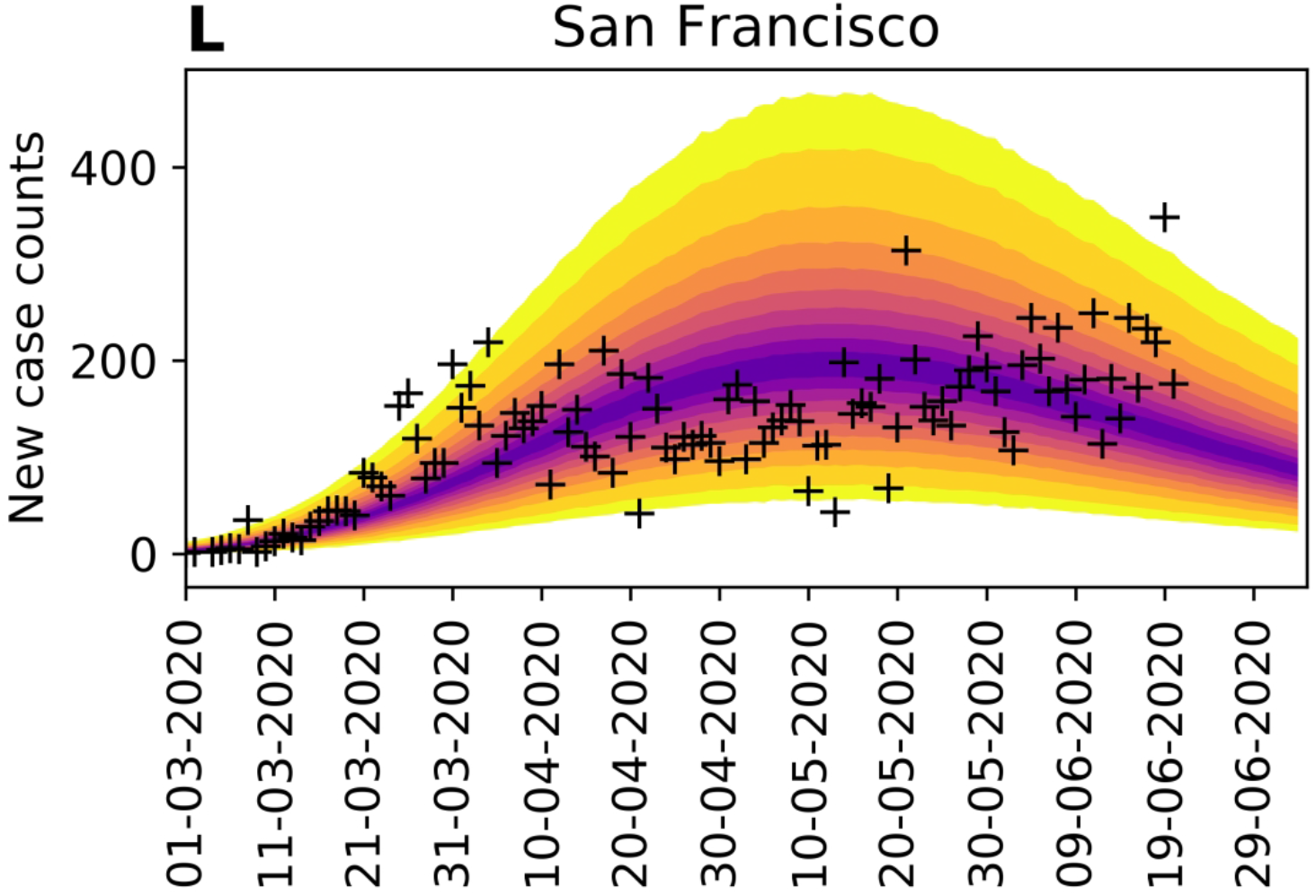
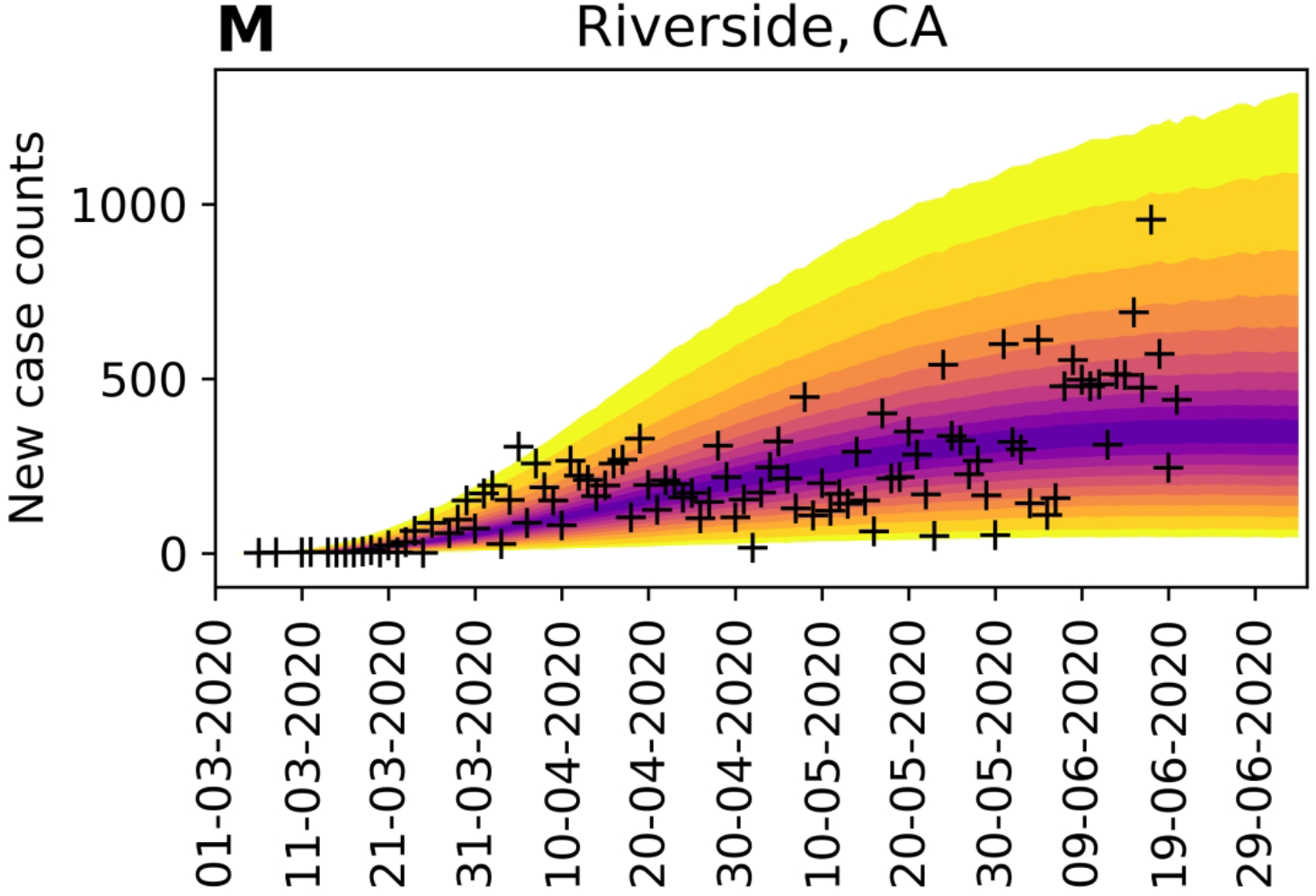
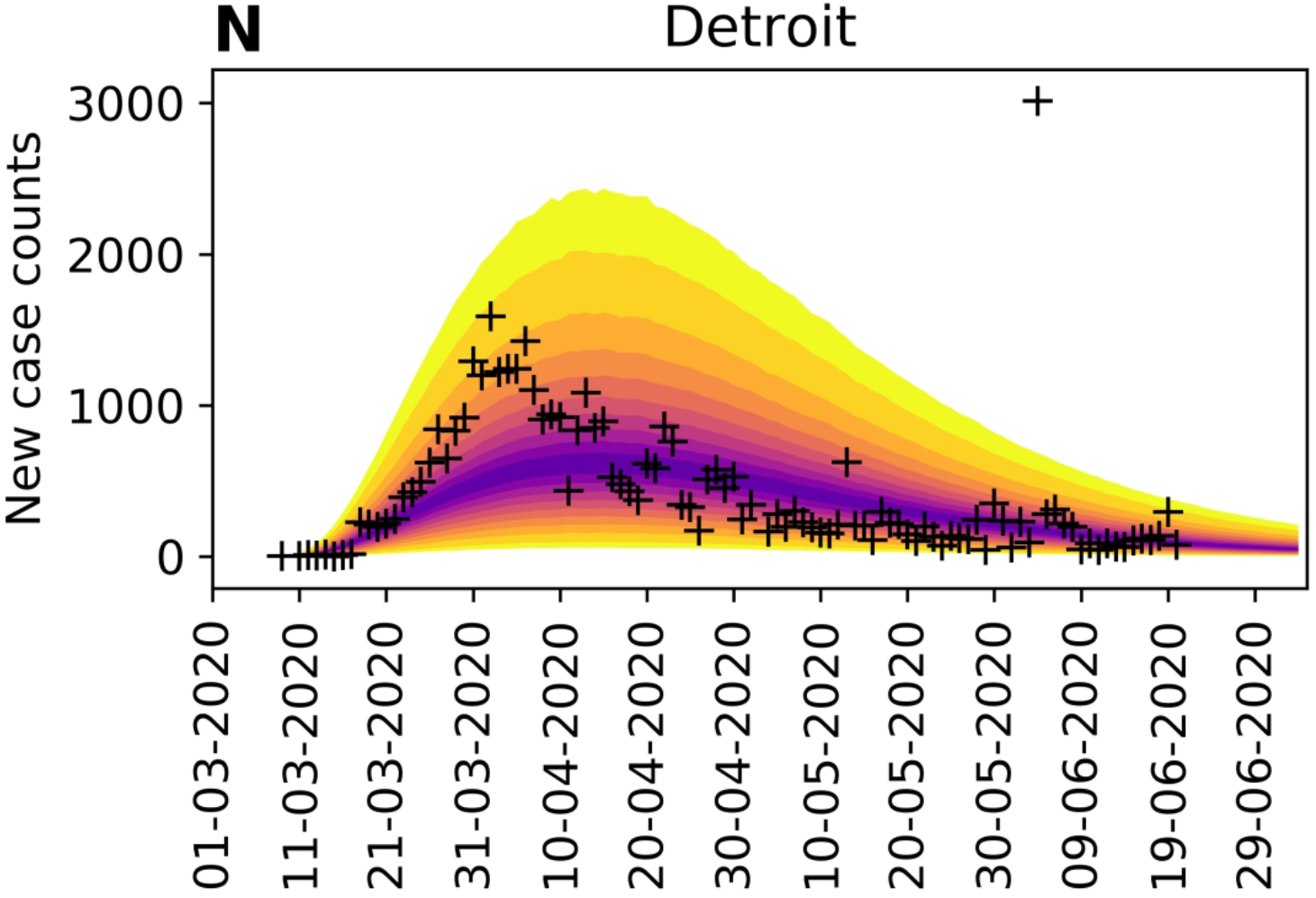
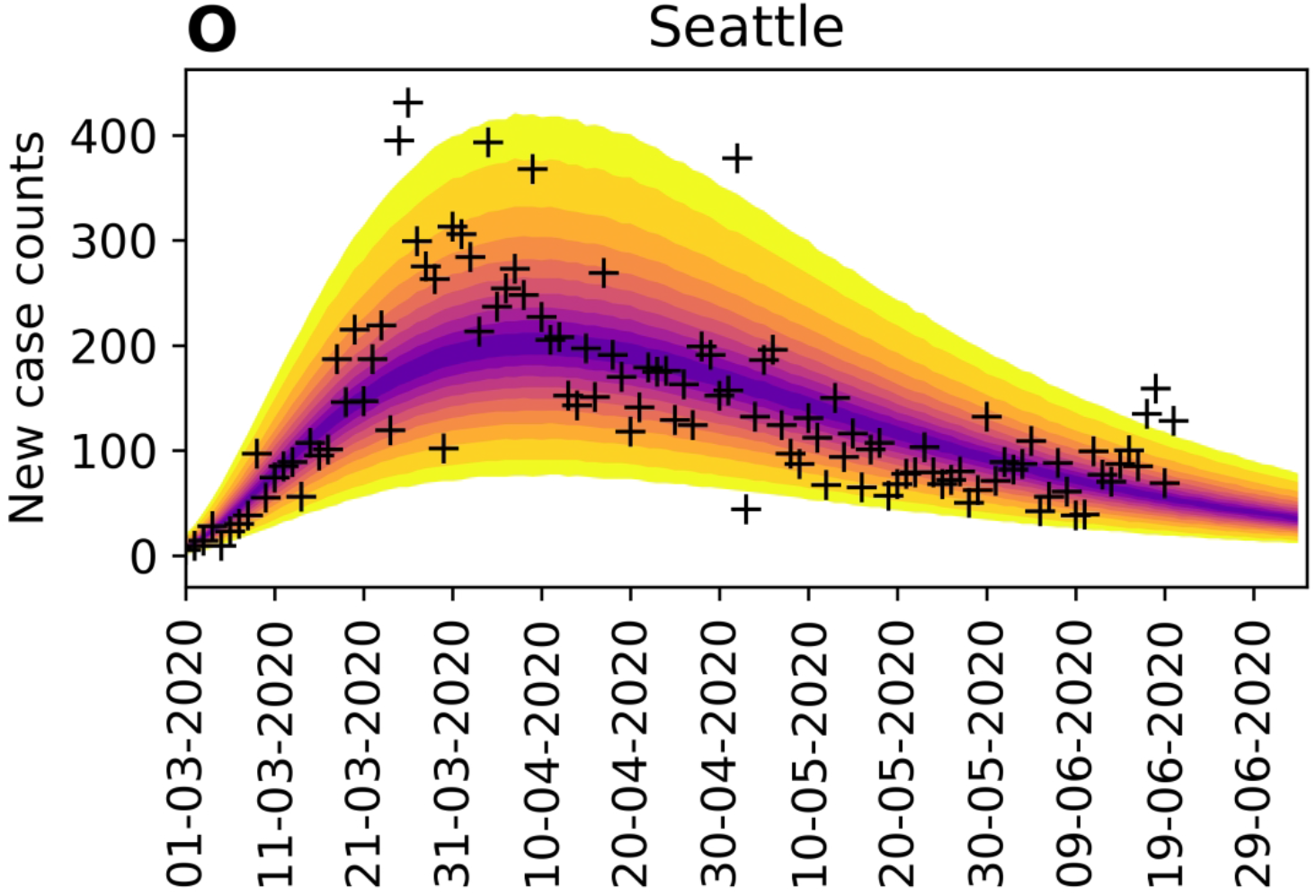
The necessity of online learning. (A)–(E) Shown are predictions for the New York City metropolitan statistical area (MSA) made over a series of progressively later dates, as indicated. (F)–(J) Shown are predictions for the Phoenix MSA made over a series of progressively later dates, as indicated. Predictive inferences, which are all conditioned on the compartmental model, are data-driven. Accurate short-term predictions are possible but continual updating of parameter estimates is required to maintain accuracy.

In practice, we find that the adjustable parameters of the compartmental model have identifiable values, meaning that their marginal posteriors are unimodal. This finding is illustrated in Figure 6, which displays a 7 × 7 matrix of 1- and 2-dimensional projections of the 7-dimensional MCMC posterior samples underlying predictive inferences for New York City. In Figure 6, plots of marginal posteriors are shown on the diagonal extending from top left to bottom right. The other 2-dimensional plots reveal correlations between pairs of parameter estimates (if any). In the context of a deterministic model, the significance of identifiability is that, despite uncertainties in parameter estimates, we can expect predictive inferences of daily new-case reports to cluster around a central trajectory. The results shown in Figure 6 are representative. We routinely recover unimodal marginal posteriors. However, it should be noted that we do not have a mathematical proof of identifiability.

**Figure 6.**
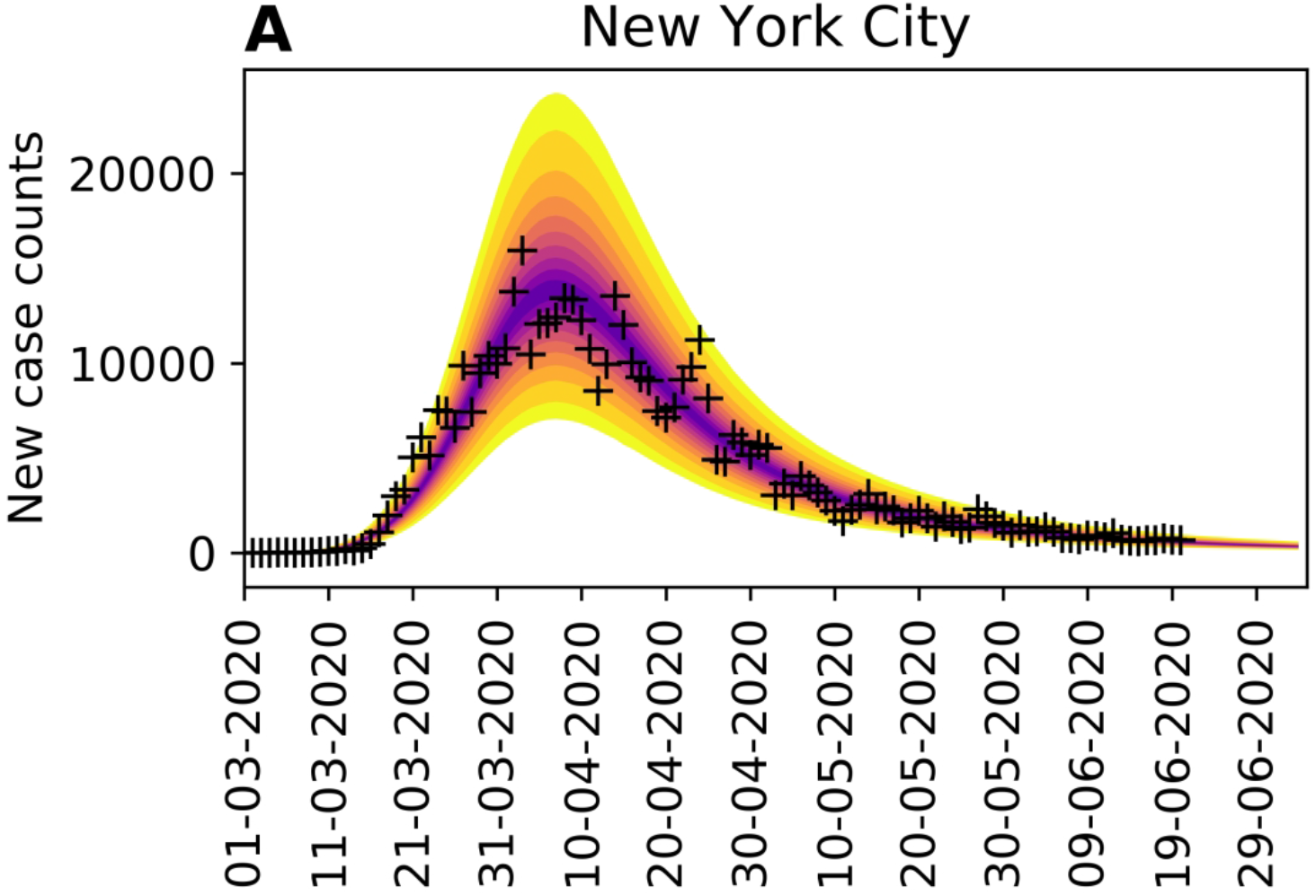
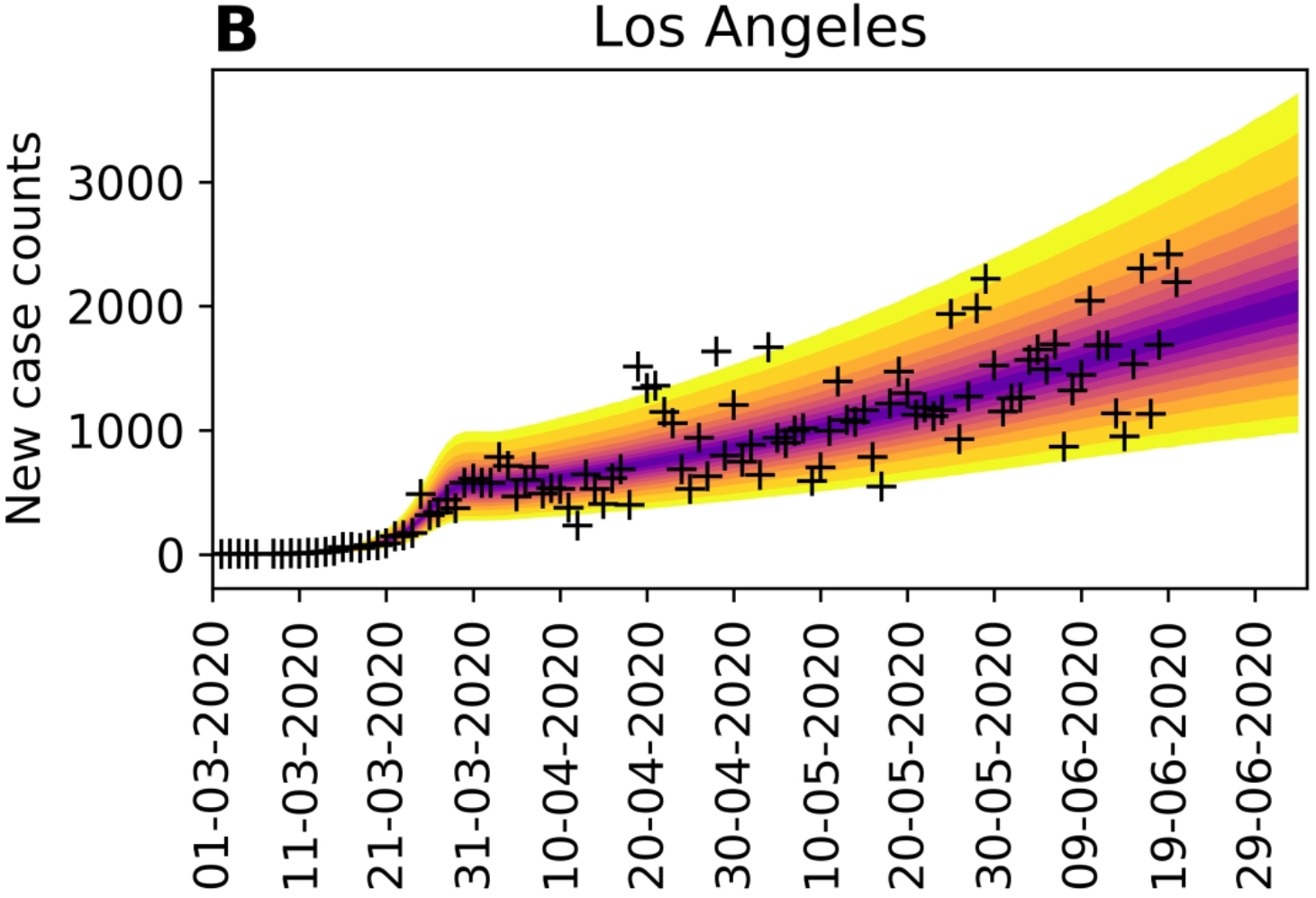
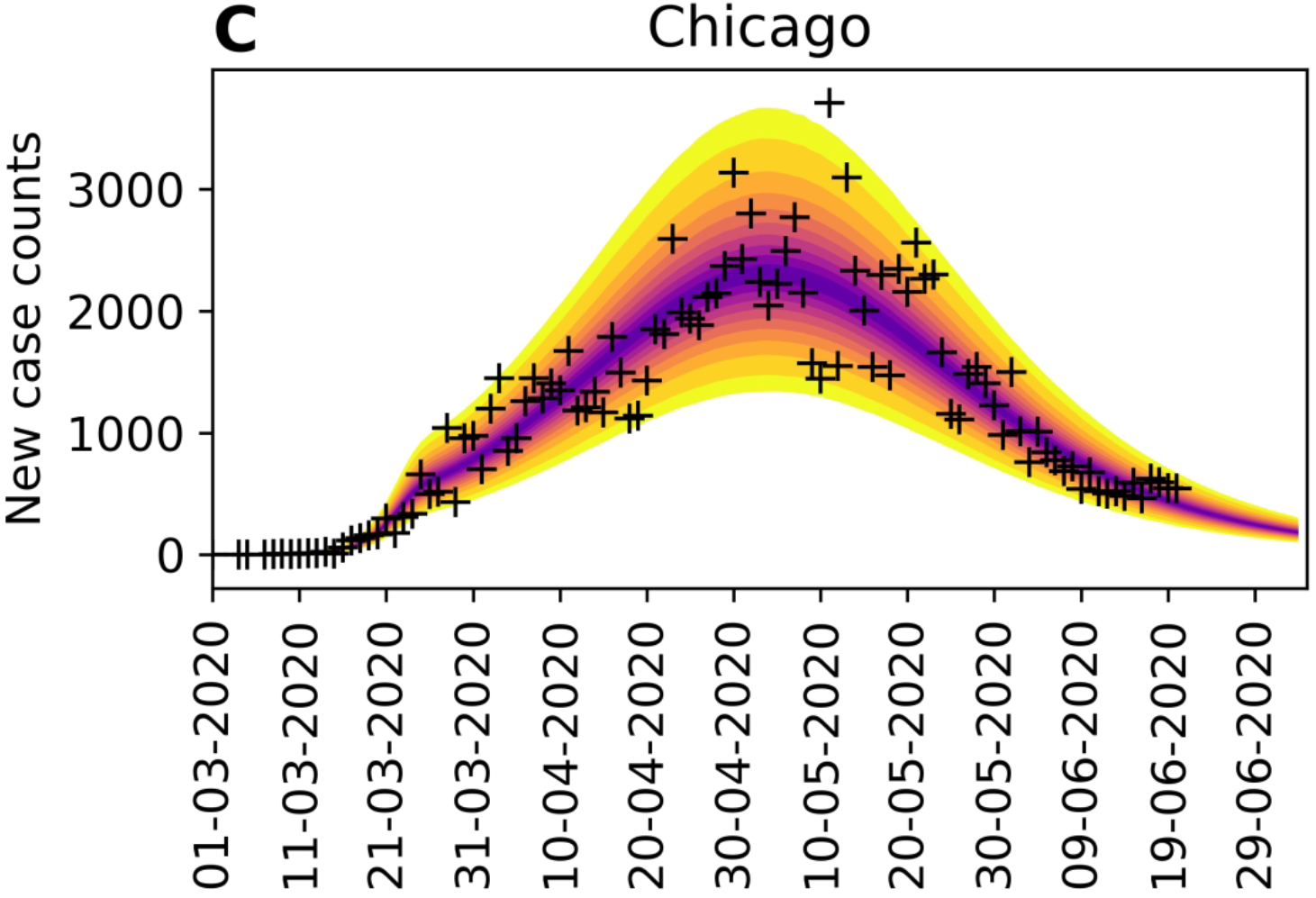
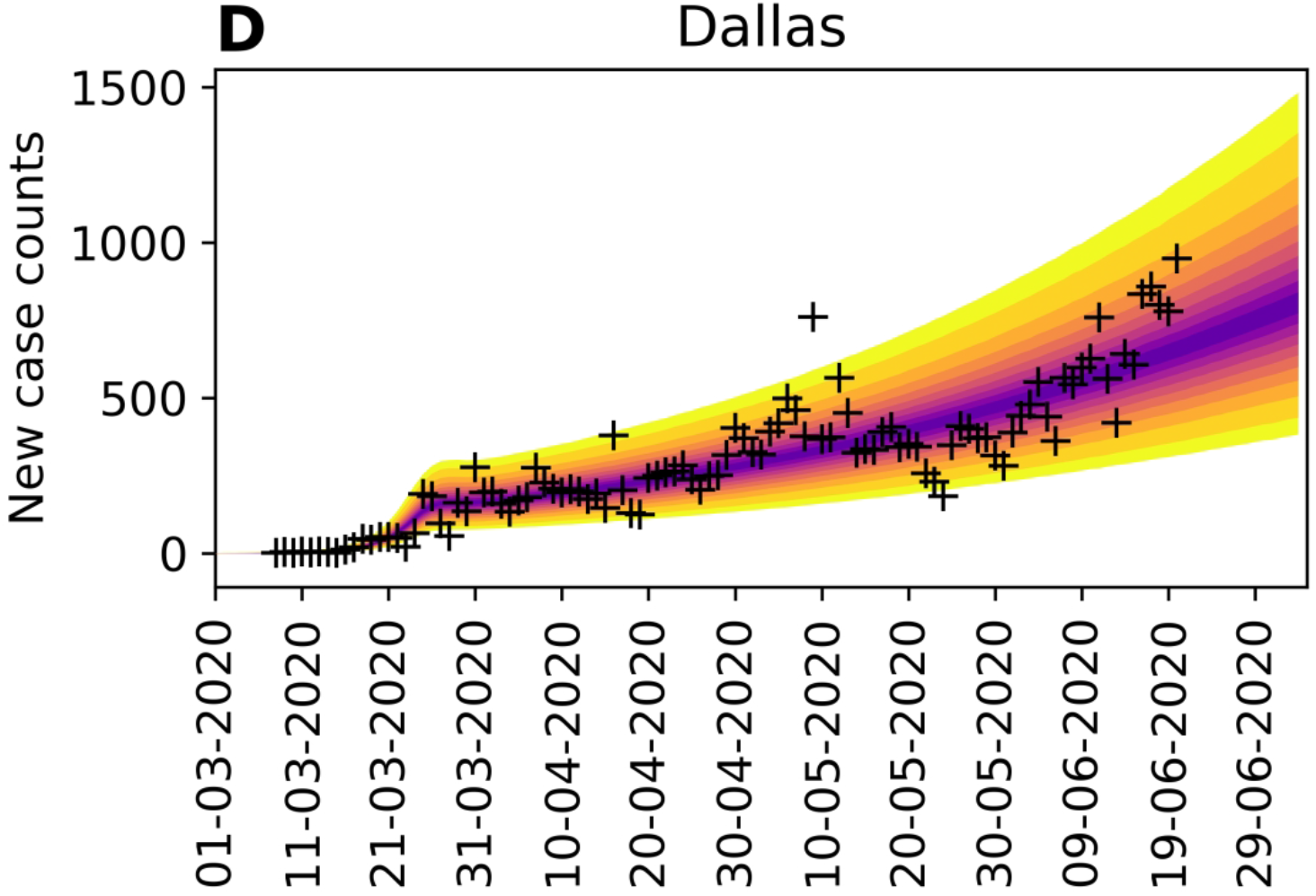
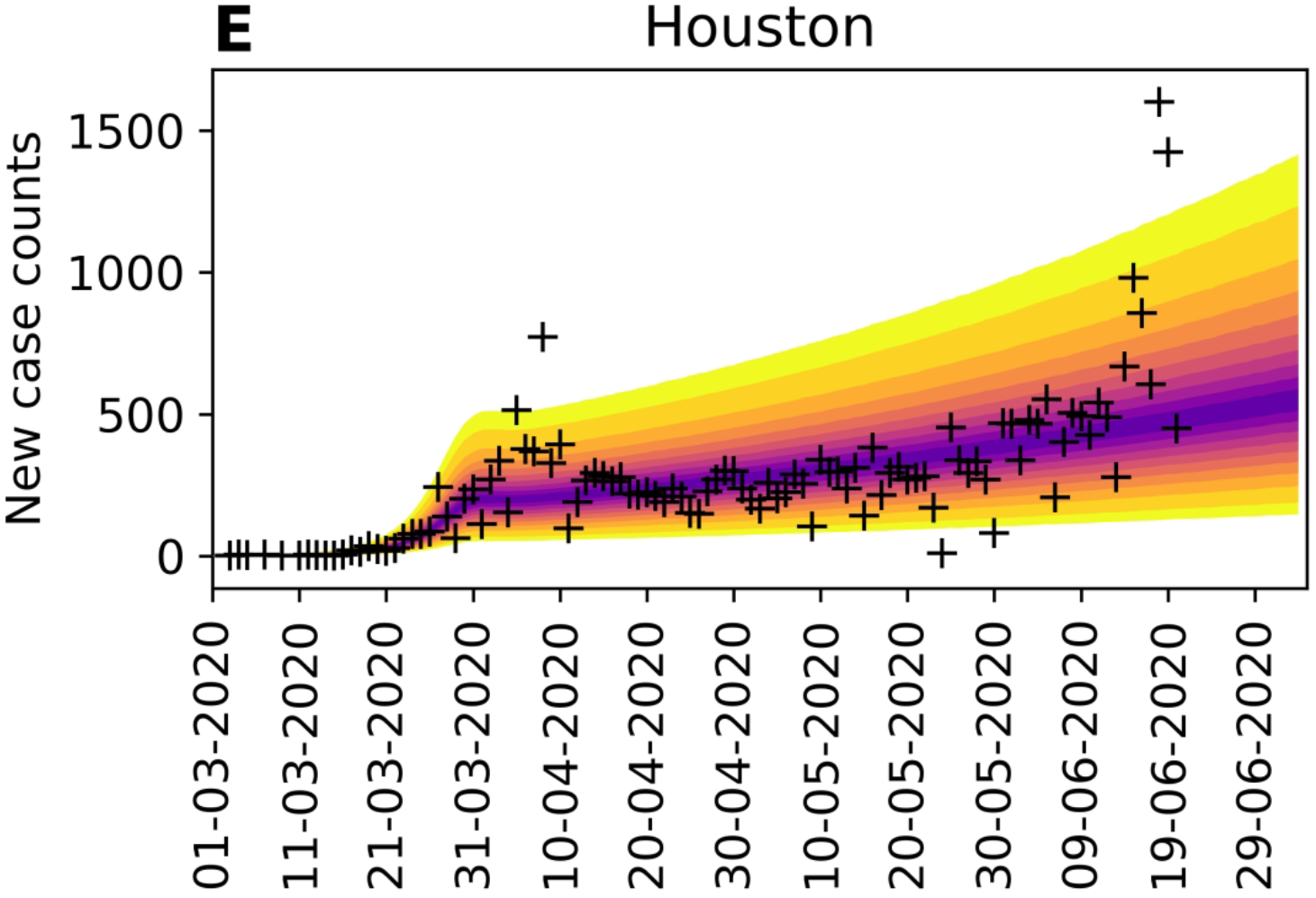
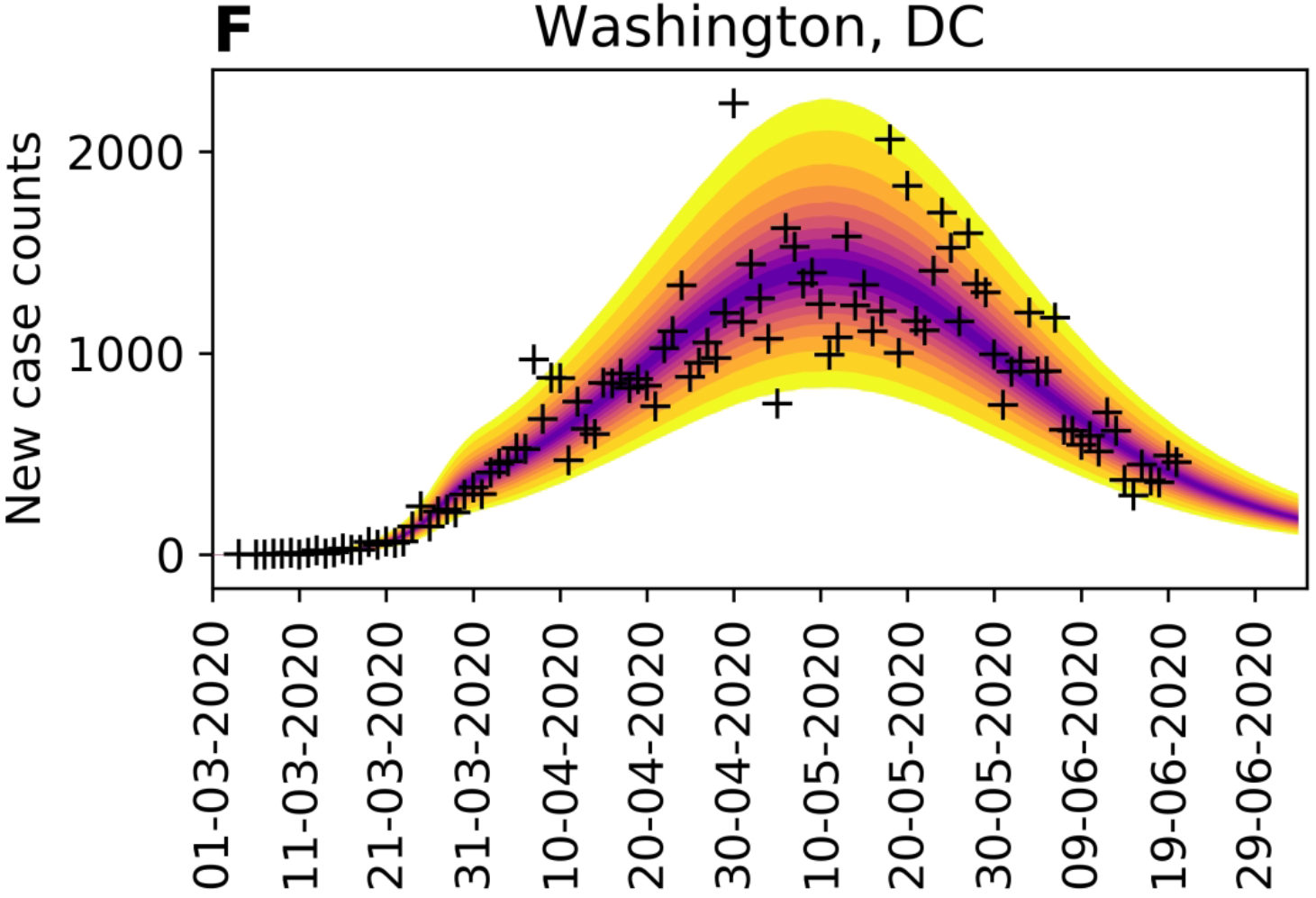
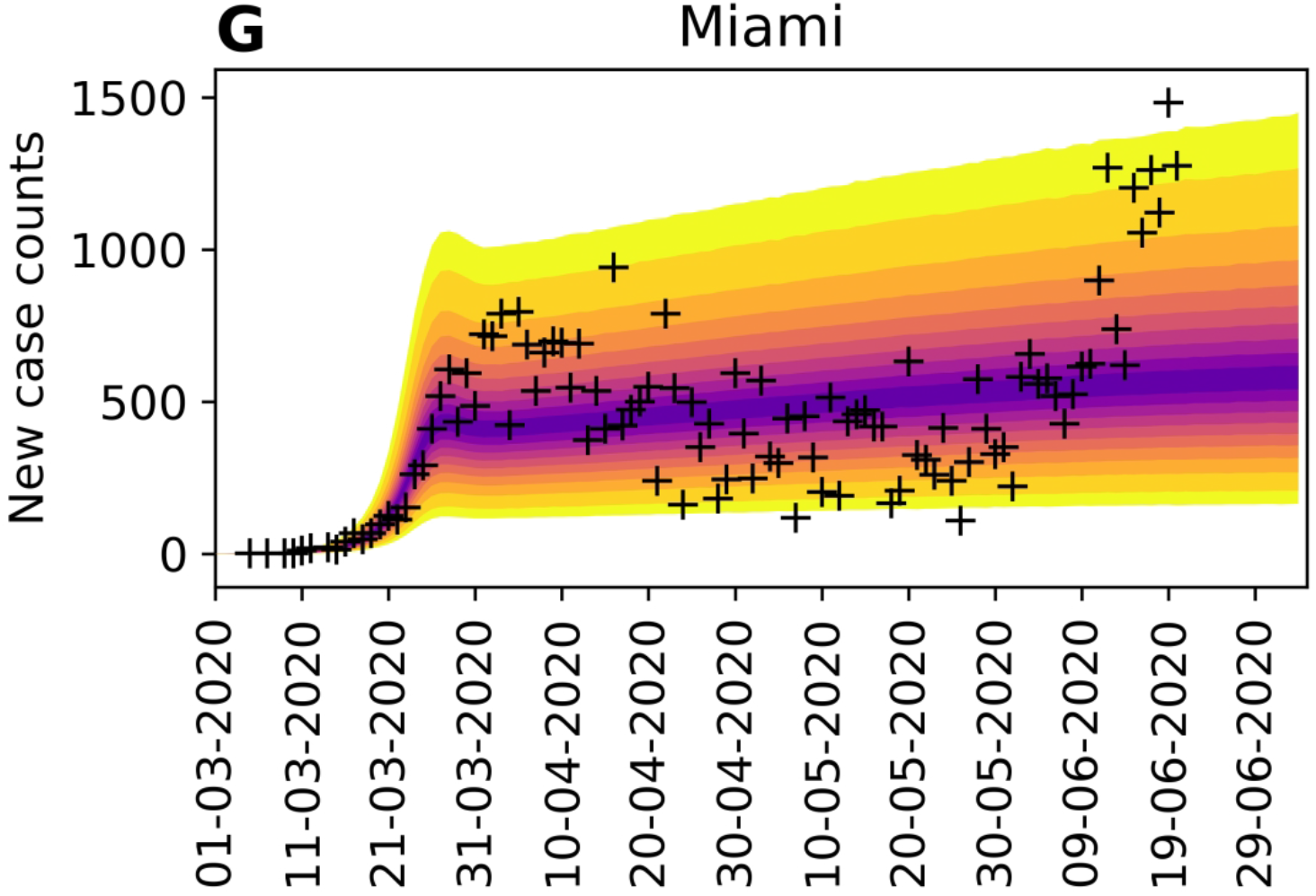
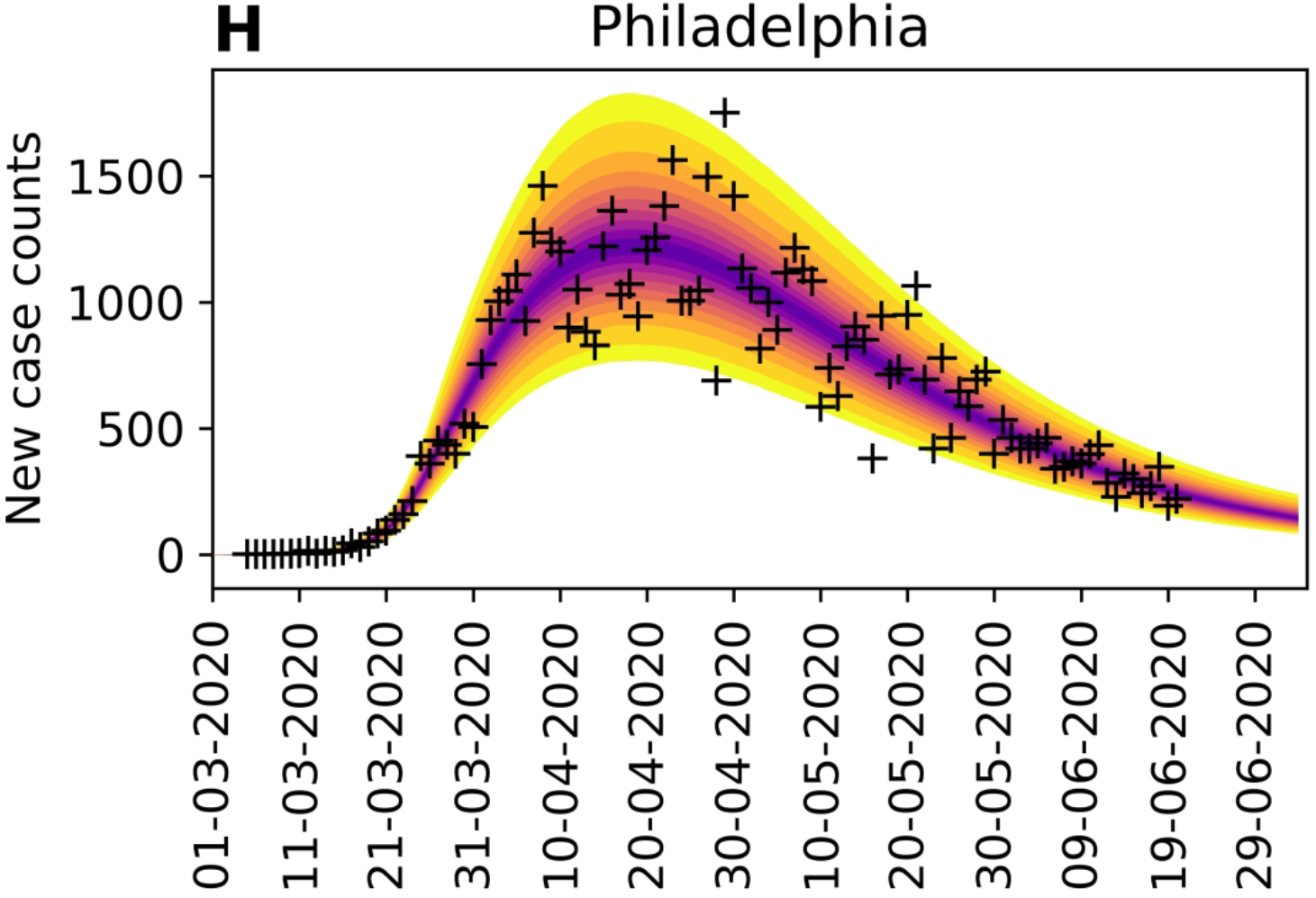
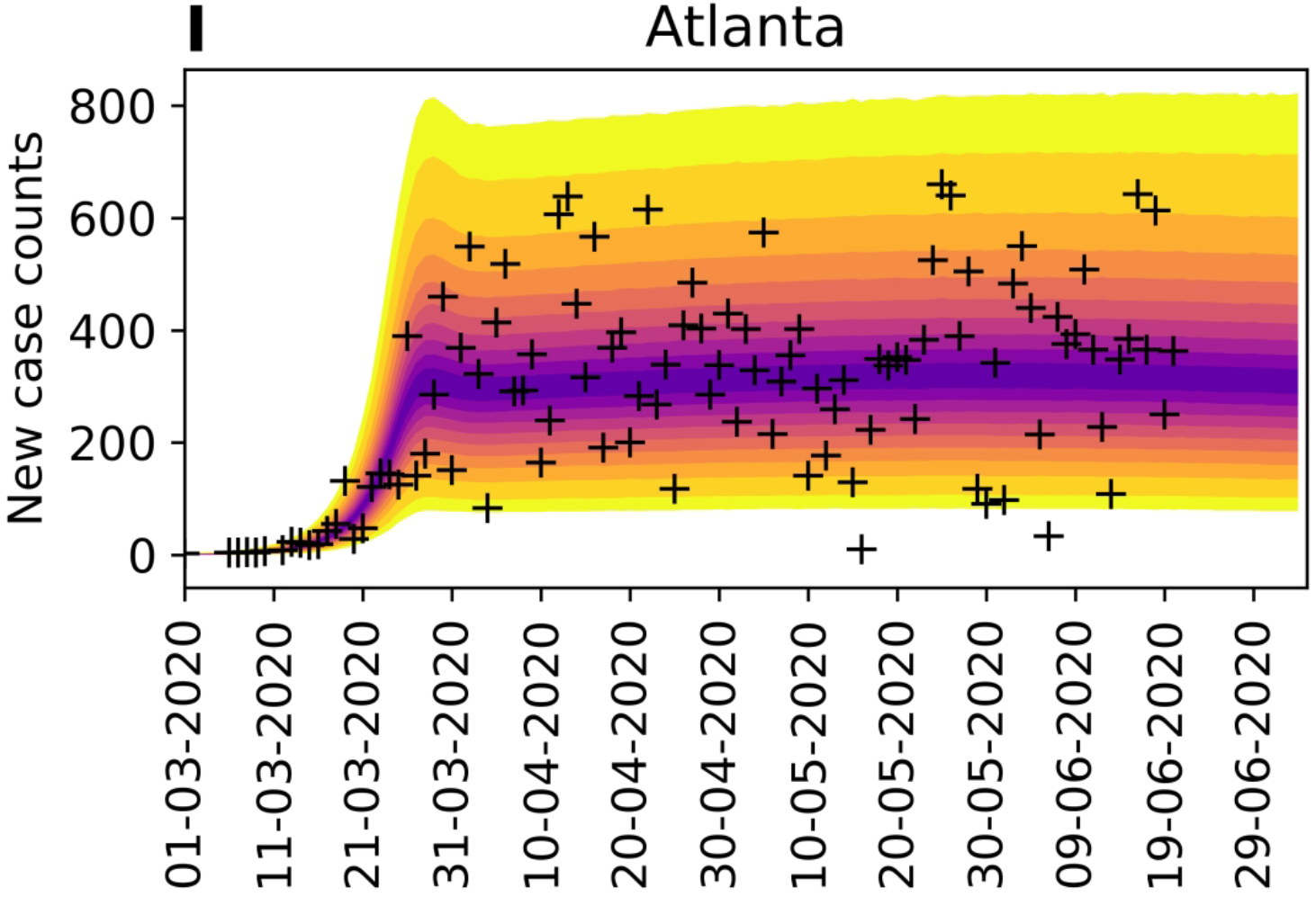
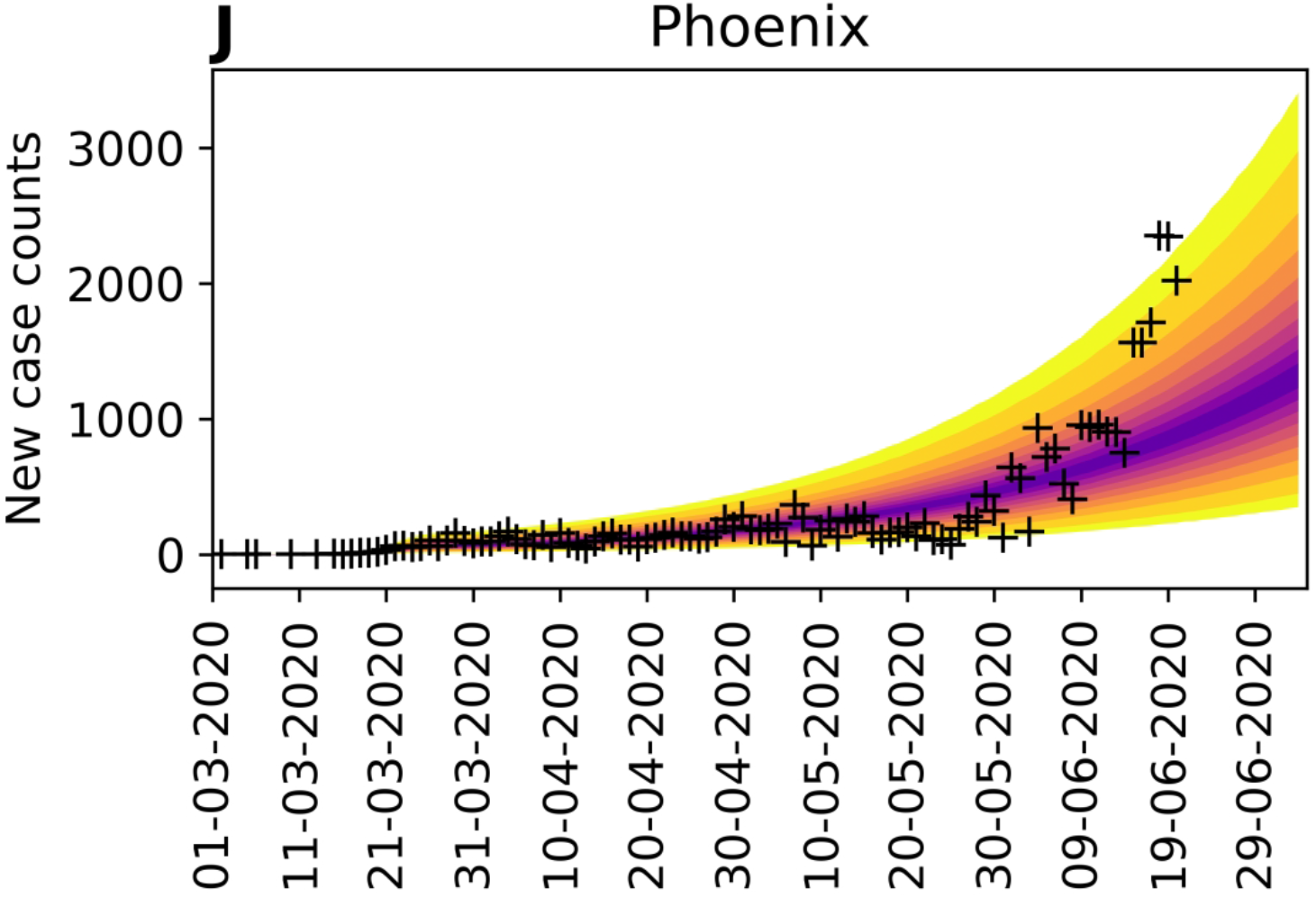
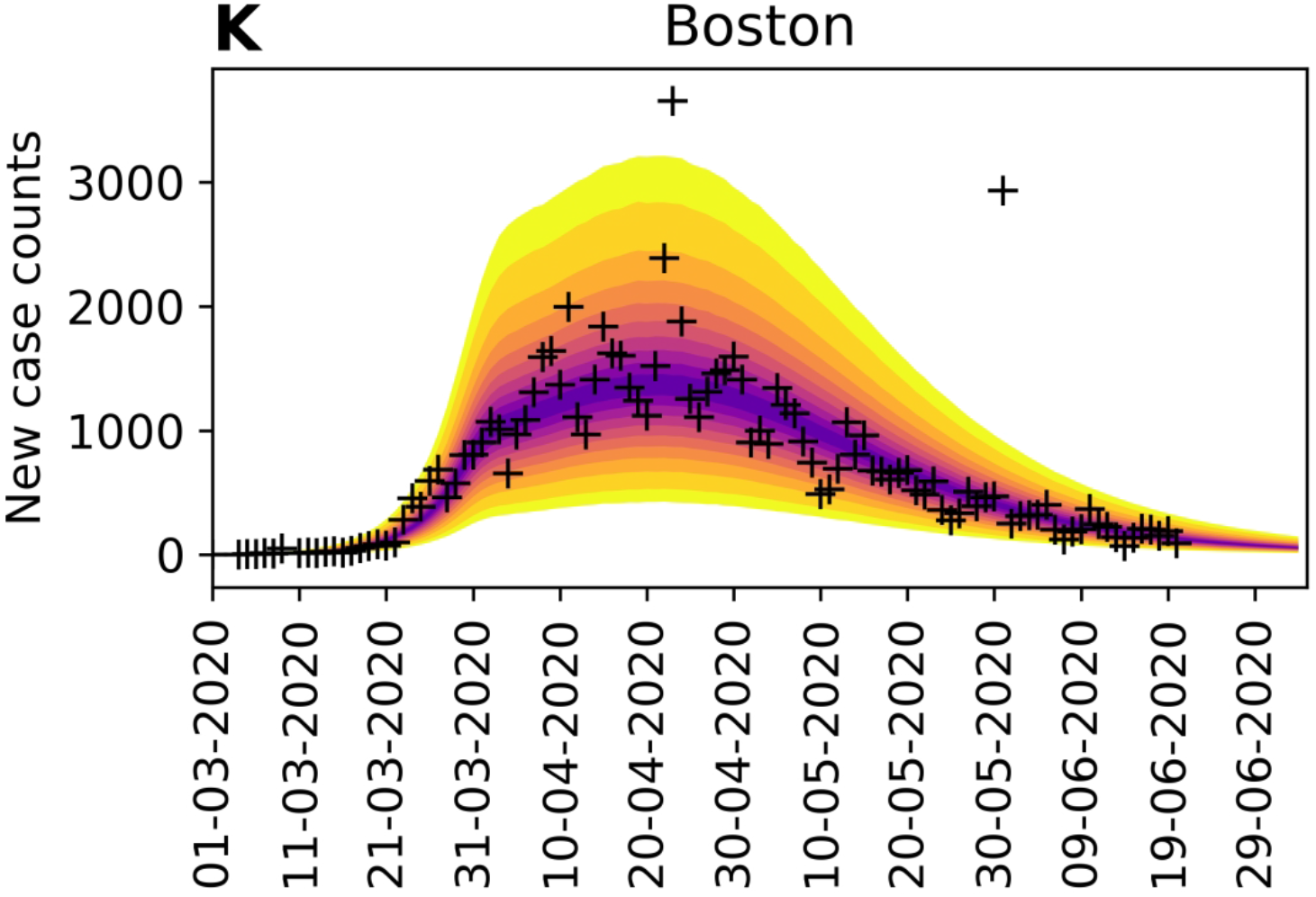
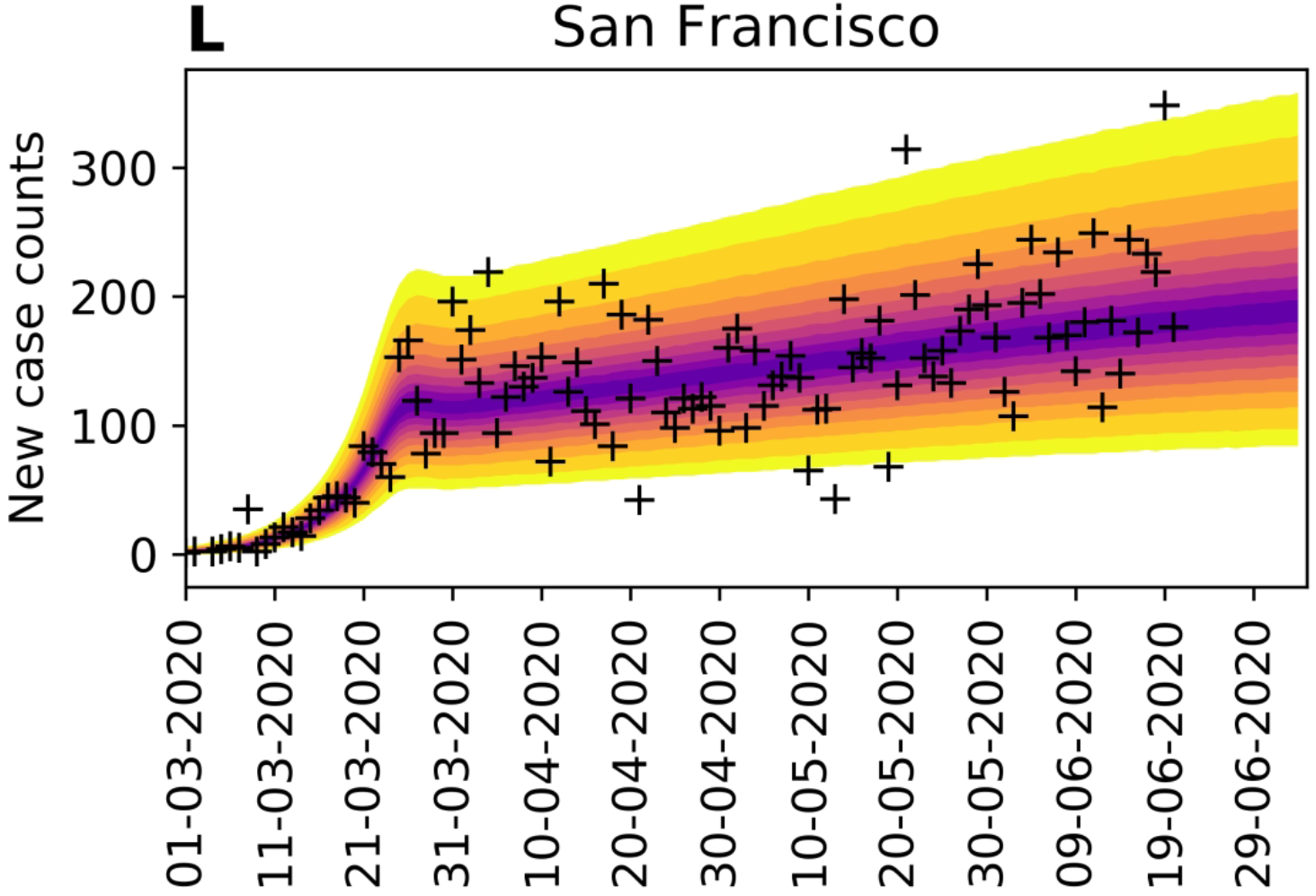
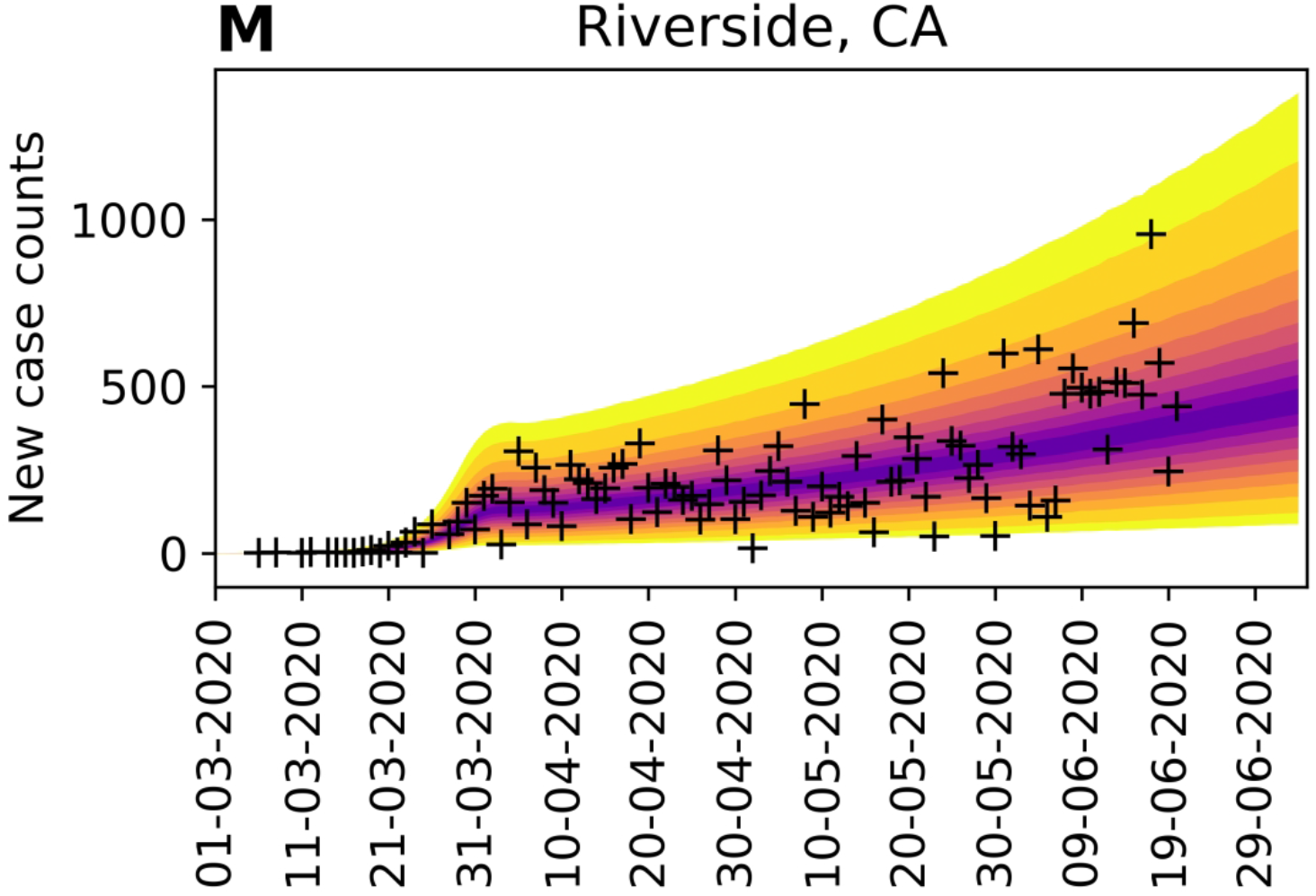
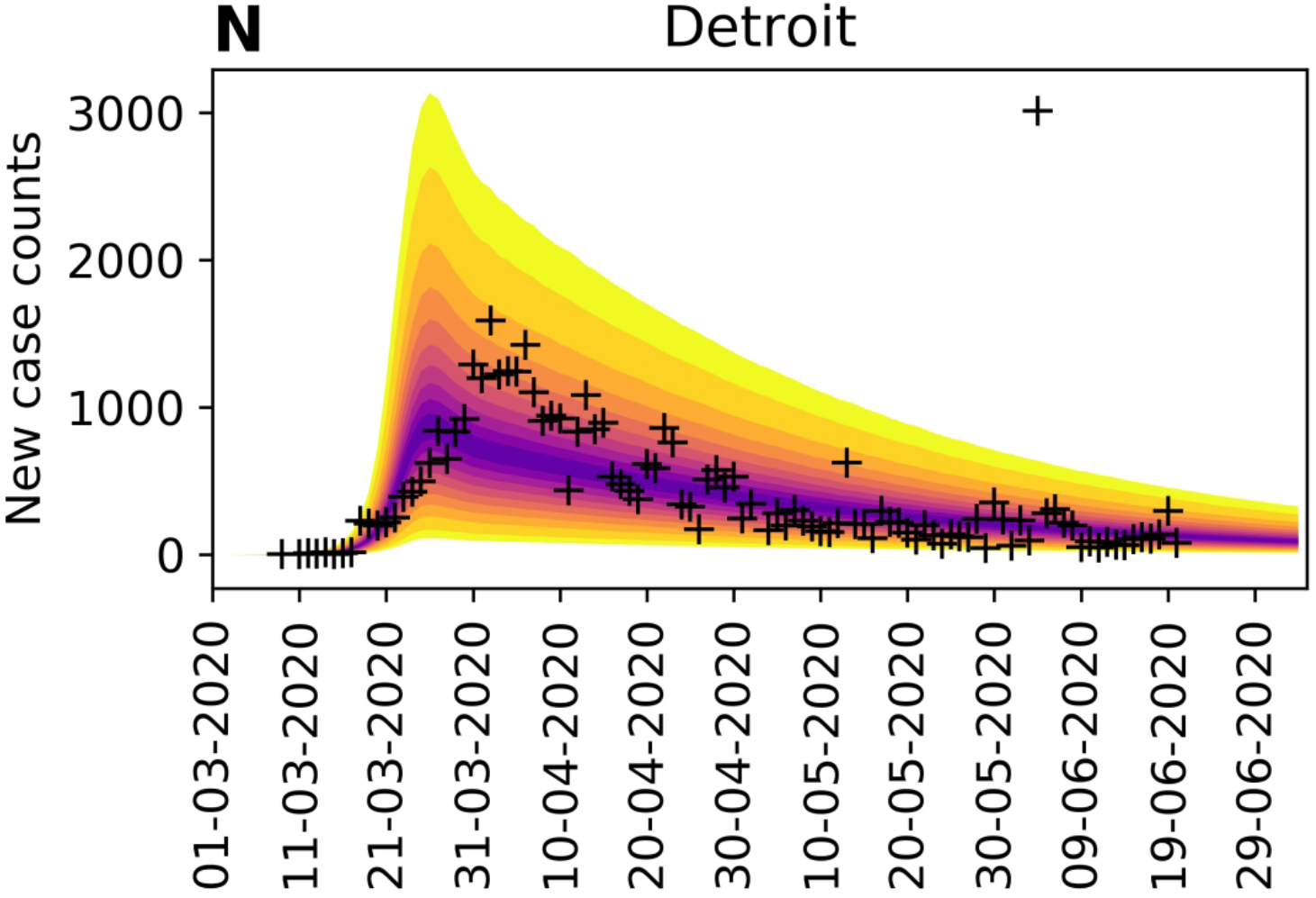
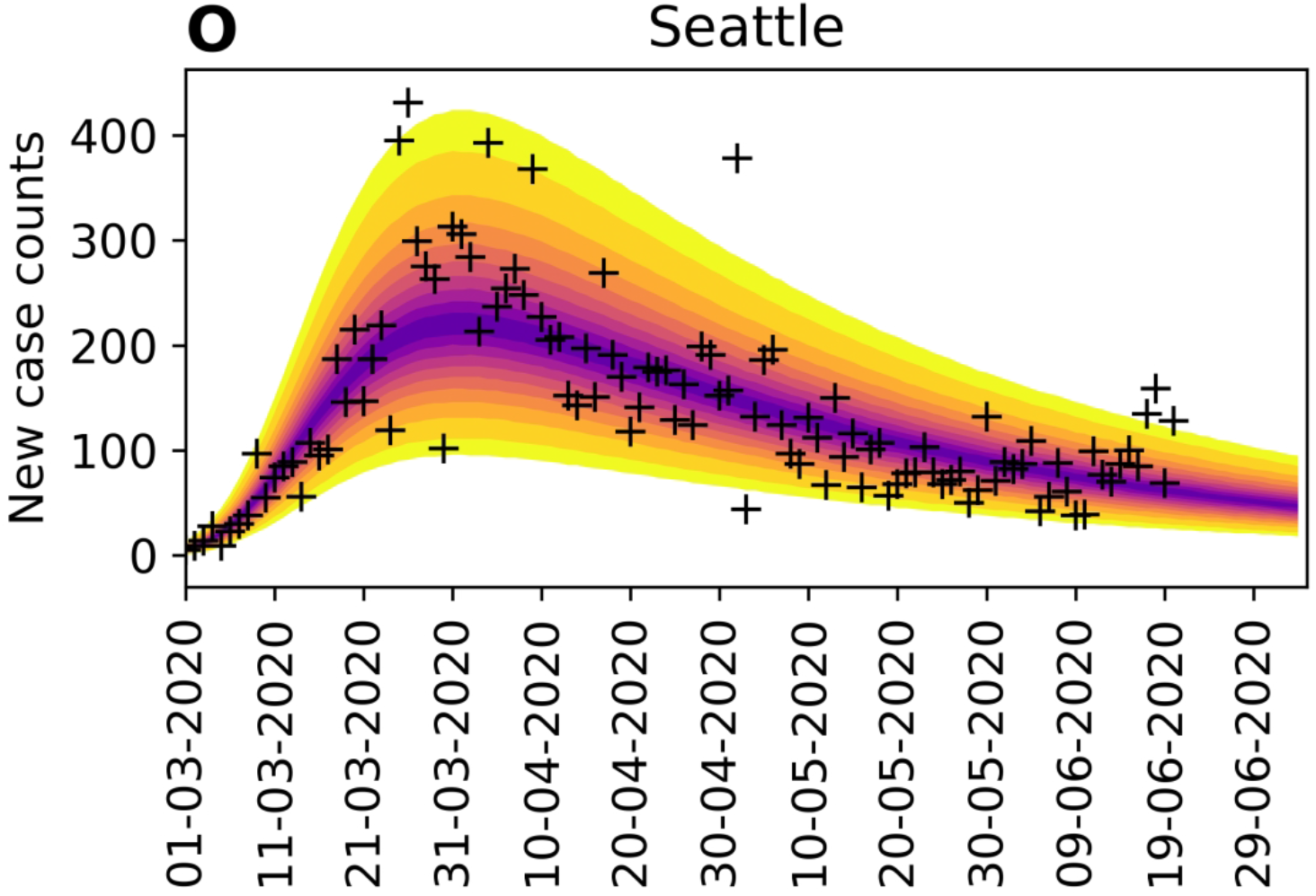
Matrix of 1- and 2-dimensional projections of the 7-dimensional posterior samples obtained for the adjustable parameters associated with the compartmental model (*n* = 0) for the New York City metropolitan statistical area (MSA) on the basis of daily reports of new confirmed coronavirus disease 2019 (COVID-19) cases from 21-January-2020 to 21-June-2020 (inclusive dates). Plots of marginal posteriors (1-dimensional projections) are shown on the diagonal from top left to bottom right. Other plots are 2-dimensional projections, which indicate how correlated pairs of parameter estimates are. Brightness indicates higher probability density. A compact bright area indicates absence of or relatively low correlation. An extended, asymmetrical bright area indicates relatively high correlation.

How does learning the region-specific adjustable parameter values of the compartmental model and a subsequent predictive inference (i.e., a forecast with UQ) improve situational awareness? In the vast majority of cases, when we forecast with UQ, the empirical new-case count for the day immediately following our inference (+1), and very often for each of several additional days, falls within the 95% credible interval of the predictive posterior. When the reported number of new cases falls outside the 95% credible interval and above the 97.5% percentile, we interpret this event, which we will call an *upward-trending rare event*, to have a probability of 0.0275 or less assuming the model is both explanatory (i.e., satisfactorily consistent with historical data) and predictive of the near future. If the model is predictive of the near future, the probability of two consecutive rare events is far smaller, less than 0.001. Thus, consecutive upward-trending rare events, which we will refer to as an *upward-trending anomaly*, can be reasonably taken as a sign that the model is not in fact predictive. Indeed, an anomaly suggests that the rate of COVID-19 transmission has increased beyond what can be explained by the model.

For New York City, anomalies are not seen, as illustrated in Figure 7, panel A. However, for Phoenix, there are several anomalies, which preceded rapid and sustained growth in the number of new cases reported per day in June (Figure 7, panel B).

**Figure 7.**
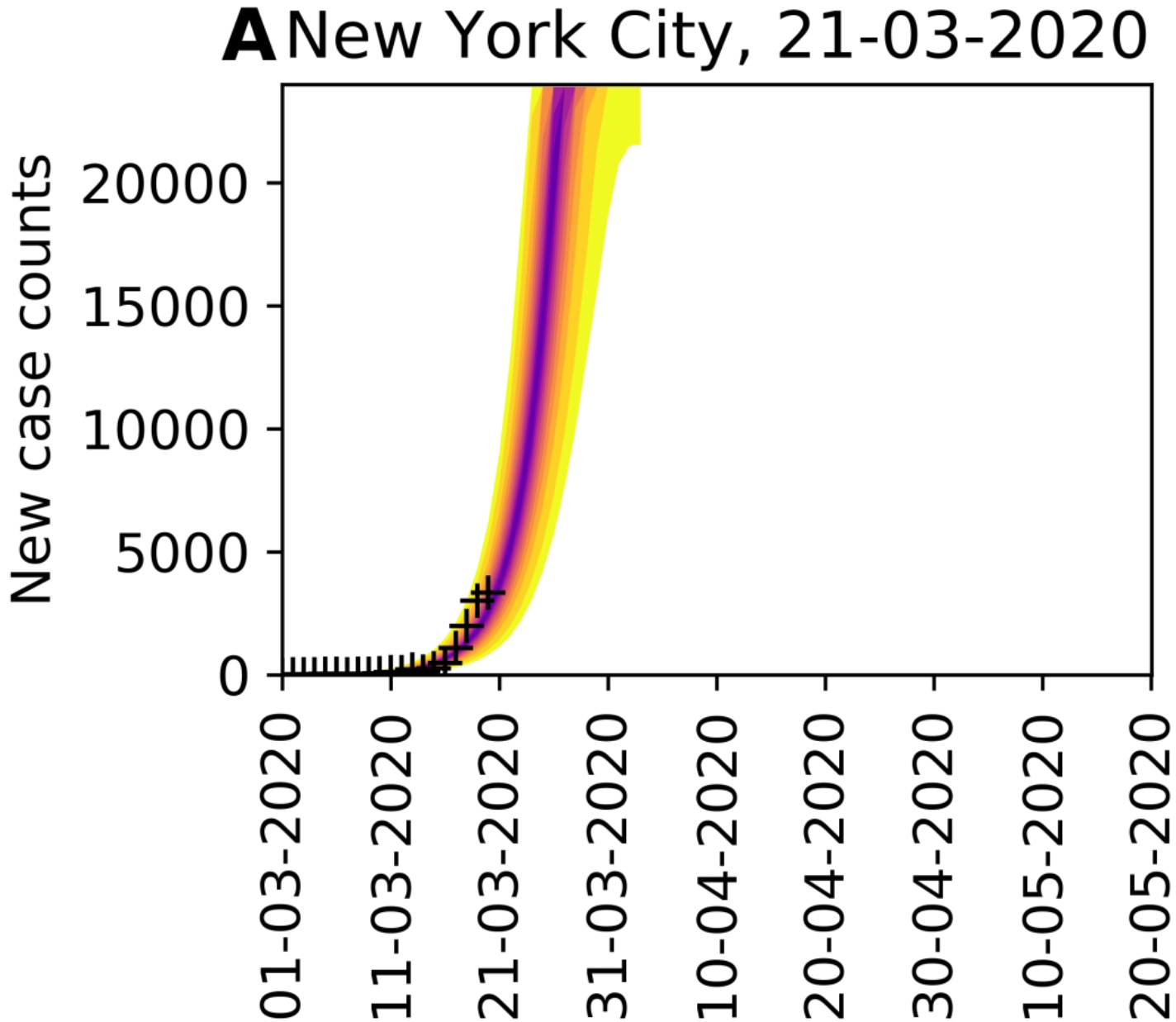
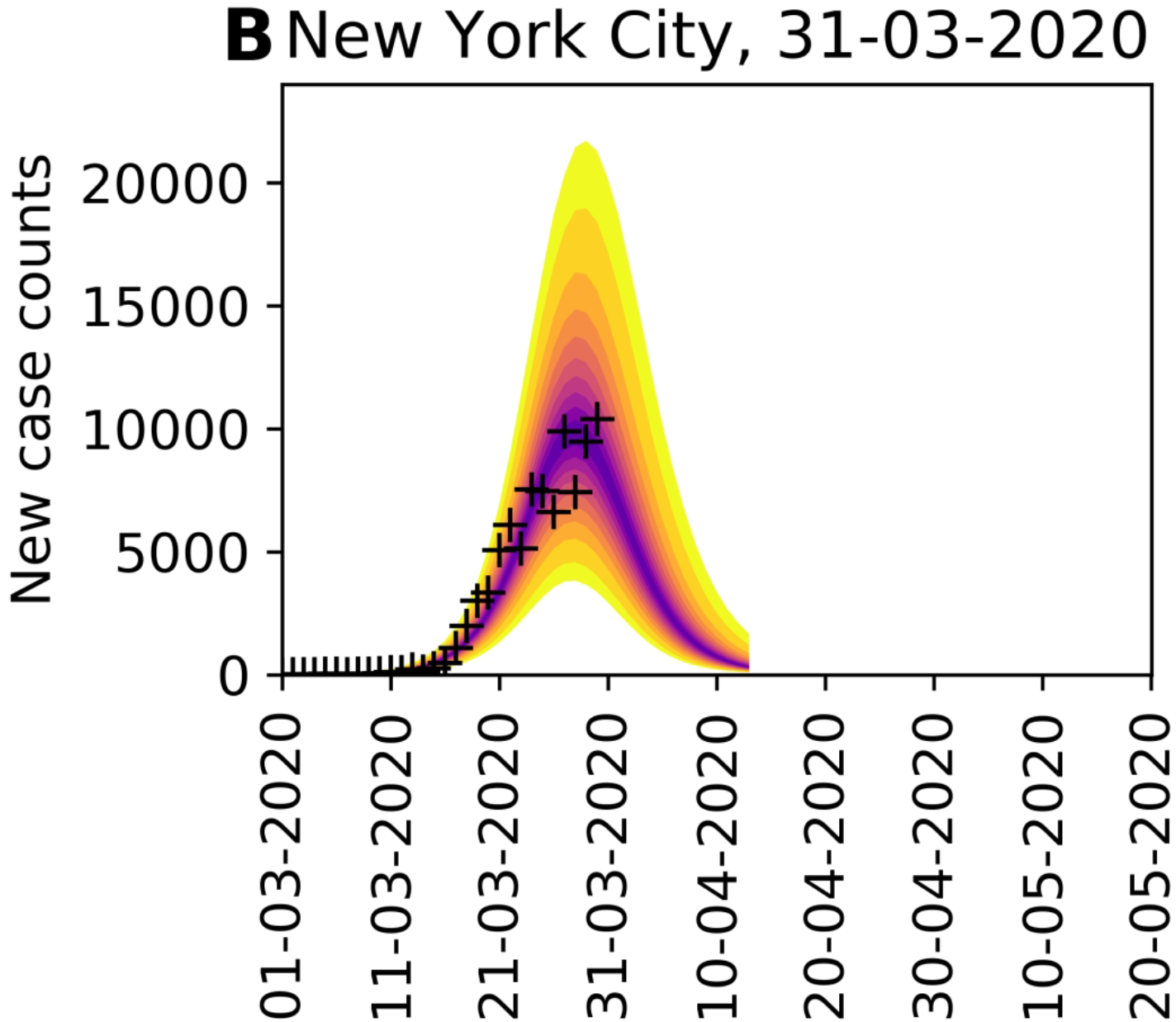
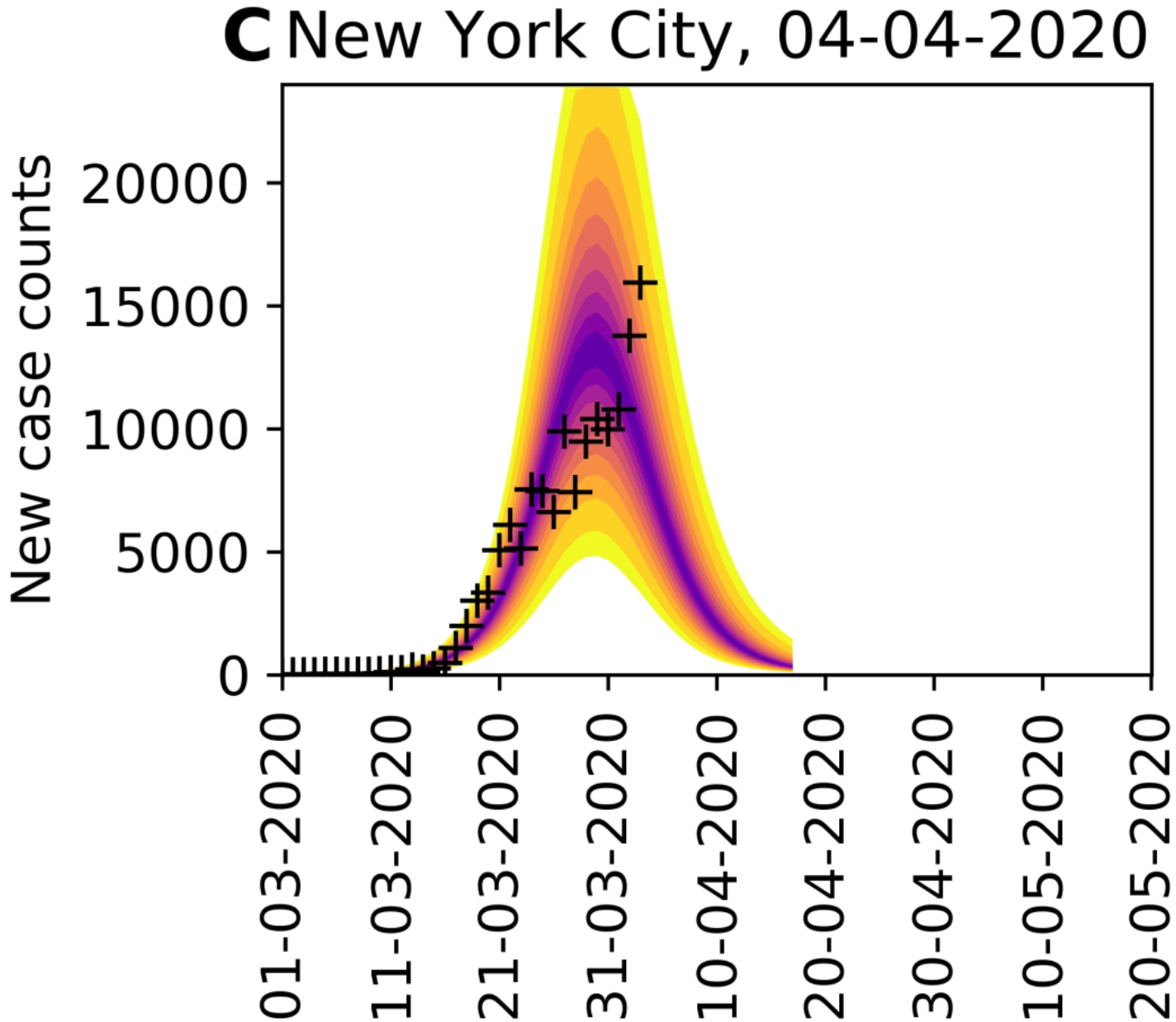
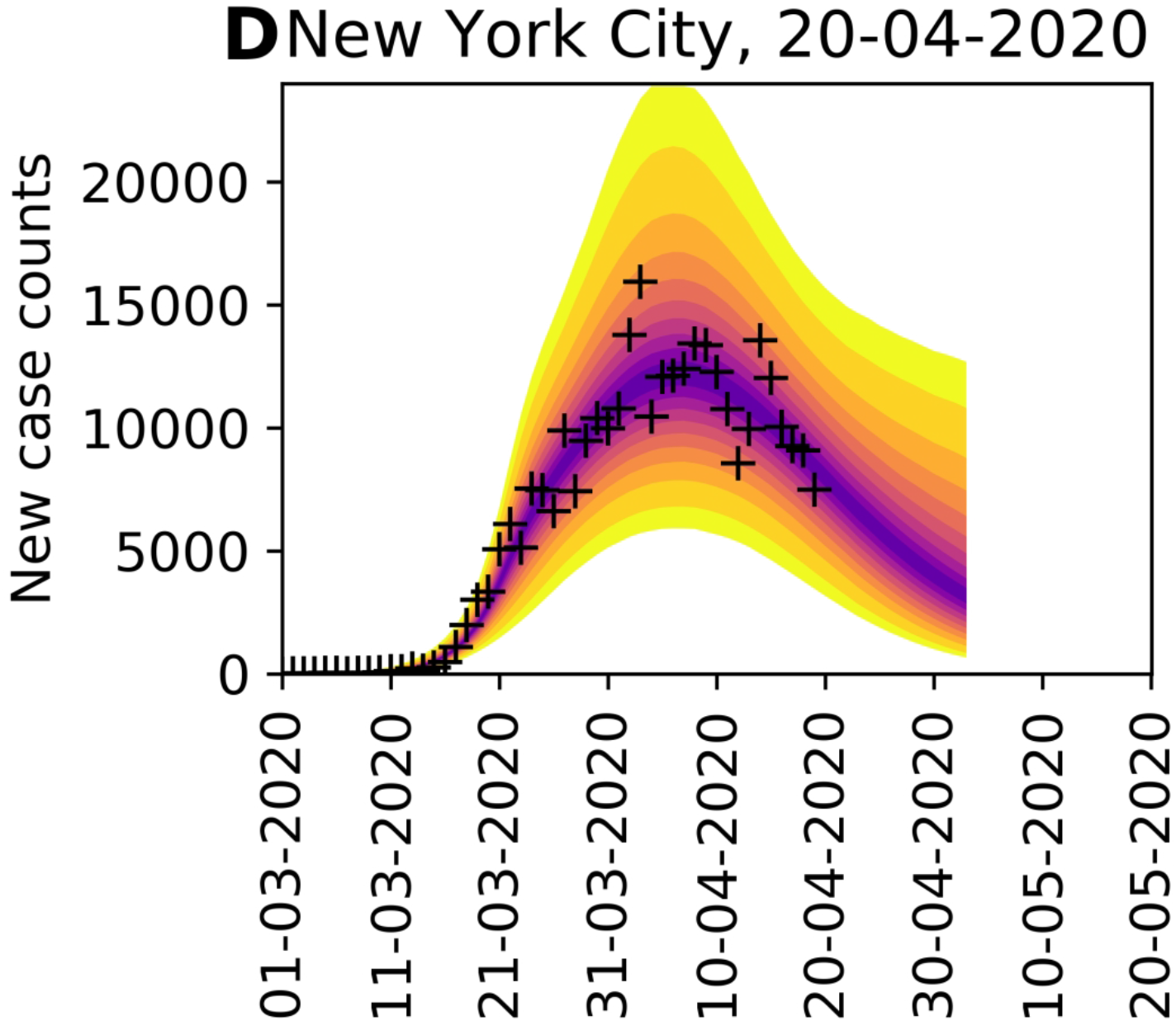
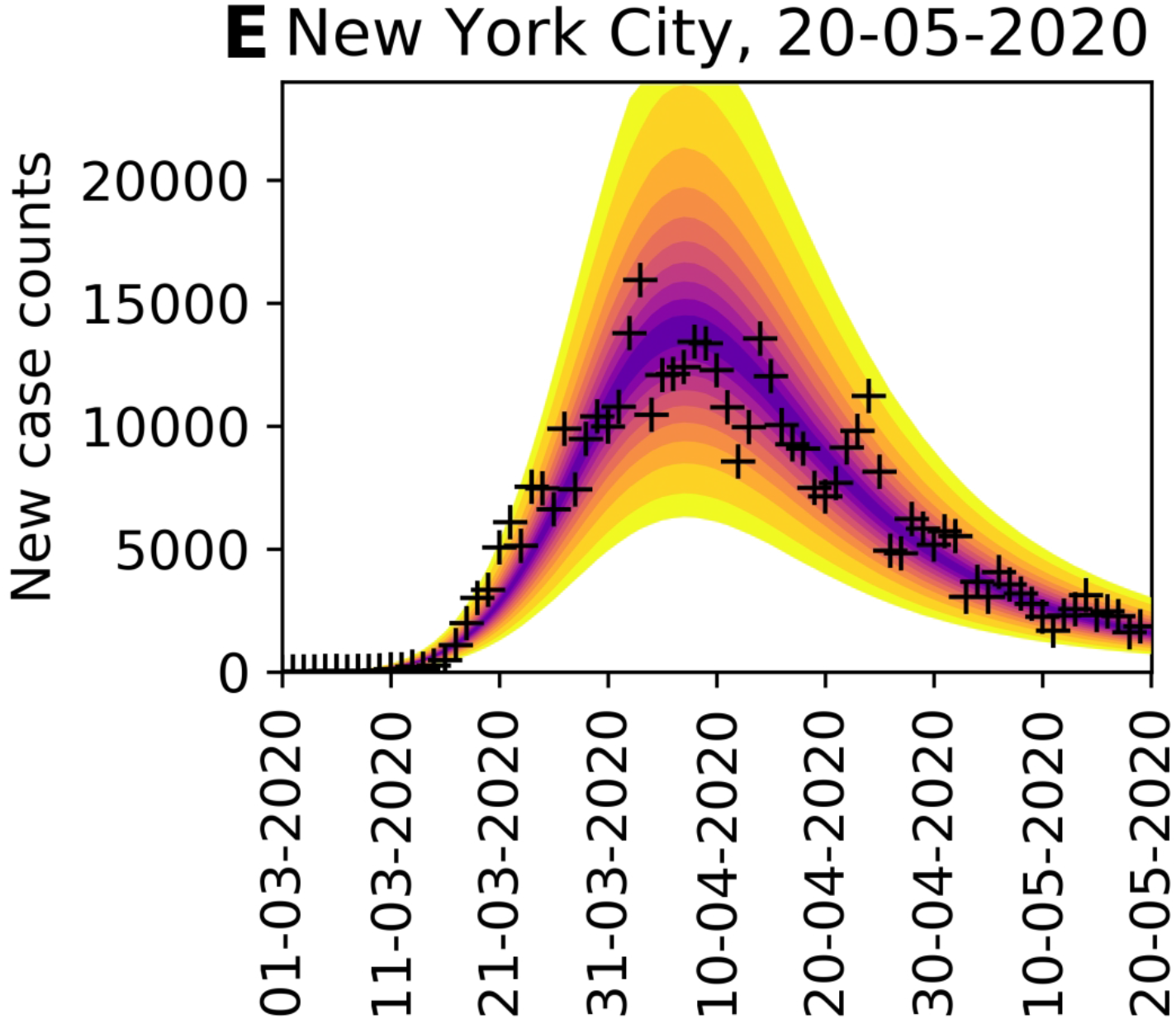
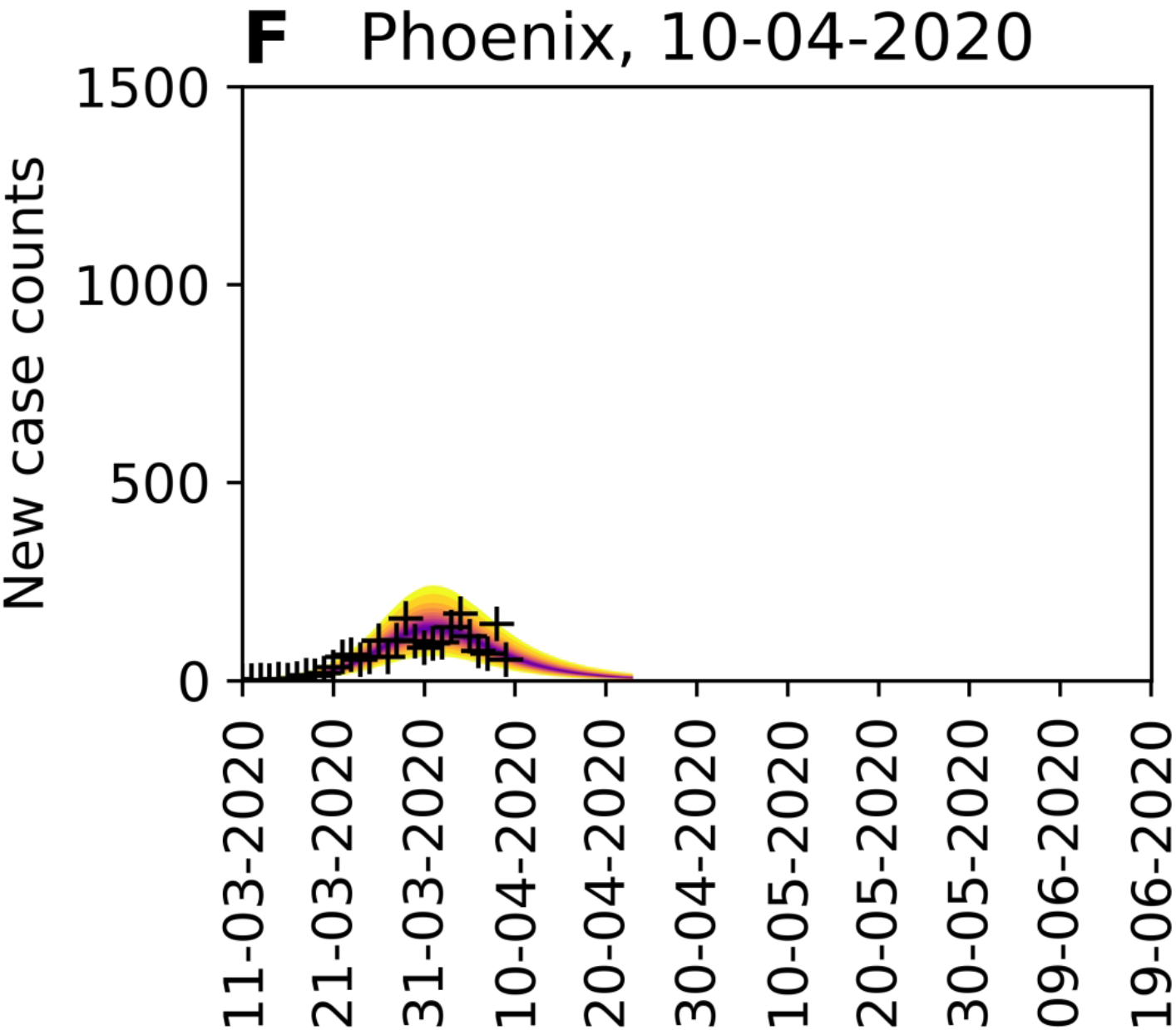
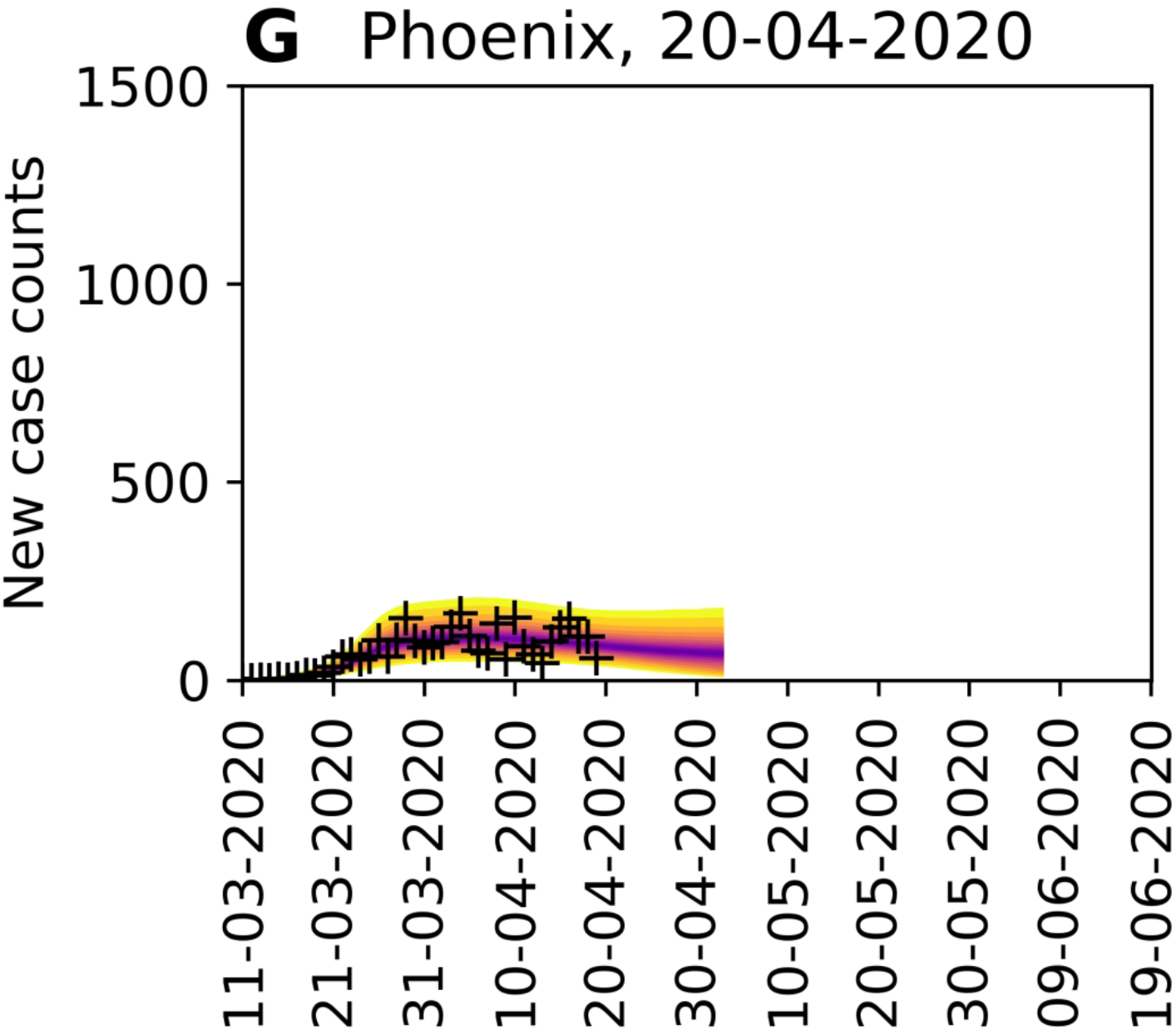
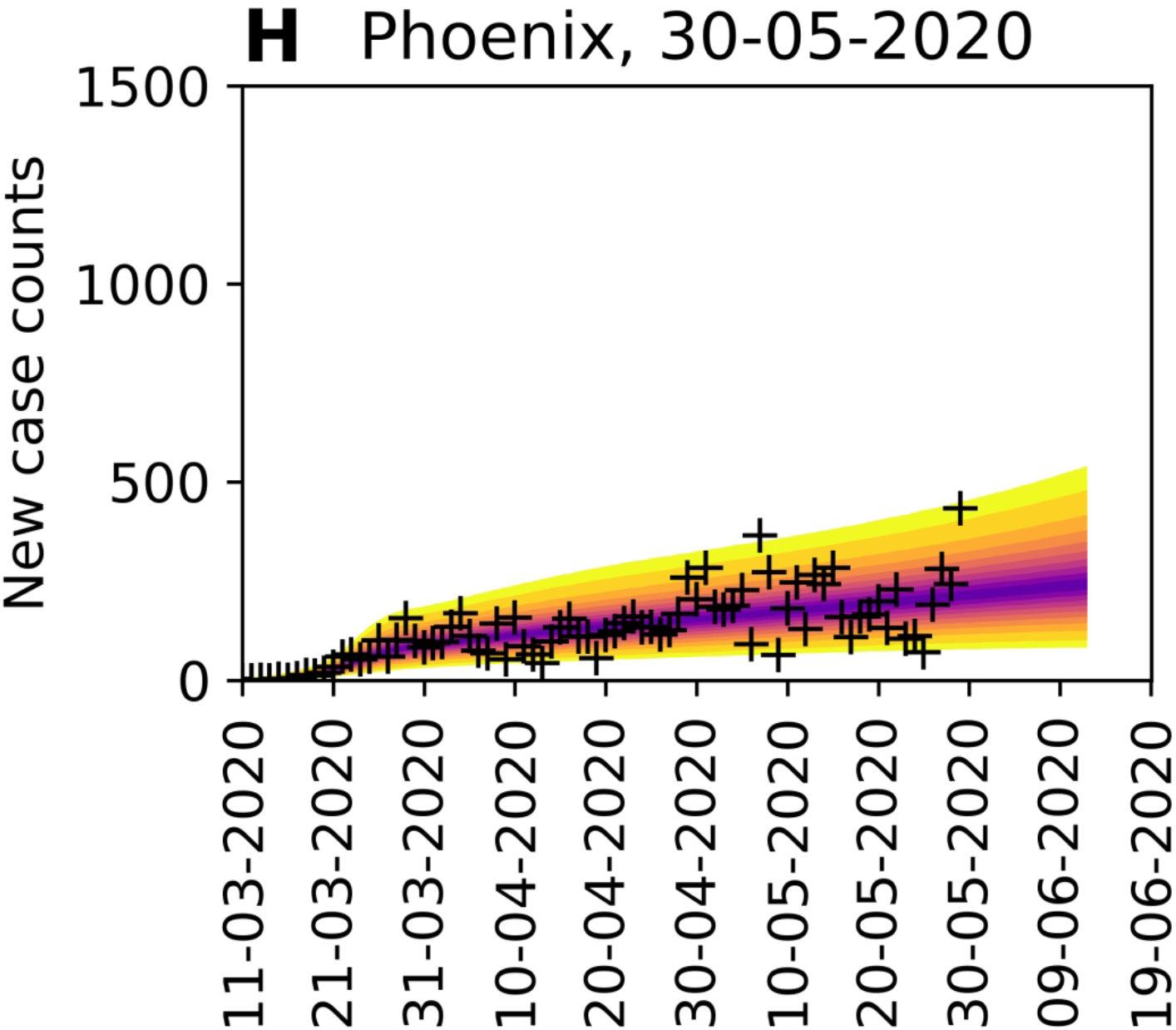
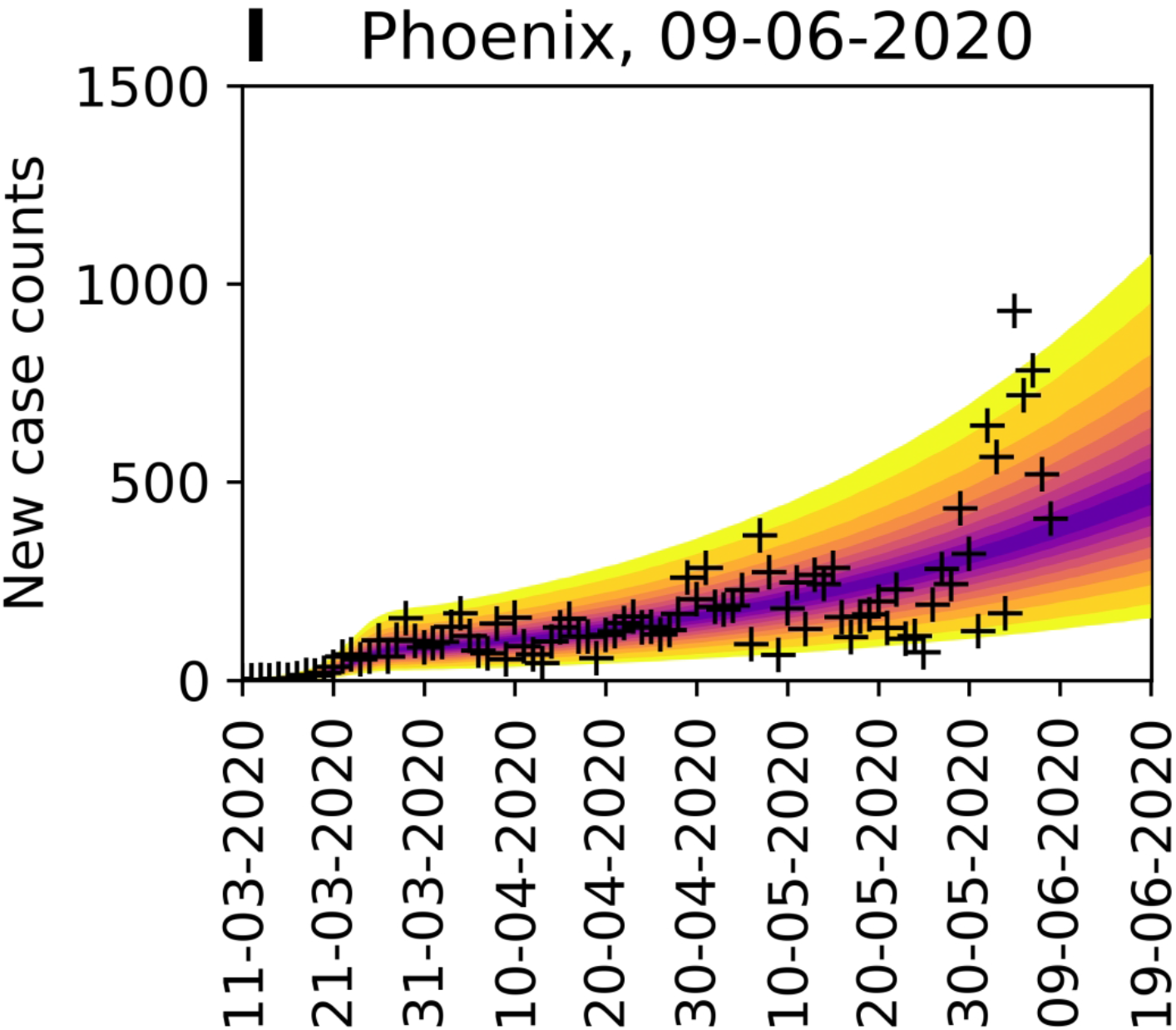
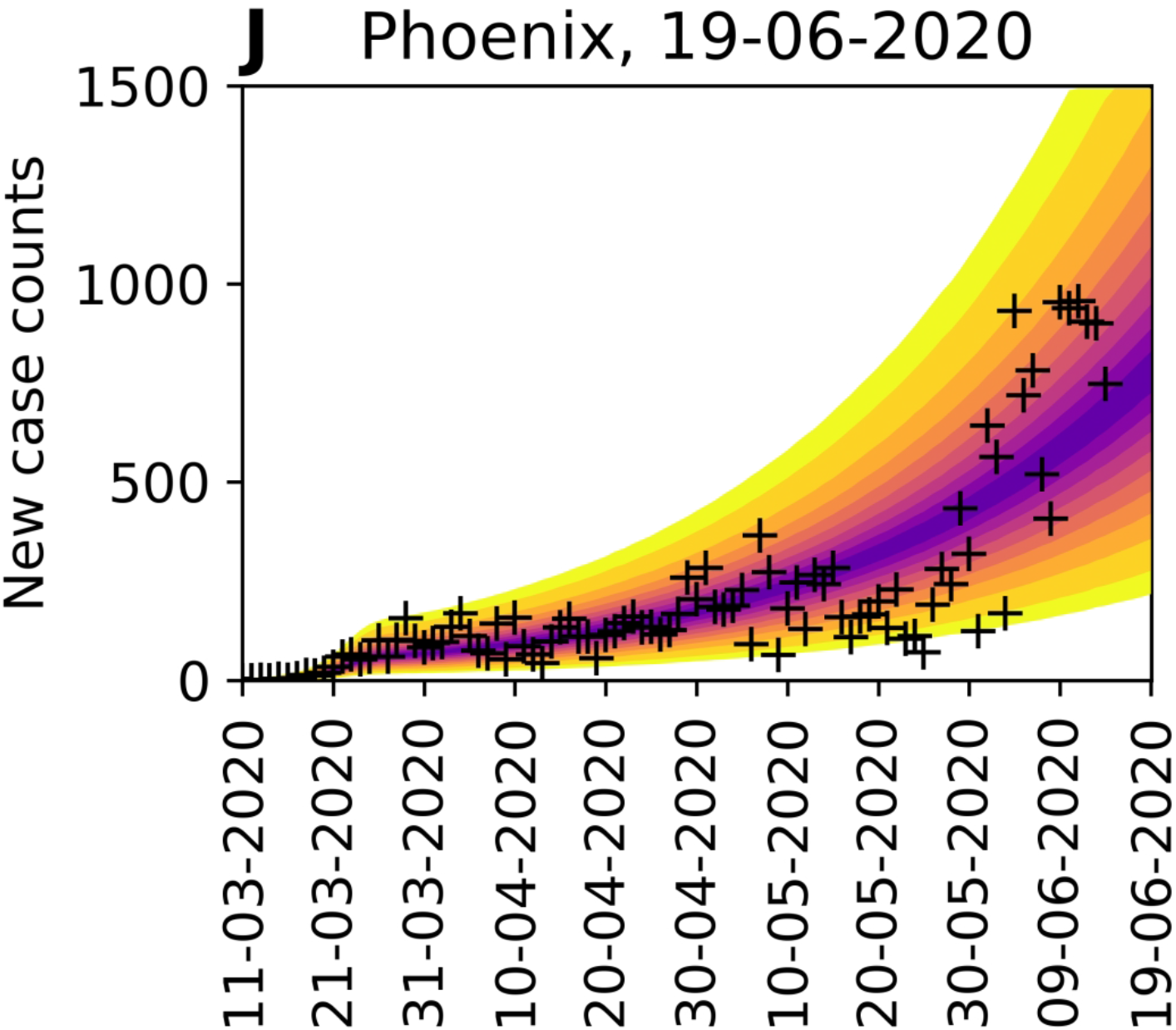
Rare events and anomalies, as defined in the main text, detected in the surveillance data available for **(A)** the New York City metropolitan statistical area (MSA) and **(B)** the Phoenix MSA. Yellow arrows mark upward-trending rare events. Red arrows mark upward-trending anomalies.

We assume that these anomalies arose from changes in behavior. Thus, to explain them, we allowed the compartmental model to account for a distinct second social-distancing period, i.e., we increased the setting for *n* from 0 to 1. With this change, the number of adjustable parameters increases from 7 to 10. One of the new parameters is *τ*_1_, the start time of the second social-distancing period. The other new parameters, *λ*_1_ and *p*_1_, replace *λ*_0_ and *p*_0_ at time *t* = *τ*_1_. As can be seen by comparing the plots in Figure 8, panels A and B, the compartmental model with two social-distancing periods (Figure 8, panel B) better explains the Phoenix data than the compartmental model with just one social-distancing period (Figure 8, panel A)^4^. Furthermore, the MAP estimate for *p*_1_ (∼0.38) is less than that for *p*_*_ (∼0.49) (cf. panels C and D, Figure 8), and the marginal posteriors for these parameters are largely non-overlapping (Figure 8, panel D). These findings suggest that the increase in COVID-19 cases in Phoenix can indeed be explained by relaxation in social distancing, which is quantified by our estimates for *p*_0_ and *p*_1_. The MAP estimate of the start time of the second period of social distancing corresponds to 24-May-2020^5^. Intriguingly, 8 of the 9 anomalies noted in Figure 8, panel B occurred after this period, with the first of these occurring on 02-June-2020.

**Figure 8.**
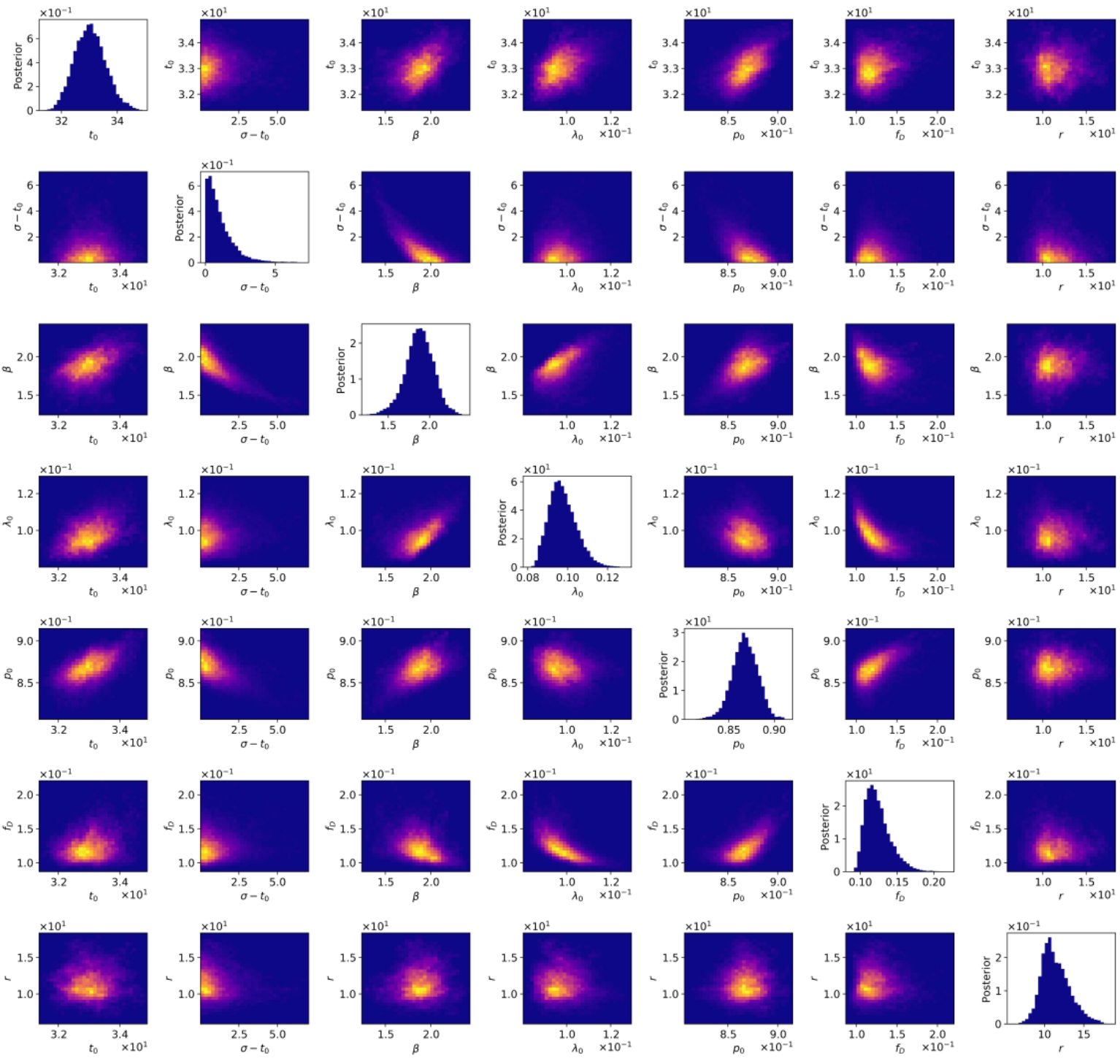
Predictions of the compartmental model **(A)** with consideration of only one period of social distancing (*n* =0) and **(B)** with consideration of an initial period of social distancing followed by a distinct period of relatively lax adherence to social-distancing practices (*n* = 1) for the Phoenix metropolitan statistical area (MSA). In panel **(C)**, the marginal posterior for the social-distancing setpoint parameter *p*_0_ inferred in the analysis of (A) is shown. In panel **(D)**, the marginal posteriors for the social-distancing parameters *p*_0_ and *p*_1_ inferred in the analysis of (B) are shown. Model selection indicates that the two-phase model is to be preferred (Appendix Table 1). In this analysis, we used data available from 21-January-2020 to 18-June-2020 (inclusive dates).

**Figure 9.**
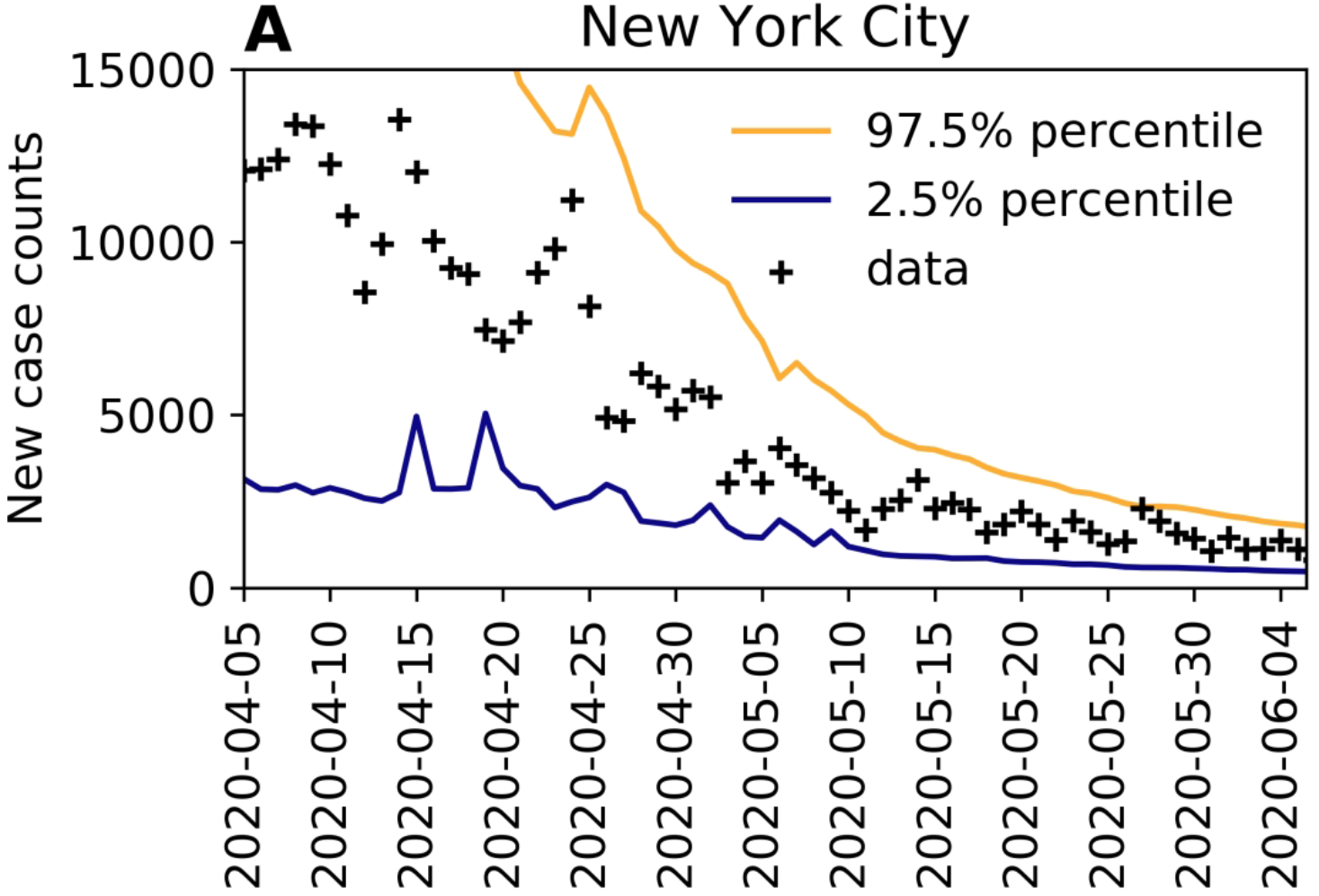
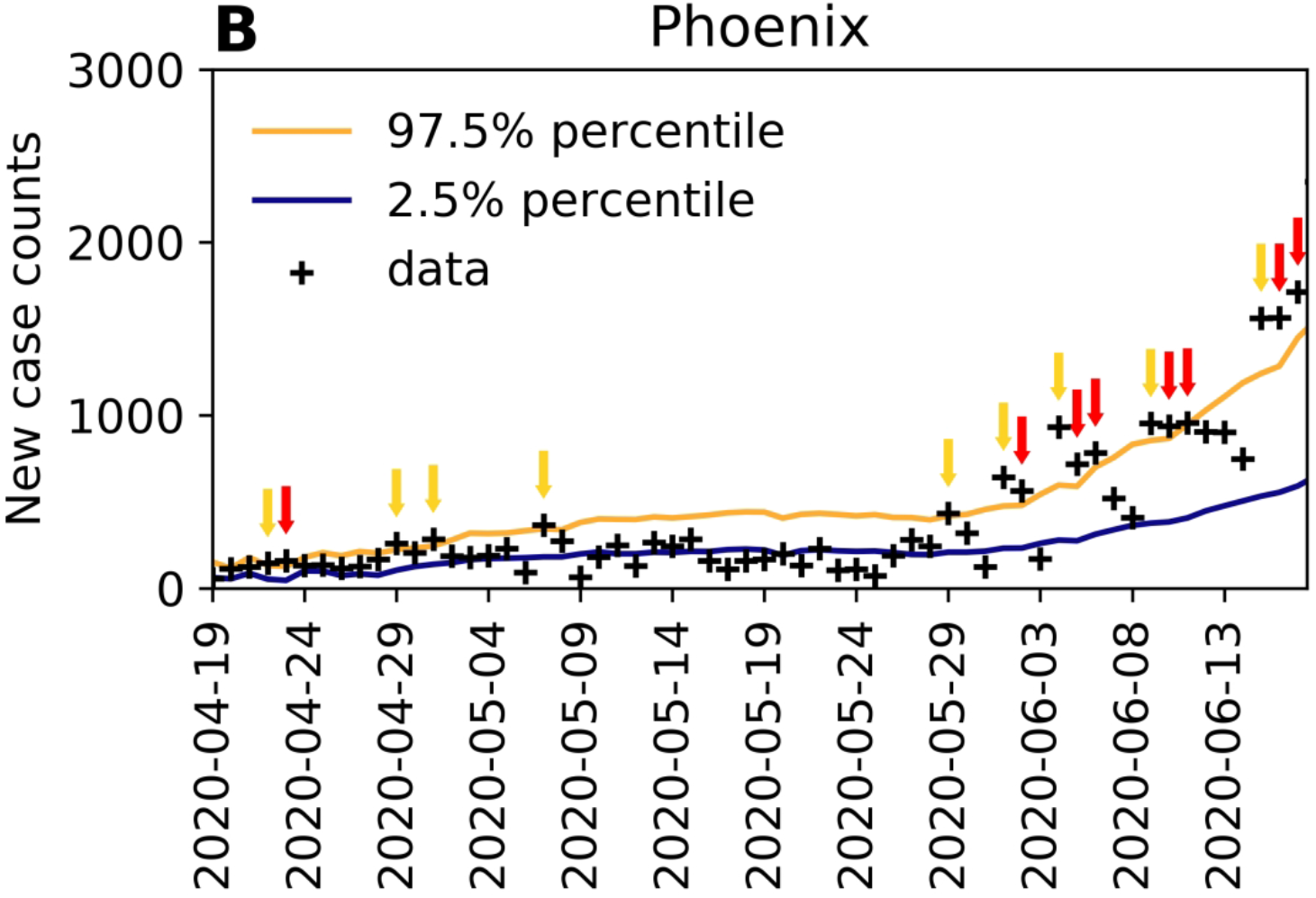
Rare events and anomalies, as defined in the main text, detected in the surveillance data available for (A) the New York City metropolitan statistical area (MSA) and (B) the Phoenix MSA. Yellow arrows mark upward-trending rare events. Red arrows mark upward-trending anomalies.

**Figure 10.**
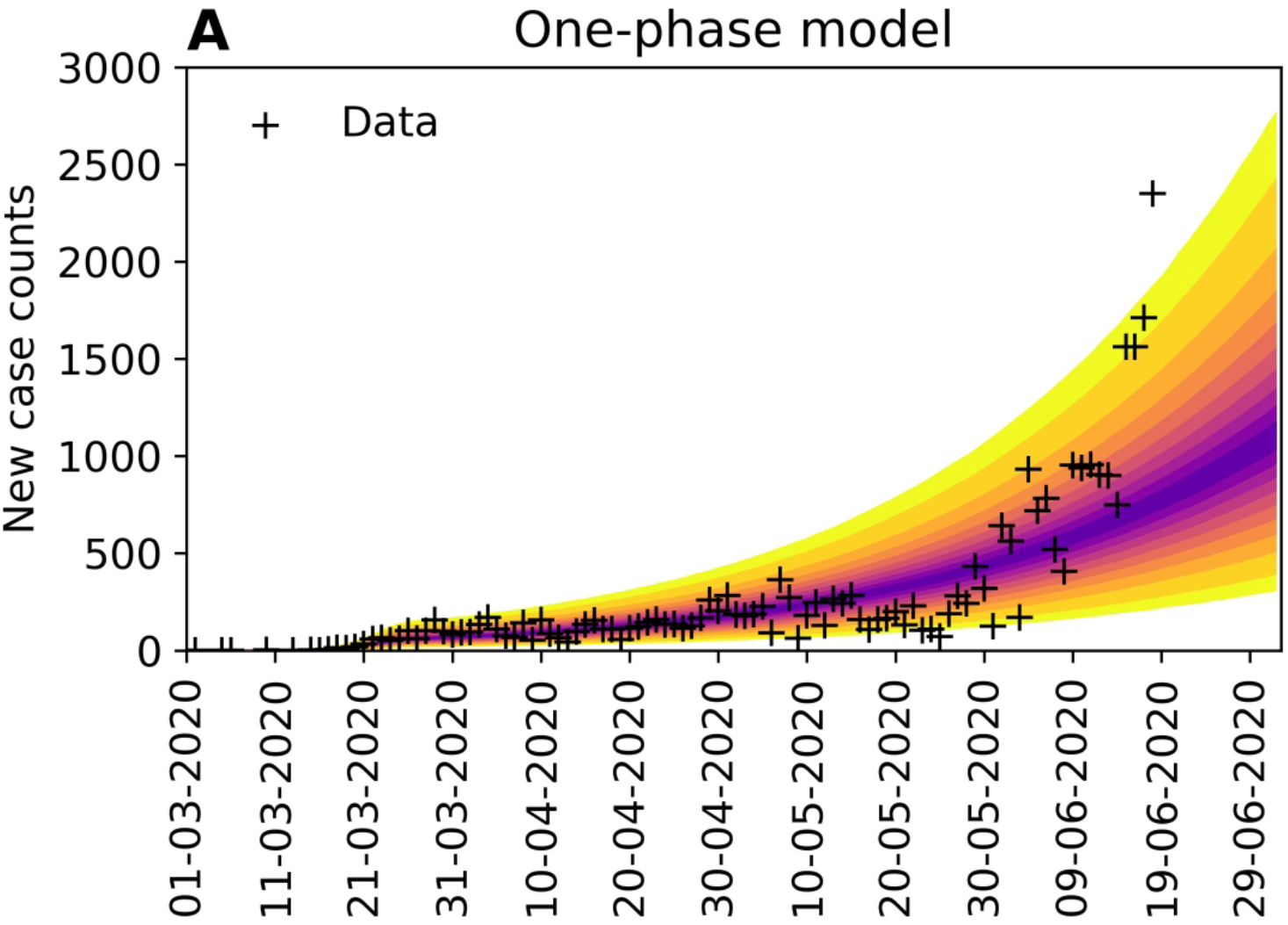
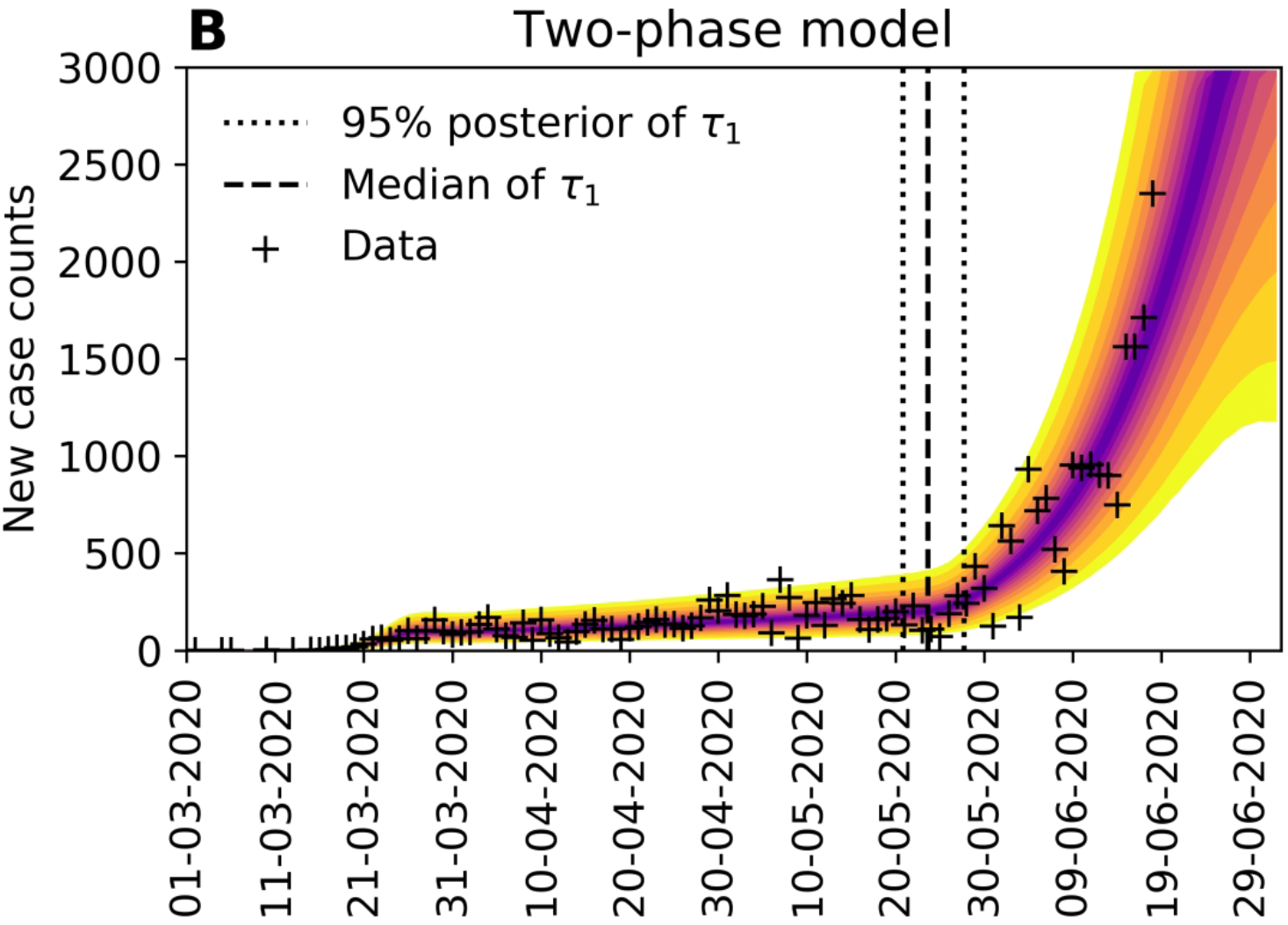
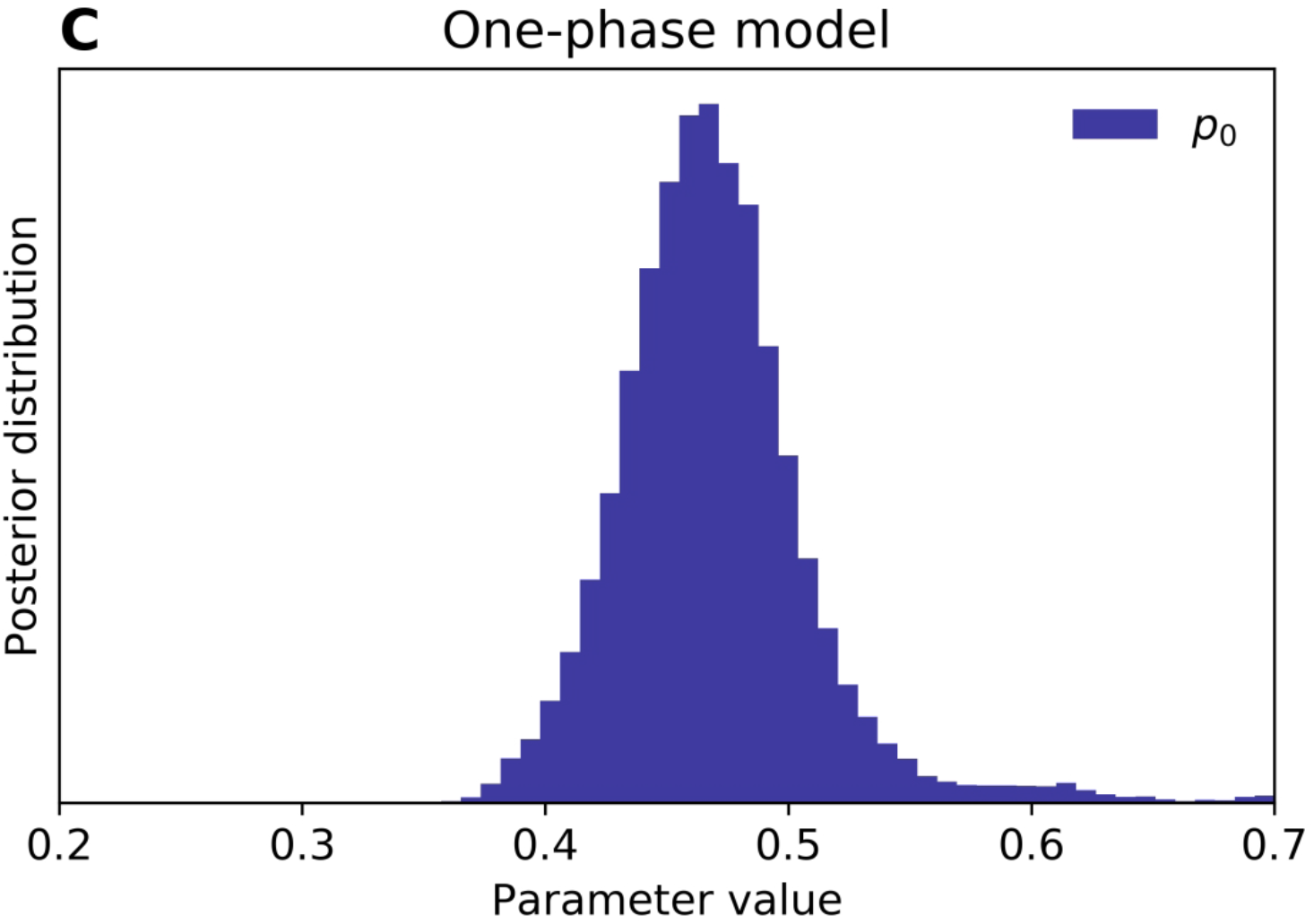
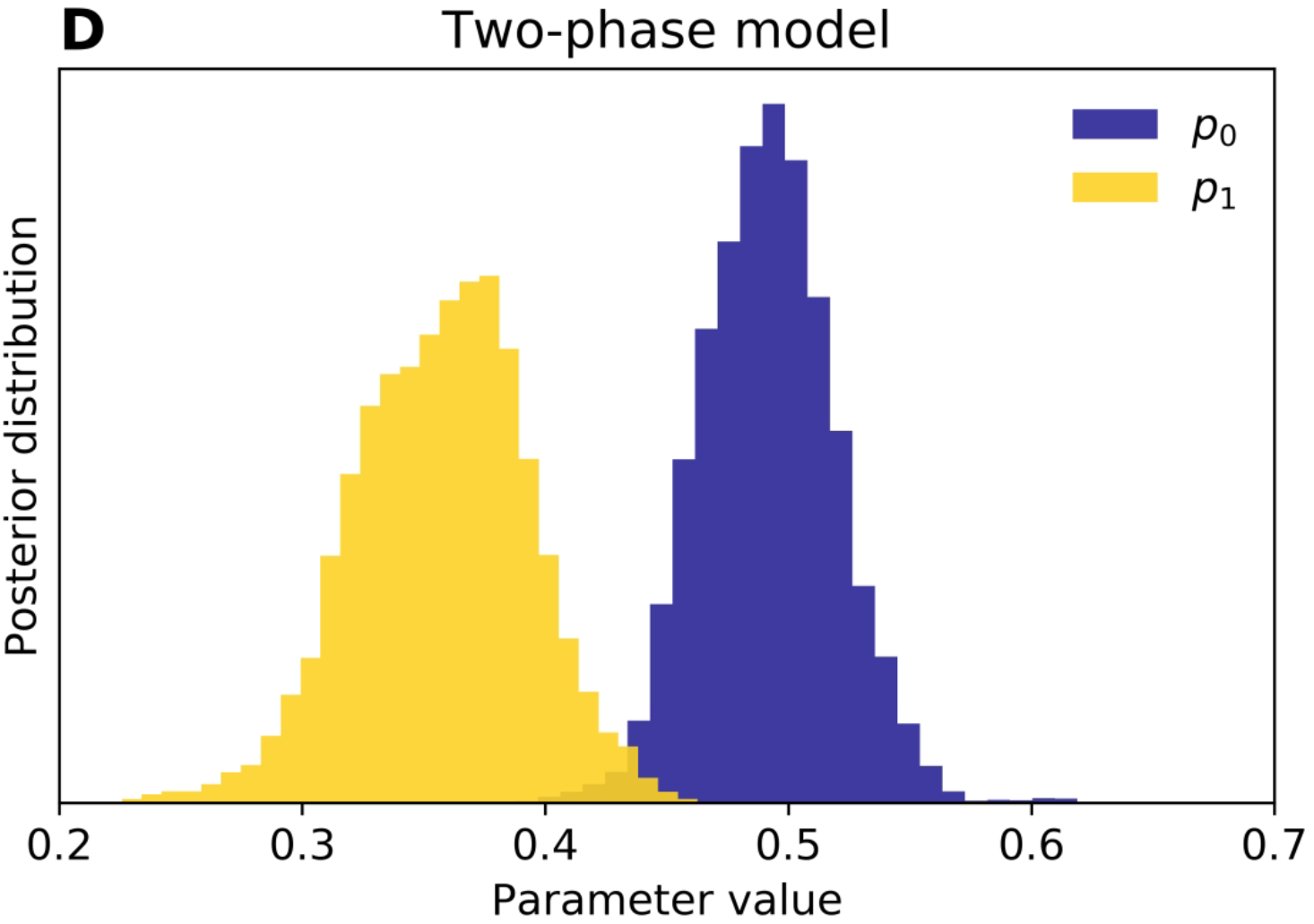
Predictions of the compartmental model (A) with consideration of only one period of social distancing (n=0) and (B) with consideration of an initial period of social distancing followed by a distinct period of relatively lax adherence to social-distancing practices (n=1) for the Phoenix metropolitan statistical area (MSA). In panel (C), the marginal posterior for the social-distancing setpoint parameter p_0 inferred in the analysis of (A) is shown. In panel (D), the marginal posteriors for the social-distancing parameters p_0 and p_1 inferred in the analysis of (B) are shown. Model selection indicates that the two-phase model is to be preferred (Appendix Table 1). In this analysis, we used data available from 21-January-2020 to 18-June-2020 (inclusive dates).

We considered the hypothesis that a one-time event generating 1000’s of new infections, such as a mass gathering, might trigger a new upward trend in COVID-19 transmission. Simulations for New York City and Phoenix do not support this hypothesis (Appendix Figure 2). In each of these simulations, at a specified time, we moved a specified number of individuals from the *S*_*M*_ (mixing susceptible) population into the *E*_1_ (exposed) population and then simulated forward in time. Each perturbation increased disease burden, but the perturbation had minimal effect on the slope of the trajectory of new case detection.

Besides Phoenix, four other MSAs have contemporaneous trends explainable by relaxation of social distancing (Appendix Figure 3 and Appendix Table 1). Upward-trending anomalies were detected for these MSAs (Appendix Figure 4, panels A–D), but not for three of four other MSAs having epidemic curves consistent with sustained social distancing (Appendix Figure 4, panels E–H). Daily predictions for the MSAs considered in Appendix Figure 4 are shown in Videos 3–10.

An assessment of the overall prediction accuracy of the region-specific compartmental models is provided in Appendix Figure 5.

## Discussion

Daily online learning of model parameter values from real-time surveillance data is feasible for mathematical models for COVID-19 transmission. Furthermore, predictive inference of the daily number of new cases reported is feasible for the regional COVID-19 epidemics occurring in multiple US metropolitan areas. Our model-based analyses are ongoing and daily forecasts are being disseminated *(23)*. However, it should be noted that inferences are computationally expensive and the cost increases as new data become available. Thus, at some point, daily inferences using our methodology may become impractical.

We have suggested how our predictive inferences can be used to identify harbingers of future growth in COVID-19 transmission rate. Two consecutive upward-trending rare events (i.e., instances where the number of new cases reported is above the upper limit of the 95% credible interval of the predictive posterior) seem to indicate potential for increased transmission in the future. This signal of future growth is perhaps especially strong when anomalies are accompanied by increasing prediction uncertainty, as is the case for Phoenix (Figure 7, panel B).

We find that the June increase in rate of transmission of COVID-19 in the Phoenix metropolitan area can be explained by a reduction in the percentage of the population adhering to effective social-distancing practices^6^ (Figure 8, panel D), from ∼49% to ∼38%. Relaxation is inferred to have begun around 24-May-2020 (Figure 8, panel B). Contemporaneous upward trends in the rate of COVID-19 transmission in the Houston, Miami, San Francisco, and Seattle metropolitan areas can also be explained by relaxation of social distancing (Appendix Figure 3 and Appendix Table 1). These findings are qualitatively consistent with earlier studies indicating that social distancing is effective at slowing the transmission of COVID-19 *(7, 8)*, and encouragingly, they suggest that the future course of the pandemic is controllable, especially with accurate recognition of when stronger nonpharmaceutical interventions are needed to slow COVID-19 transmission.

We caution that our work has several limitations. One is that trend detection is data-driven, which means that a new trend cannot be detected until enough evidence of it has accumulated. The data we are using are reports of new cases, which reflect transmission dynamics of the past vs. current transmission dynamics. Other types of surveillance data, such as assays of viral RNA in wastewater samples, may permit improved situational awareness. Another limitation is that our inferences are based on a mathematical model associated with considerable structure and fixed parameter uncertainties as well as simplifications. Among the simplifications is the replacement of certain time-varying parameters, such as that characterizing testing capacity, with constants, which are assumed to provide an adequate time-averaged characterization. The model form is that of a deterministic compartmental model. A stochastic version of the model may be more appropriate if conditions change (from the current situation of high disease prevalence to low prevalence). Although the model is capable of reproducing historical data and making accurate short-term forecasts, its structure and fixed parameters are subject to revision as we learn more about COVID-19. In the future, results from serological studies and estimates of excess deaths should allow model improvements.

## Data Availability

The data for daily inference was extracted from the New York Times repository https://github.com/nytimes/covid-19-data.
The results of the daily inference will be deposited in our GitHub repository: https://github.com/lanl/COVID-19-Predictions

https://github.com/lanl/COVID-19-Predictions

## Acknowledgments

Y.T.L. was supported by the Laboratory Directed Research and Development Program at Los Alamos National Laboratory (Project XX01); this support enabled early feasibility studies. Y.T.L., C.S., J.R., G.T., S.C., and W.S.H. were supported by the US Department of Energy (DOE) Office of Science through the National Virtual Biotechnology Laboratory, a consortium of national laboratories (Argonne, Los Alamos, Oak Ridge, and Sandia) focused on responding to COVID-19, with funding provided by the Coronavirus CARES Act. J.N., E.M., and R.G.P. were supported by a grant from the National Institute of General Medical Sciences of the National Institutes of Health (R01GM111510). A.M. was supported by the 2020 Mathematical Sciences Graduate Internship program, which is sponsored by the Division of Mathematical Sciences of the National Science Foundation. Computational resources used in this study included the following: the Darwin cluster at Los Alamos National Laboratory (LANL), which is supported by the Computational Systems and Software Environment subprogram of the Advanced Simulation and Computing program at LANL, which is funded by National Nuclear Security Administration of the DOE and Northern Arizona University’s Monsoon computer cluster, which is funded by Arizona’s Technology and Research Initiative Fund.

## Disclaimers

None.

## Author Bio

Dr. Lin works as a Scientist in the Information Sciences Group of the Computer, Computational, and Statistical Sciences Division at Los Alamos National Laboratory. His primary research interest is development and application of advanced data-science methods in modeling of biological systems.

---

**Appendix Table 1.** Strength-of-evidence comparison of compartmental models accounting for an initial social-distancing period only (*n* = 0) and for an initial social-distancing period followed by a distinct second-phase social distancing period (*n* = 1).

| MSA | $\Delta AIC^*$ | $\Delta BIC^*$ | $p_0^{n=0}$ (95%)** | $p_0^{n=1}$ (95%)** | $p_1^{n=1}$ (95%)** |
| --- | --- | --- | --- | --- | --- |
| New York City | 17 | 8.6 | 0.88 (0.85–0.90) | 0.87 (0.80–0.89) | 0.36 (0.11–0.83) |
| Los Angeles | −6.5 | −15 | 0.45 (0.38–0.45) | 0.47 (0.42–0.80) | 0.38 (0.33–0.97) |
| Chicago | 18 | 9.5 | 0.57 (0.46–0.61) | 0.52 (0.46–0.75) | 0.25 (0.03–0.68) |
| Dallas | 18 | 9.4 | 0.52 (0.41–0.52) | 0.59 (0.49–0.77) | 0.41 (0.33–0.60) |
| Houston | 50 | 42 | 0.39 (0.34–0.45) | 0.49 (0.39–0.79) | 0.30 (0.20–0.56) |
| Washington | 1.0 | −7.5 | 0.39 (0.30–0.47) | 0.77 (0.71–0.80) | 0.68 (0.63–0.76) |
| Miami | 75 | 67 | 0.51 (0.46–0.57) | 0.92 (0.81–0.97) | 0.69 (0.61–0.80) |
| Philadelphia | 12 | 3.7 | 0.65 (0.57–0.69) | 0.55 (0.49–0.81) | 0.22 (0.03–0.69) |
| Atlanta | 9.9 | 1.5 | 0.54 (0.41–0.52) | 0.58 (0.44–0.78) | 0.29 (0.06–0.63) |
| Phoenix | 66 | 58 | 0.43 (0.37–0.49) | 0.55 (0.43–0.73) | 0.34 (0.26–0.54) |
| Boston | −31 | −39 | 0.36 (0.29–0.37) | 0.80 (0.69–0.85) | 0.18 (0.06–0.97) |
| San Francisco | 20 | 12 | 0.32 (0.29–0.35) | 0.36 (0.34–0.74) | 0.17 (0.07–0.63) |
| Riverside | 3.8 | −4.7 | 0.41 (0.36–0.46) | 0.43 (0.38–0.74) | 0.34 (0.03–0.48) |
| Detroit | 5.9 | -2.6 | 0.75 (0.60–0.78) | 0.80 (0.64–0.92) | 0.93 (0.14–0.97) |
| Seattle | 55 | 46 | 0.87 (0.75–0.90) | 0.82 (0.76–0.85) | 0.59 (0.48–0.68) |

\* $\Delta AIC \equiv AIC^{n=0} - AIC^{n=1}$ and $\Delta BIC \equiv BIC^{n=0} - BIC^{n=1}$ , where $AIC^{n=0}$ and $AIC^{n=1}$ are the Aikake information criterion (AIC) (24) values calculated for the $n = 0$ and $n = 1$ versions of the compartmental model and, similarly, $BIC^{n=0}$ and $BIC^{n=1}$ are the Bayesian information criterion (BIC) (24) values calculated for the $n = 0$ and $n = 1$ versions of the compartmental model. There are five MSAs for which $\Delta AIC$ and $\Delta BIC$ are both greater than 10: Houston, Miami, Phoenix, San Francisco, and Seattle. There is one MSA for which $\Delta AIC$ and $\Delta BIC$ are both less than $-10$ : Boston. \*\*The first entry in each row of this column is the maximum a posteriori (MAP) estimate. The next entry, a pair of numbers within parentheses, indicates the 95% credible interval. In this analysis, we used data available from 21-January-2020 to 26-June-2020 (inclusive dates).

**Appendix Table 2.** Parameters of the curve-fitting model (*N, t*_0_, *k, θ, μ*_*LN*_, and *σ* _*LN*_) and the associated likelihood function (*p* and *r*) used in predictive inference.

| Parameter | Estimate (Units) | Comment |
| --- | --- | --- |
| $N$ | 470,000* | Population size |
| $t_0$ | 35* (d) | Start of COVID-19 transmission |
| $k$ | 6.6* | Shape parameter of $\Gamma(k, \theta)$ |
| $\theta$ | 7.9* | Scale parameter of $\Gamma(k, \theta)$ |
| $\mu_{LN}$ | 1.6** | μ-parameter of log-normal distribution |
| $\sigma_{LN}$ | 0.42** | σ-parameter of log-normal distribution |
| $p$ | Constrained*** | Probability parameter of NB($r, p$) |
| $r$ | 4.4* | Dispersal parameter of NB($r, p$) |

\*Estimates of the adjustable parameters ( $N$ , $t_0$ , $k$ , $\theta$ , and $r$ ) are region-specific and inference-time-dependent. Inferences are performed daily. Here, we report the maximum a posteriori (MAP) estimates inferred for New York City using all confirmed COVID-19 case-count data available in the GitHub repository maintained by *The New York Times* newspaper (11) for the period starting on 21-January-2020 and ending on 21-June-2020 (inclusive dates). Time $t = 0$ corresponds to 0000 hours on 21-January-2020. \*\*Estimates of the fixed parameters $\mu_{LN}$ and $\sigma_{LN}$ are those of Lauer et al. (12). These parameter estimates define a log-normal distribution that reproduces the empirical distribution of waiting times for the onset of symptoms after infection with SARS-CoV-2. \*\*\*The value of $p$ is constrained, i.e., its reporting-time-dependent value is determined by a formula, which is given by Equation (26).

**Appendix Figure 1**. Detailed diagram of the populations and processes considered in the mechanistic compartmental model. In this illustration of the compartmental model, the labels attached to arrows indicate the parameters and variables that affect the rates of the processes represented by the arrows. There is a one-to-one correspondence between arrow labels and terms on the right-hand sides of Equations (1)–(17). The diagram is otherwise the same as that shown in Figure 2.

**Appendix Figure 2**. We evaluated the potential impact of a one-time mass gathering that causes 50,000 and 5,000 individuals to become newly infected on 30-May-2020 in **(A)** the New York City metropolitan statistical area (MSA) and **(B)** the Phoenix MSA, respectively. As can be seen, such an event will step the epidemic curve up without significantly changing the slope of the curve. According to the model, the trend in the slope of the curve is determined by the sustained level of adherence to effective social-distancing practices. We conclude that the recent trajectory of the epidemic curve of the Phoenix MSA cannot be explained by a one-time mass gathering.

**Appendix Figure 3**. Predictive inferences conditioned on the compartmental model with either **(A)–(E)** one or **(F)–(J)** two distinct social-distancing periods. We consider five metropolitan statistical areas (MSAs), which according to Appendix Table 1, have epidemic curves better explained by the compartmental model with two social-distancing periods than by the compartmental model with just one. Maximum a posteriori (MAP) estimates for *τ*, indicate that the second social distancing period began on 27-May-2020 for Houston, 19-April-2020 for Miami, 24-May-2020 for Phoenix, 12-June-2020 for San Francisco, and 07-June-2020 for Seattle. In this analysis, we used the data available from 21-January-2020 to 26-June-2020 (inclusive dates).

**Appendix Figure 4**. Comparison of next-day predictions and the corresponding empirical case reports for **(A)** Houston, **(B)** Miami, **(C)** San Francisco, **(D)** Seattle, **(E)** Los Angeles, **(F)** Chicago, **(G)** Dallas, and **(H)** Washington, DC. Like Phoenix, Houston, Miami, San Francisco, and Seattle have epidemic curves that are better explained by the compartmental model with two social-distancing periods vs. just one (Appendix Table 1). Upward-trending anomalies were detected for each of these MSAs. Los Angeles, Chicago, Dallas, and Washington, DC have epidemic curves better explained by the compartmental model with just one social-distancing period vs. two (Appendix Table 1). Upward-trending anomalies were not detected for three of these MSAs. The exception is Chicago. Two anomalies were detected for Chicago near the end of the period considered in the analysis.

**Appendix Figure 5**. Out-of-sample validation of forecasting accuracy. For region-specific compartmental models with values of *n* determined by model selection, the empirical coverage was calculated for predicted detection of new cases 1, 4 and 7 days into the future (as indicated in the legend) over the period from 14-July-2020 to 9-September-2020. Coverage indicates the frequency at which the actual future number of new cases detected fell below the indicated predicted quantile. To create this figure, we combined the predictions and out-of-sample new-cases-detected data for the 15 metropolitan statistical areas (MSAs). The dotted line indicates the coverage expected for unbiased prediction.

**Appendix Figure 6**. Comparison of forecasting accuracy of deterministic and stochastic versions of the compartmental model for the New York City (NYC) metropolitan statistical area (MSA). In the three leftmost panels, credible intervals of predictive posteriors are shown. Inferences are based on the NYC data shown in the plots. At far left, results conditioned on the deterministic (ordinary differential equation, ODE) compartmental model are shown. In the two center plots, we show results conditioned on a comparable stochastic differential equation (SDE) model. Two realizations of the inference procedure were performed. In the rightmost panel, we show in-sample validation results. As can be seen, the ODE is less biased than the SDE model.

**Appendix Figure 7**. Comparison of forecasting accuracy of deterministic and stochastic versions of the compartmental model for the Miami metropolitan statistical area (MSA). In the three leftmost panels, credible intervals of predictive posteriors are shown. Inferences are based on the Miami data shown in the plots. At far left, results conditioned on the deterministic (ordinary differential equation, ODE) compartmental model are shown. In the two center plots, we show results conditioned on a comparable stochastic differential equation (SDE) model. Two realizations of the inference procedure were performed. In the rightmost panel, we show in-sample validation results. As can be seen, the ODE is less biased than the SDE model.

**Appendix Figure 8**. Illustration of shapes that can be produced by the fitting function that we used to capture trends in regional COVID-19 epidemic curves. The curve-fitting model is formulated such that it has the capacity to reproduce the shape of an epidemic curve having two timescales.

**Appendix Figure 9**. Bayesian predictive inferences for the 15 most populous metropolitan statistical areas (MSAs) in the United States. Predictions are conditioned on the curve-fitting model.

**Video 1**. An animation showing daily predictive inferences made for the New York City metropolitan statistical area from 19-Mar-2020 to 6-Jun-2020 (inclusive dates). Inferences are conditioned on the single-phase (*n* = 0) compartmental model.

**Video 2**. An animation showing daily predictive inferences made for the Phoenix metropolitan statistical area from 30-Mar-2020 to 17-Jun-2020 (inclusive dates). Inferences are conditioned on the single-phase (*n* = 0) compartmental model.

**Video 3**. An animation showing daily predictive inferences made for the Houston metropolitan statistical area from 30-Mar-2020 to 17-Jun-2020 (inclusive dates). Inferences are conditioned on the single-phase (*n* = 0) compartmental model.

**Video 4**. An animation showing daily predictive inferences made for the Miami metropolitan statistical area from 30-Mar-2020 to 17-Jun-2020 (inclusive dates). Inferences are conditioned on the single-phase (*n* = 0) compartmental model.

**Video 5**. An animation showing daily predictive inferences made for the San Francisco metropolitan statistical area from 30-Mar-2020 to 17-Jun-2020 (inclusive dates). Inferences are conditioned on the single-phase (*n* = 0) compartmental model.

**Video 6**. An animation showing daily predictive inferences made for the Seattle metropolitan statistical area from 30-Mar-2020 to 17-Jun-2020 (inclusive dates). Inferences are conditioned on the single-phase (*n* = 0) compartmental model.

**Video 7**. An animation showing daily predictive inferences made for the Los Angeles metropolitan statistical area from 30-Mar-2020 to 17-Jun-2020 (inclusive dates). Inferences are conditioned on the single-phase (*n* = 0) compartmental model.

**Video 8**. An animation showing daily predictive inferences made for the Chicago metropolitan statistical area from 30-Mar-2020 to 17-Jun-2020 (inclusive dates). Inferences are conditioned on the single-phase (*n* = 0) compartmental model.

**Video 9**. An animation showing daily predictive inferences made for the Dallas metropolitan statistical area from 30-Mar-2020 to 17-Jun-2020 (inclusive dates). Inferences are conditioned on the single-phase (*n* = 0) compartmental model.

**Video 10**. An animation showing daily predictive inferences made for the Washington, DC metropolitan statistical area from 30-Mar-2020 to 17-Jun-2020 (inclusive dates). Inferences are conditioned on the single-phase (*n* = 0) compartmental model.

## Appendix Text

### Full Description of the Mechanistic Compartmental Model

The compartmental model, which is illustrated in Appendix Figure 1, consists of the following 25 ordinary differential equations (ODEs):

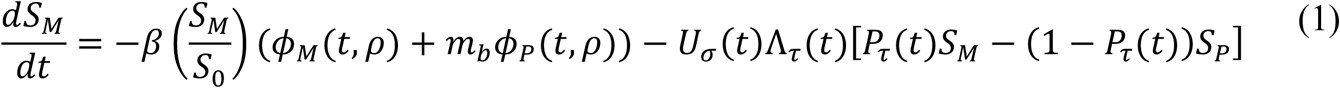

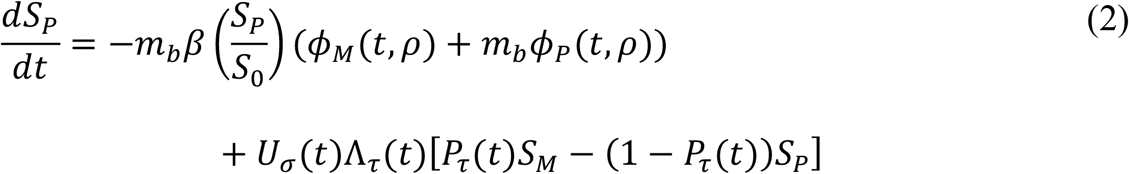

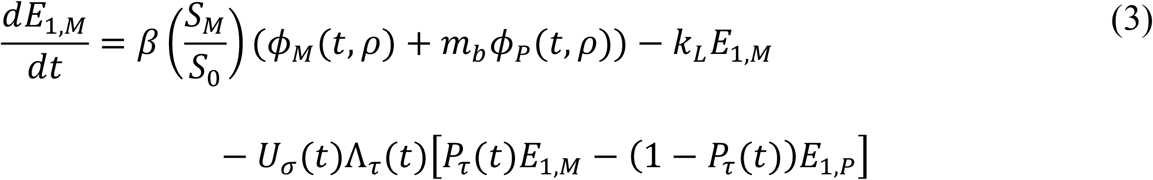

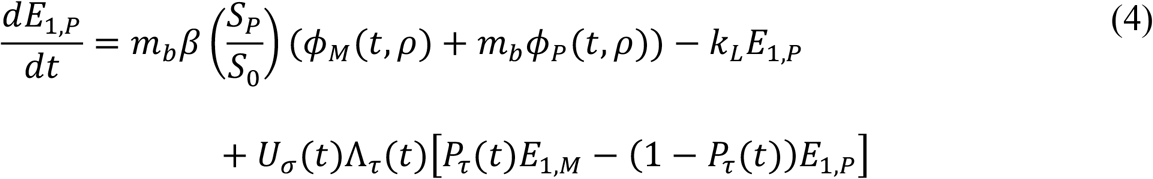

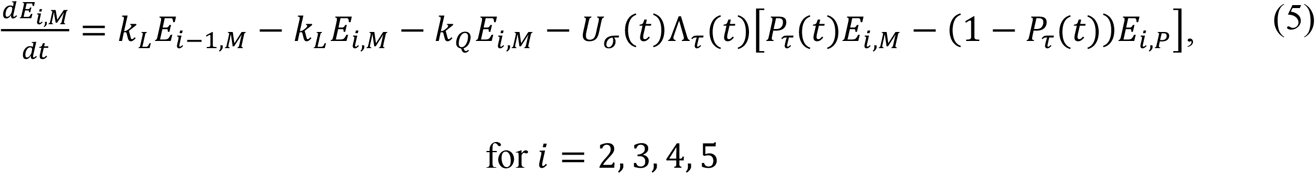

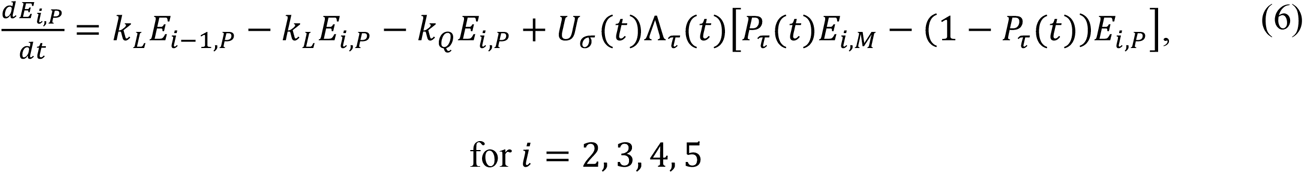

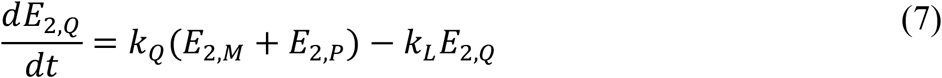

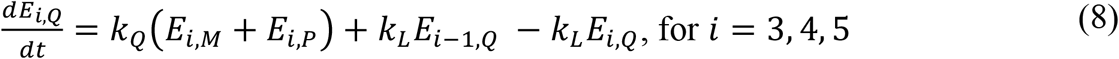

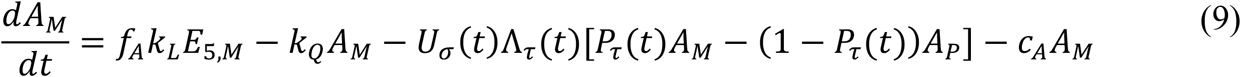

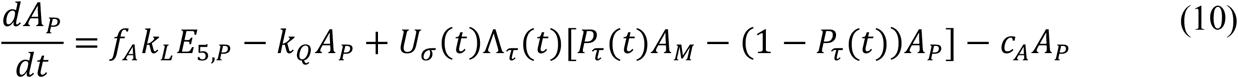

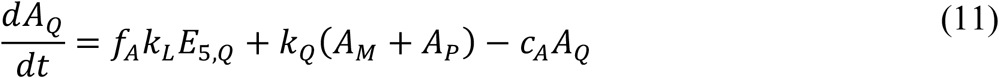

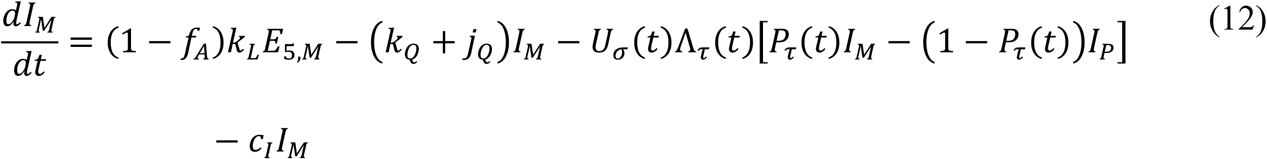

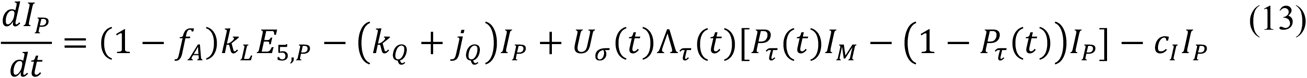

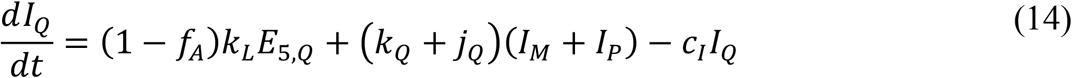

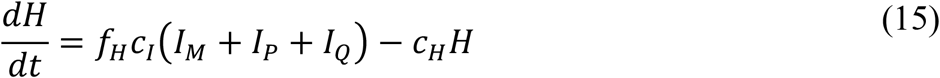

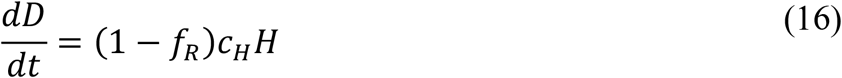

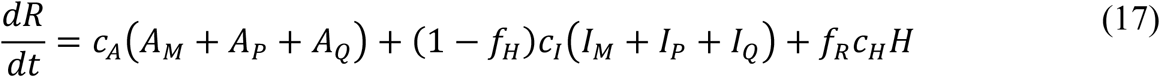

where *β, S*_0_, *m*_*b*_, *k*_*L*_, *k*_*Q*_, *j*_*Q*_, *f*_*A*_, *f*_*H*_, *f*_*R*_, *c*_*A*_, *c*_*H*_, and *c*_*H*_ are positive-valued time-invariant parameters. (It should be noted that parameter names are unique but only within the namespace of a given model.) Each ODE in Equations (1)–(17) defines the time-rate of change of a (sub)population, i.e., the time-rate of change of a state variable. There are 25 state variables, one for each ODE. Note that Equations (5), (6), and (8) define 4, 4, and 3 ODEs of the model, respectively. It should be noted that the model does not include generation of new cases by travelers.

The initial condition is taken to be *S*_*M*_(*t*_0_) = *S*_0_, *I*_*M*_ (*t*_0_) = *I*_0_ = 1, with all other populations (*S*_*P*_, *E*_1,*M*_, …, *E*_5, *M*_, *E*_1,*P*_, …, *E*_5,*P*_, *E*_2,*Q*_, …, *E*_5,*Q*_, *A*_*M*_, *A*_*P*_, *A* _*Q*_, *I*_*P*_, *I* _*Q*_, *H, D*, and *R*) equal to 0. The parameter *S*_0_ denotes the total region-specific population size. Thus, we assume that the entire population is susceptible at the start of the epidemic at time *t* = *t*_0_>0, where time *t* = 0 is 0000 hours on 21-January-2020. The parameter *I*_0_, which we always take to be 1, denotes the number of infectious symptomatic individuals at the start of the regional epidemic.

Subscripts attached to state variables are used to denote subpopulations. The subscripts *M* and *P* are attached to variables representing mixing and protected populations, respectively. For example, the variables *S*_*M*_ and *S*_*P*_ denote the population sizes of mixing and protected individuals who are susceptible to infection. Individuals in a protected population practice social distancing; individuals in a mixing population do not. The approach that we have taken to model social distancing is similar to that of Anderson et al. *(24)*.

The incubation period is divided into five stages. The numerical subscripts 1, 2, 3, 4, and 5 attached to *E* variables indicate progression through these five stages. Exposed individuals in the incubation period, except for those in the first stage, are taken to be infectious. They are also taken to lack symptoms. They are either presymptomatic (i.e., individuals who will later develop symptoms) or asymptomatic (i.e., individuals who will never develop symptoms).

The subscript *Q* is attached to variables representing populations of quarantined individuals. The state variable *I*_*Q*_ is a special case; it accounts for symptomatic individuals who are quarantined as well as individuals who are self-isolating because of symptom awareness.

The parameter *k*_*Q*_ characterizes the rate at which infected individuals move into quarantine because of testing and contact tracing. The parameter *j*_*Q*_ characterizes the rate at which symptomatic individuals self-isolate because of symptom awareness. We recognize that susceptible individuals may enter quarantine (through contact tracing) but we assume that the size of the quarantined population is negligible compared to that of the total susceptible population and that susceptible individuals entering quarantine leave quarantine as susceptible individuals.

The parameters *β* and *m*_*b*_ < 1 characterize transmission of disease: *β* characterizes the rate of transmission attributable to contacts between two mixing individuals, *m*_*b*_*β* characterizes the rate of transmission attributable to contacts between one mixing and one protected individual, and 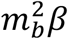 characterizes the rate of transmission attributable to contacts between two protected individuals. Infectious individuals taken to contribute to COVID-19 transmission include those in the following pools: *E*_2,*M*_, …, *E*_5,*M*_ and *E*_2,*p*_, …, *E*_5,*p*_, *A*_*M*_ and *A*_*P*_, and *I*_*M*_ and *I*_*P*_. Recall that we do not consider individuals in the first stage of the incubation period (i.e., individuals in *E*_1_ pools) to be infectious. The assumption is that these individuals are not shedding enough virus to be infectious (or detectable in surveillance testing). In experiments with an animal model (the golden hamster), infectious virus could be recovered from animals 2 d post-inoculation *(25)*. Moreover, it was found that SARS-CoV-2 could be detected in contacts of infected animals just 1 d post-contact *(25)*. Kucirka et al. *(26)* estimated that the false negative rate for nasal samples from exposed individuals tested for SARS-CoV-2 infection an estimated 1 d after exposure is 100% but less than 100% thereafter. Thus, it seems reasonable to assume that exposed individuals beyond the first incubation stage, which has a duration of approximately 1 d (based on our estimate for *k*_*L*_, which is discussed below), are infectious and may be detected as such.

The variables *E*_1,*M*_, …, *E*_5,*M*_ and *E*_1,*P*_, …, *E*_5,*P*_ denote the population sizes of mixing and protected exposed individuals in the five stages of the incubation period. The variables *E*_2,*Q*_, …, *E*_5,*Q*_ denote the population sizes of quarantined exposed individuals in the five stages. There is no *E*_1,*Q*_ population, as we assume that individuals in the first stage of the incubation period are unlikely to test positive for SARS-CoV-2 or to be reached in contact tracing efforts before leaving the *E*_1_ state. The parameter *k*_*L*_ characterizes disease progression, from one stage of the incubation period to the next and ultimately to an immune clearance phase. Individuals leaving the *E*_5_ pools enter the immune clearance phase, meaning that they become eligible for recovery. An individual leaving an *E*_5_ pool with symptom onset enters an *I* pool, whereas an individual leaving an *E*_5_ pool without symptom onset enters an *A* pool. Individuals in *I* pools are considered to have mild disease (with the possibility to progress to severe disease).

The dynamics of social distancing are characterized by three step functions (i.e., piecewise constant functions having only finitely many pieces): *U*_*σ*_, Λ_*τ*_, and *P*_*τ*_. The subscripts attached to these functions denote times: *σ* is a particular time, whereas *τ* is a set of times, as discussed later. The value of *U*_*σ*_ switches from 0 to 1 at time *t* = *σ* > *t*_0_, the start of an initial social-distancing period. As discussed later, the function Λ_6_ defines a timescale for change in social-distancing practices for one or more distinct periods of social distancing, and the function *P*_*τ*_ establishes a setpoint for the fraction of the total population of susceptible and infectious, non-quarantined/non-self-isolated/non-hospitalized individuals adhering to social-distancing practices for one or more distinct periods of social distancing.

The parameter *f*_*A*_ denotes the fraction of infected individuals who never develop symptoms (i.e., the fraction of all cases that are asymptomatic). The variables *A*_*M*_ and *A*_*P*_ denote the sizes of the populations of mixing and protected individuals who have been infected, progressed through the incubation period, are currently in the immune clearance phase, and will never develop symptoms. The parameter *c*_*A*_ characterizes the rate at which asymptomatic individuals recover. It should be noted that the duration of the immune clearance phase for asymptomatic individuals, 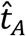, is distributed according to 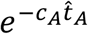. The mean value of 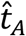 is 1/*c*_*A*_.

The variable *R* tracks recoveries of asymptomatic individuals, symptomatic individuals with mild disease, and symptomatic (hospitalized) individuals with severe disease. All individuals who recover are assumed to have immunity. This assumption is supported by the finding that SARS-CoV-2 infection elicits functional T cell memory *(27)*. Moreover, neutralizing antibodies evidently protect against SARS-CoV-2 infection *(28)*. Reinfection has been detected *(29)* but the importance of this apparently rare phenomenon has yet to be determined.

The variables *I*_*M*_ and *I*_*P*_ denote the sizes of the populations of mixing and protected symptomatic individuals with mild disease. The parameter *c*_*I*_ characterizes the rate at which symptomatic individuals with mild disease recover or progress to severe disease. The parameter *f*_*H*_ is the fraction of symptomatic individuals who progress to severe disease requiring hospitalization. As a simplification, we assume that all individuals with severe disease are hospitalized (or isolated at home in an equivalent state). It should be noted that the duration of the immune clearance phase for symptomatic individuals who never progress to severe disease, 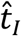, is distributed according to 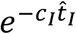. The mean value of 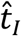 is 1/*c*_*I*_. As is implicit in our definition of *c*_*I*_, the time required for progression from mild to severe disease is taken to be the same as the recovery time of symptomatic individuals who experience only mild disease.

The variable *H* represents the population of hospitalized/severely ill individuals. In the model, these individuals are taken to be quarantined. Thus, the model does not consider nosocomial transmission. The parameter *f*_*R*_ denotes the fraction of hospitalized/severely ill individuals who recover. The parameter *c*_*H*_ characterizes the hospital discharge rate, i.e., the rate at which hospitalized individuals with severe disease either recover or die. The variable *D* tracks deaths. Many deaths occur outside a hospital setting *(30)*. As a simplification, the model does not distinguish between deaths at home and deaths in hospital. It should be noted that the mean duration of the immune clearance phase for hospitalized/severely ill individuals who recover, 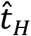, is distributed according to 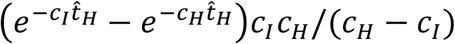, assuming *c*_*H*_ > *c*_*I*_. The mean value of 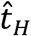 is 1/*c*_*I*_ + 1/*c*_*H*_. As is implicit in our definition of *c*_*H*_, the time required for progression from severe disease to death is taken to be the same as the recovery time of hospitalized/severely ill individuals.

The time-dependent terms *ϕ*_*M*_(*t, ρ*) and *ϕ*_*P*_(*t, ρ*) appearing in Equations (1)–(4) represent the effective population sizes of infectious individuals in the mixing and protected subpopulations, respectively. These quantities are defined as follows:

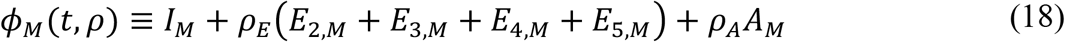

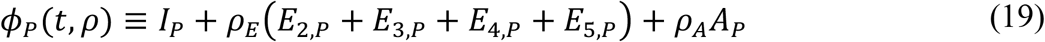

where *ρ* = (*ρ*_*E*_, *ρ*_*A*_), *ρ*_*E*_ is a constant characterizing the relative infectiousness of presymptomatic individuals compared to symptomatic individuals (with the same behaviors) and *ρ*_*A*_ is a constant characterizing the relative infectiousness of asymptomatic individuals compared to symptomatic individuals (with the same behaviors). Recall that infectiousness due to social-distancing behaviors is captured in Equations (1) and (2). Further recall that we assume that individuals in the first stage of the incubation period (i.e., individuals in either the *E*_1,*M*_ or *E*_1,*P*_ population) are not infectious. We also assume that the individuals in these populations cannot be quarantined until after transitioning to the *E*_2,*M*_ or *E*_2,*P*_ population (because they are assumed to test negative and because contact tracing is assumed to be too slow to catch individuals in the transient first stage of incubation). Recall that individuals in the *A*_*M*_, *A*_*P*_, and *A*_*Q*_ populations are defined as individuals who became infected, passed through all five stages of the incubation period, are currently in the immune clearance phase, and will never develop symptoms. Thus, individuals in the exposed *E* populations include both presymptomatic individuals (i.e., individuals who will enter the *I* populations) and asymptomatic individuals (i.e., individuals who will enter the *A* populations).

The time-dependent terms *U*_*σ*_(t), *P*_*τ*_(*t*), and Λ_*τ*_(*t*) appearing in Equations (1)–(6), Equations (9) and (10) and Equations (12) and (13) are step functions defined as follows:

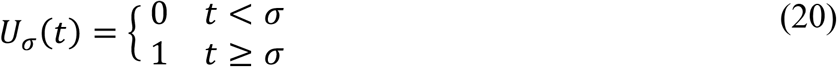

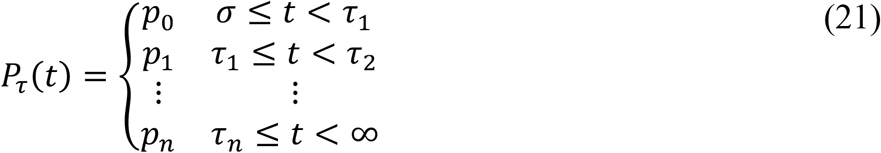

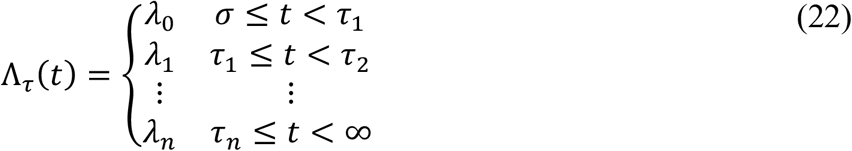

where *σ* > *t*_0_ is the time at which widespread social distancing initially begins, the integer *n* ≥ 0 is the number of societal (major/widespread) shifts in social-distancing practices after the initial onset of social distancing, each *p*_*i*_ < 1 is a parameter characterizing the quasi-stationary fraction of susceptible individuals practicing social distancing during the (*i* + 1)th period of social distancing, each *λ*_*i*_ is a constant defining a timescale for change in social-distancing practices during the (*i* + 1)th period of social distancing, *τ* = {*τ*_0_, …, *τ*_*n*+1_}, *τ*_0_ ≡ *σ, τ* _*n*+1_ ≡ ∞, and *τ*_*i*+1_ > *τ*_*i*_ for *i* = 0, …, *n* − 1. The value of *P*_*τ*_(*t*) defines a setpoint for the quasi-stationary size of the protected population of susceptible individuals: *P*_*τ*_(*t*) × 100% of the total susceptible population. The value of Λ_*τ*_(*t*) determines how quickly the setpoint is reached. As indicated in Equations (21) and (22), we only consider step-changes in the values of *P*_*τ*_(*t*) and Λ_*τ*_(*t*), a simplification. Thus, for a period during which social-distancing practices are intensifying (relaxing), we increase (decrease) the value of *P*_*τ*_(*t*) at the start of the period in a step-change and then hold it constant until the next step-change, if any. It should be noted that *σ* is the start time of the initial social-distancing period. The time at which the initial social-distancing setpoint, determined by *p*_0_, is reached occurs later and is determined by *λ*_0_. We caution the reader not to confuse the setpoint parameters *p*_0_, *p*_1_, …, *p*_*n*_ with the distributional parameter *p* in the negative binomial distribution N*B*(*r, p*).

### Full Description of the Auxiliary Measurement Model

To determine how consistent a particular parameterization of the compartmental model is with available COVID-19 surveillance data, we need to define a quantity—a model output—that corresponds to daily reports of the number of new confirmed COVID-19 cases. Case reporting by public health officials is typically daily. We expect that the vast majority of cases are detected because of symptom-driven (vs. random) testing and/or presentation in a clinical setting. Accordingly, as a simplification, we assume that individuals detected in surveillance are symptomatic. To define a model output comparable to the number of new cases reported on a given day, we start by considering the predicted cumulative number of presymptomatic individuals who become symptomatic while evading quarantine (because of contact tracing) until at least the onset of symptoms, which we will denote as *C*_*S*_. According to the model, the time rate of change of *C*_*S*_ is given by the following equation:

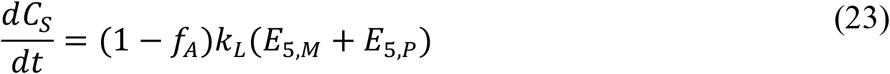

The right-hand side of this equation gives the rate at which non-quarantined presymptomatic individuals exit the incubation period and enter the immune clearance phase, in which they are symptomatic and therefore taken to be detectable in local surveillance efforts. We assume that symptomatic individuals in quarantine make a negligible contribution to detection of new cases.

Equation (23) and the ODEs of the compartmental model form a coupled system of equations, which can be numerically integrated to obtain trajectories for the state variables and *C*_*S*_, the expected cumulative number of symptomatic cases. From the trajectory for *C*_*S*_, we obtain a prediction for *I*(*t*_*i*_, *t*_*i*+1_), the expected number of new COVID-19 cases reported on a given calendar date 𝒟_*i*_, from the following equation:

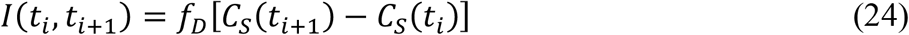

where *f*_*D*_ is taken to be an adjustable region-specific parameter characterizing the time-averaged fraction of symptomatic cases detected among non-quarantined and hospitalized individuals.

Equation (24) completes the formulation of our measurement model. *I*(*t*_*i*_, *t* _*i*+1_) is the model output that we compare to *δC*_*i*_, the number of new cases reported on calendar date 𝒟_*i*_.

The Adjustable and Fixed Parameters of the Compartmental Model and Auxiliary Measurement Model

The parameters of the compartmental model (Equations (1)–(22)) and the auxiliary measurement model (Equations (23) and (24)) are taken to have either adjustable or fixed values. The adjustable parameter values are estimated (daily) through Bayesian inference on the basis of surveillance data (i.e., reports of newly detected cases). The fixed parameter values are held constant during inference; they are based on non-surveillance data and/or assumptions, which are discussed in the section below. In this section, we simply delineate the parameters with adjustable and fixed values. The compartmental model formulated for a given regional epidemic has a total of 16 + 3(*n* + 1) parameters. The value of *n* is structural; it sets the number social-distancing periods considered.

The value of *n* corresponds to the number of periods of distinct social-distancing behaviors that follow an initial period of social distancing, which we take to begin at time *t* = *σ* > *t*_0_. Here, we take *n* = 0 or 1 for all regional epidemics of interest. Initially, we set *n* = 0. In cases where we set *n* = 1, this setting was motivated by second wave-type dynamics suggested by the surveillance data, which we take to indicate a relaxation of social-distancing practices at time *t* = *τ*_1_ > *σ*. The parameters of the initial social-distancing period are *σ, p*_0_, and *λ*_0_. The parameters of the second social-distancing period, if considered, are *τ*_1_, *p*_1_, and *λ*_1_. Thus, there are 3(*n* + 1) social-distancing parameters, all of which are taken to be adjustable.

In addition to the 3(*n* + 1) social-distancing parameters, we have 16 other parameters. Three of these define the initial condition: *t*_0_, *S*_*M*_(*t* = *t*_0_) = *S*_0_, and *I*_*M*_(*t* = *t*_0_) = *I*_0_, where *t*_0_ is the time at which the epidemic begins, *S*_0_ is taken to be the total population of the region of interest, and *I*_0_ (the initial number of infected individuals) is always taken to be 1 (an assumption). We take *t*_0_ to be adjustable and *S*_0_ and *I*_0_ to be fixed. The value of *S*_0_ is set on the basis of population estimates by the US Census Bureau for the metropolitan statistical areas of interest *(13)*, which are delineated by the US Office of Management and Budget *(10)*.

There is only one more adjustable parameter of the compartmental model: *β*, which characterizes the rate of disease transmission attributable to contacts among individuals within the mixing population. In the period before the onset of social distancing, from *t*_0_ to *σ*, when *S*_*M*_/*S*_0_ ≈ 1, the instantaneous rate of disease transmission is *βϕ*_*M*_(*t, ρ*), where *ϕ*_*M*_(*t, ρ*) is the effective number of infectious individuals at time *t*, a weighted sum of the numbers of symptomatic, presymptomatic, and asymptomatic individuals determined by *ρ* = (*ρ*_*E*_, *ρ*_*A*_). We assume that exposed individuals after the first stage of disease incubation are infectious, as are asymptomatic individuals in the immune clearance phase who have passed through all 5 stages of disease incubation and who will never develop symptoms.

The remaining 12 parameters of the compartmental model, which are taken to have fixed, region-independent values, are as follows: *m*_*b*_, *ρ*_*E*_, *ρ*_*A*_, *k*_*L*_, *k*_*Q*_, *j*_*Q*_, *f*_*A*_, *f*_*H*_, *c*_*A*_, *c*_*I*_, *f*_*R*_ and *c*_*H*_. Our estimates for these parameters are discussed in the section immediately below. It should be noted that settings for *f*_*R*_ and *c*_*H*_ do not affect predictions of new cases because these parameters characterize recovery/morbidity of hospitalized individuals. The parameter *f*_*R*_ is the fraction of hospitalized individuals who recover, and the parameter *c*_*H*_ characterizes the hospital discharge rate. Although nosocomial disease transmission is a significant concern, we assume that hospitalized individuals are effectively quarantined such that the overall rate of disease transmission in a given region is insensitive to the number of hospitalized individuals in that region.

### Estimates of 12 Fixed Parameter Values of the Compartmental Model

Here, in this section, we summarize the rationale/justification for each of our estimates for the values of the following 12 parameters of the compartmental model: *m*_*b*_, *ρ*_*E*_, *ρ*_*A*_, *k*_*L*_, *k*_*Q*_, *j*_*Q*_, *f*_*A*_, *f*_*H*_, *c*_*A*_, *c*_*I*_, *f*_*R*_, and *c*_*H*_. These estimates are assumed to apply to all regions, i.e., we take these parameters to have region-independent values. The estimates given below should be understood to be rough and provisional. The information available to inform these estimates is limited. Although using point estimates for some of the model parameters can lead to underestimates of parametric uncertainty *(31)*, it is necessary to aggressively leverage prior knowledge to reduce the number of adjustable parameters, because it is not feasible to infer all of the model parameters from case reporting data. For each region of interest, we focus on inferring model parameters that characterize when disease transmission started (*t*_0_), how disease transmission depends on behavior (*σ, p*_0_, *λ*_0_, and *β*), and surveillance (*f*_*D*_ and *r*). Given 1) the data streams being analyzed, 2) the evident importance of behavior/social distancing on disease transmission, and 3) our goal of situational awareness, it seems reasonable to focus on inference of these parameters. (As we discuss below, we fix the value of *m*_*b*_, which characterizes social distancing, only because we found it to be correlated with the value of *p*_0_, another social-distancing parameter, when both are inferred.)

The parameter *m*_*b*_ characterizes the effects of social distancing on disease transmission. Without social distancing, all contacts responsible for disease transmission are between mixing individuals (i.e., between individuals in the *I*_*M*_ and *S*_*M*_ pools) and the rate of transmission is characterized by *β*. With social distancing, there are contacts involving 1 individual in a mixing population and 1 individual in a protected population (e.g., between individuals in the *I*_*M*_ and *S*_*P*_ pools or in the *S*_*M*_ and *I*_*P*_ pools) and also contacts involving 2 individuals in protected populations (e.g., between individuals in the *I*_*M*_ and *S*_*P*_ pools). In the model, the rates of transmission associated with these types of contacts are characterized by *m*_*b*_*β* and 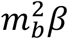, respectively. We can be confident that social distancing is protective (i.e., *m*_*b*_ < 1) but there is little information available to suggest the magnitude of the effect. We arbitrarily set *m*_*b*_ = 0.1, which can be interpreted to mean that a susceptible individual practicing social distancing has a 10-fold smaller chance of becoming infected than a susceptible individual that is not practicing social distancing. In exploratory analyses, wherein we allowed *m*_*b*_ to be a free parameter, we found that its inferred value is positively correlated with the extent of social distancing, which is determined by the relevant social-distancing setpoint parameter (for example, *p*_0_ during the initial social-distancing period). Thus, we interpret the inferred quasi-stationary value of *S*_*P*_ to be an effective population size. If our estimate for *m*_*b*_ is too high (i.e., we underestimate the protective effect of social distancing), the effective size will be larger than the true size. Conversely, if our estimate for *m*_*b*_ is too low, the effective size will be smaller than the true size.

The parameters *ρ*_*E*_ and *ρ*_*A*_ characterize the relative infectiousness of individuals without symptoms during the incubation period and the immune clearance phase, respectively. Infectiousness is compared to that of a symptomatic individual. Using a one-step real-time reverse transcriptase-polymerase chain reaction (rRT-PCR) assay to quantify viral RNA abundance in nasopharyngeal and oropharyngeal samples, Arons et al. *(14)* determined rRT-PCR threshold cycle (Ct) values for 17 symptomatic and 24 presymptomatic individuals. The former group of individuals had typical symptoms, and the latter group of individuals lacked symptoms at the time of testing but later developed symptoms (within 1 week after testing). At the time of testing, the median Ct values for symptomatic and presymptomatic individuals were 24.8 and 23.1, respectively. (NB: Ct value is inversely proportional to abundance.) Based on these results and an assumption that infectiousness is proportional to viral load, we estimate that *ρ*_*E*_ = 1.1. An estimate for *ρ*_*E*_ greater than 1 is consistent with the findings of He et al. *(32)*, who inferred that viral load is maximal 0.7 d before the onset of symptoms from an analysis of temporal viral load data and information available about infector-infectee transmission pairs. A review of the literature by Benefield *(33)* indicates that viral load is maximal before onset of symptoms. Over a period of 19 d, Nguyen et al. *(15)* performed daily rRT-PCR assays for viral RNA in nasopharyngeal samples from 17 symptomatic and 13 asymptomatic individuals. Ngyuen et al. *(15)* developed a curve-fitting model for each group to characterize their viral decay kinetics. These models indicate that the mean Ct value for symptomatic individuals was roughly 90% of the mean Ct value for asymptomatic individuals over the first week of the study, after which most individuals tested negative or had a Ct value near the threshold of detection, 40. Thus, we estimate that *ρ*_*A*_ = 0.9. Estimates of *ρ*_*E*_ and *ρ*_*A*_ should be considered crude.

The parameter *k*_*L*_ characterizes the duration of the incubation period. In the model, the incubation period is divided into 5 stages (for reasons explained shortly). The waiting time for completion of all 5 stages is described by an Erlang distribution with a shape parameter *k* = 5 and a scale parameter *μ* = 1/*k*_*L*_. Lauer et al. *(12)* estimated times of exposure and symptom onset for 181 confirmed cases and found that the median time between SARS-CoV-2 infection and onset of COVID-19 symptoms is 5.1 d. Lauer et al. *(12)* also found that the empirical distribution of waiting times is fit by an Erlang distribution with *k* = 6 and *μ* = 0.88 d. This latter finding suggests that the empirical waiting time distribution can be reproduced by dividing the incubation period into 6 stages and setting *k*_*L*_ = 1.14 *D*^−1^. However, an Erlang distribution with *k* = 5 and *μ* = 1.06 d has a nearly identical shape. Because simulation costs are reduced by dividing the incubation period into 5 instead of 6 stages, we considered 5 stages in the model. The distribution of waiting times estimated by Lauer et al. *(12)* is reproduced by our model when we set *k*_*L*_ = 0.94 *D*^−1^.

The parameters *k*_*Q*_ and *j*_*Q*_ characterize testing-driven quarantine and symptom-driven self-isolation. We assume that testing is random. Thus, the number of infected individuals moving into quarantine per d is the number of infected individuals subject to quarantine times the fraction of the total population tested per d times a multiplier capturing the effect of contact tracing. We take the multiplier to be average household size, 2.5 (US Census Bureau). Thus, based on approximately 500,000 tests per d in the US (https://covidtracking.com/data/us-daily) and a total population of 330 million (https://www.census.gov/popclock/), we estimate *k*_*Q*_ = 0.0038 *D*^−1^. It should be understood that the *k*_*Q*_ parameter, which characterizes the rate at which exposed individuals move to quarantine because of testing and contact tracing, incorporates factors such as false negative test results. As a simplification, we assume that *k*_/_ is the same for each stage of disease progression. We assume *j*_*Q*_ = 0.4 *D*^−1^. With this setting, the median waiting time from onset of symptoms to initiation of self-isolation is approximately 40 h. A faster timescale for self-isolation is probably not realistic despite general awareness of the COVID-19 pandemic, because as considered in the study of Böhmer et al. *(34)*, for any given individual, there may be a prodromal phase of ∼1 d marked by non-COVID-19-specific symptoms other than fever and cough.

The parameter *f*_*A*_ is the fraction of infected individuals who never develop symptoms. We estimate *f*_*A*_ on the basis of information about the COVID-19 outbreak on the *Diamond Princess* cruise ship, as recounted by Sakurai et al. *(17)*. See also Refs. *(35)* and *(36)*. Before disembarking, 3,618 passengers and crew members were tested for SARS-CoV-2 infection. 410 of 712 individuals testing positive for SARS-CoV-2 were without symptoms at the time of testing. The Ministry of Health, Labour, and Welfare of Japan *(16)* reported that 311 (76%) of these individuals remained asymptomatic over the course of long-term follow-up. See also Refs. *(35)* and *(36)*. Thus, we estimate that 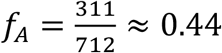. This estimate is consistent with the results of other studies. Lavezzo et al. *(37)* estimated that 43% of all infections are asymptomatic. In the study of Gudbjartsson et al. *(38)*, 7 of 13 individuals detected to have SARS-CoV-2 infection in random-sample population screening did not report symptoms; 43% of all SARS-CoV-2-positive participants in the study were symptom-free.

The parameter *f*_*H*_ is the fraction of symptomatic individuals progressing to severe disease. We set *f*_*H*_ such that our model predicts a uniform infection fatality rate (IFR) consistent with that determined by Perez-Saez et al. *(18)* from serological survey results and death incidence reports: 0.0064 (∼0.64%). For a discussion of other IFR estimates, which tend to be similar, see Grewelle and De Leo *(39)*. According to our model, IFR is given by (1 − *f*_*A*_)*f*_*H*_(1 − *f*_*R*_). This quantity is the fraction of all infected individuals predicted to develop symptoms and then to progress to severe disease (and hospitalization) and finally a fatal outcome. Thus, based on our estimates for *f*_*A*_ (0.44) and *f*_*R*_ (0.79) and the empirical IFR (0.0064), we set 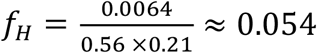.

The parameter *c*_*A*_ characterizes the duration of infectiousness of asymptomatic individuals in the immune response phase. For each of 89 asymptomatic individuals, Sakurai et al. *(17)* reported the time between the first positive PCR test for SARS-CoV-2 and the first of two serial negative PCR tests. The mean duration of this period was ∼9.1 d. We assume that this period coincides with the period of infectiousness and that this period encompasses both the incubation period and the immune response phase. With the incubation period for both presymptomatic and asymptomatic individuals divided into five stages of equal mean duration 1/*k*_*L*_, the overall mean duration of the incubation period is 5/*k*_*L*_. Based on our earlier estimate that *k*_*L*_ = 0.94 *d*^−1^, the mean duration of the incubation period is estimated as 5.3 d. Accordingly, the mean duration of the immune clearance phase for asymptomatic individuals is estimated as 9.1 *d* − 5.3 *d* = 3.8 *d*, and it follows that 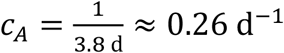

If *f*_*H*_ ≪ 1, the parameter *c*_3_ characterizes the duration of infectiousness of individuals who develop mild COVID-19 symptoms (i.e., symptoms not severe enough to require hospitalization). Wölfel et al. *(20)* attempted to isolate live virus from clinical throat swab and sputum samples collected from 9 patients at multiple time points after the onset of mild COVID-19 symptoms. Roughly 67%, 38%, and 0% of attempts to isolate virus were successful at 6, 8, and 10 d after infection, respectively. Assuming that a negative culture coincides with loss of infectiousness, we estimate that 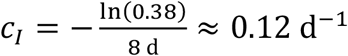.

The parameters *f*_*R*_ and *c*_*H*_ characterize the hospital stays of the severely ill. These parameters affect predictions of COVID-19-caused deaths and hospital resource utilization but do not affect the predicted transmission dynamics, because we assume that hospitalized patients are effectively quarantined and do not contribute significantly to disease transmission, i.e., there is no *I*_*H*_ term in *ϕ*_*M*_ or *ϕ*_*P*_ (see Equations (18) and (19)). The parameter *f*_*R*_ is the fraction of hospitalized patients who recover, and the parameter *c*_*H*_ characterizes the rate at which patients are discharged (as either recovered or dead). Richardson et al. *(19)* reported that the overall median length of hospital stay for 2,634 discharged patients (alive or dead) was 4.1 d. Thus, we estimate that 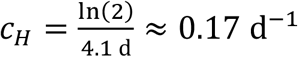. Among the discharged patients, 553 (21%) died. Thus, we estimate that *f*_*R*_ = 0.79.

#### Likelihood Function Used in Inference of Model Parameter Values

We assume that the likelihood of a set of adjustable parameter values *θ*_*F*_ given a report of *δC*_*i*_ new cases on calendar date 𝒟_*i*_, which we will denote as ℒ_*i*_ (*θ*_*F*_; *δC*_*i*_), is given by the following equation:

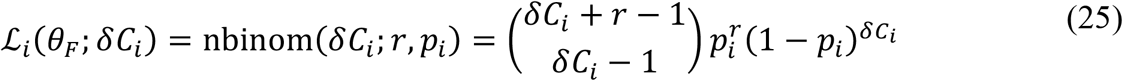

where *δC*_*i*_ is a non-negative integer (the number of new cases reported), *i* is an integer indicating the date 𝒟_*i*_ or the period (*t*_*i*_, *t*_*i*+1_); nbInoM(*δC*_*i*_; *r, p*_*i*_) is the probability mass function of the negative binomial distribution N*B*(*r, p*_*i*_), which has two parameters, *r* > 0 and *p*_*i*_ ∈ [0,1]; and *θ*_*F*_ is a model-dependent ordered set of feasible (i.e., allowable) values for the adjustable model parameters (e.g., *N, t*_0_, *k*, and *θ* in the case of the curve-fitting model) augmented with a feasible value for *r*. Recall that *t*_*i*_ ≡ *t*_0_ + *i D*, where *t*_0_ > 0 is 0000 hours of 𝒟_0_, the start date of the local epidemic. We take the dispersion parameter *r* of N*B*(*r, p*_*i*_) to be date/time-independent (i.e., applicable to all surveillance days) and infer the value of *r* jointly with the values of the model parameters.

Recall the following modeling assumptions. We take surveillance testing to be a stochastic process, i.e., we assume that the fraction of cases detected varies stochastically from day to day. Furthermore, we take the deterministic compartmental model parameterized for a given region to reproduce the mean daily outcomes of testing in that region. We also assume that the randomness in the number of new cases detected on a given date *t*_*i*_ is captured by a negative binomial distribution N*B*(*r, p*_*i*_), where *r* is a testing date-independent (i.e., *t*_*i*_-independent) parameter and *p*_*i*_ is a testing date-dependent (i.e., *t*_*i*_-dependent) parameter. The above assumptions mean that, for each date *t*_*i*_, we are taking *I*(*t*_*i*_, *t*_*i*+1_), the predicted number of new cases, to correspond to 𝔼[N*B*(*r,p*_*i*_)], which equals *r*(1 − *p*_*i*_)/*p*_*i*_. For this relationship to hold true, each distributional parameter *p*_*i*_ must satisfy the following constraint:

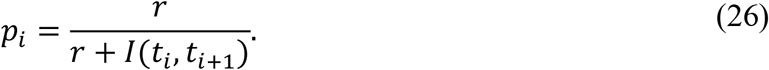

We use this constraint to determine the value of *p*_*i*_ for all *t*_*i*_. Recall that the value of *r* is jointly inferred with the values of the adjustable compartmental model parameters.

If *m* + 1 daily case reports are available, from date 𝒟_0_ to date 𝒟_*M*_, we assume that each likelihood ℒ_*i*_ (*θ*_*F*_; *δC*_*i*_) given by Equations (25) and (26) is independent. Thus, we have

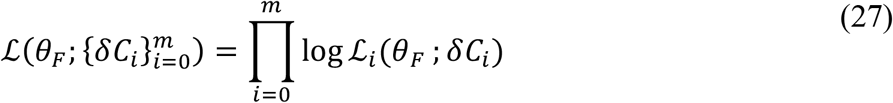

where 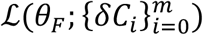 is the likelihood of *θ*_*F*_ given all available case reports 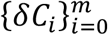. Recall that *δC*_*i*_ is the number of new cases reported on date 𝒟_*i*_ and *I*(*t*_*i*_, *t*_*i*+1_) is the model prediction of *δC*_*i*_. Furthermore, recall that *θ*_*F*_ is defined as a model-dependent ordered set of feasible adjustable model parameter values augmented with a feasible value for the likelihood function parameter *r*. The identity of *θ*_*F*_ depends on whether we are using Equation (27) to make inferences conditioned on the curve-fitting model or the compartmental model (i.e., we use Equation (27) in both cases but the identity of *θ*_*F*_ depends on the model being considered). The ordering of parameter values within the set *θ*_*F*_ is arbitrary but should be consistent.

When Equations (25)–(27) are used with the compartmental model having structure defined by *n* = 0, the elements of *θ*_*F*_ are *t*_0_, *σ, p*_0_, *λ*_0_, *β, f*_*D*_, and *r*, and *I*(*t*_*i*_, *t*_*i*+1_) is obtained from Equation (24). When Equations (25)–(27) are used with the curve-fitting model (see below), the elements of *θ*_*F*_ are *N, t*_0_, *k, θ*, and *r*, and *I*(*t*_*i*_, *t*_*i*+1_) is obtained from Equation (31) (see below).

#### Bayesian Inference and Online Learning

We chose the Bayesian inference framework to parametrize the models with uncertainty quantification. In Bayesian inference, given a set of data *D*, the probability of each set of the parameters, denoted in *θ*_*F*_ is constrained by the Bayes formula

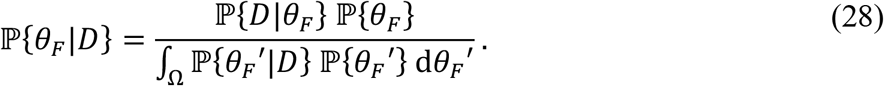

Here, the ℙ{*θ*_*F*_} is the prior parameter distribution, which represents our belief of how the model parameters should distribute in the parameter space Ω, and ℙ{*D*|*θ*_*F*_} is the likelihood of the parameter set *θ*_*F*_ given the dataset *D*, that is, 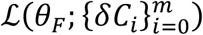 in Equation (27). In general, evaluating the posterior parameter distribution ℙ{*θ*_*F*_|*D*} is a difficult computation, mainly because of the high-dimensional integration of the term ∫_Ω_ ℙ{*θ*_*F*_′|*D*} ℙ{*θ*_*F*_′} *dθ*_*F*_′, a term often referred to as the *evidence*. Thus, for high-dimensional models, one relies on Markov chain Monte Carlo (MCMC) techniques to sample the posterior parameter distribution ℙ{*θ*_*F*_|*D*}.

In contrast of many modeling analyses which focus on identifying the parameter distributions, we are interested in projections of the models, whose parameters are inferred by past data, into the future. To this end, we evaluate the model with a probabilistic parameter set distributed by the obtained posterior distribution ℙ{*θ*_*F*_|*D*}. Formally, we denote the prediction of the confirmed cases between future day *t*_*i*_ and *t*_*i*+1_ by our deterministic model with a set of parameters *θ*_*F*_ by *I*(*t*_*i*_, *t*_*i*+1_; *θ*_*F*_). Recall that this deterministic prediction represents the mean of the fundamentally random new confirmed cases reported in a future interval (*t*_*i*_, *t*_*i*+1_). If there was only parametric uncertainty which propagates through the deterministic model, the confirmed cases reported in a future interval (*t*_*i*_, *t*_*i*+1_) would be distributed according to ∫Ω *I*(*t*_*i*_, *t*_*i*+1_; *θ*_*F*_) ℙ{*θ*_*F*_|*D*} *dθ*_*F*_. However, there is also observation noise, which we model by a negative binomial distribution. The observation noise also needs to be injected into the prediction to quantify the full uncertainty. The full prediction accounting for parametric uncertainty is a random variable distributed according to

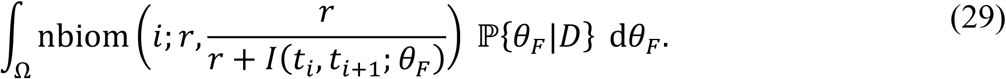

In practice, the above random variable is resampled from the posterior chain derived from the MCMC sampling. We denote the MCMC posterior chain by 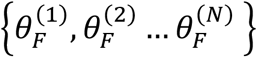. We sample the posterior chain and denote the resampled parameter set by 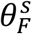 and the deterministic prediction of that resampled parameter in interval (*t*_*i*_, *t*_*i*+1_) by 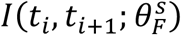. Then, we generate a negative binomial random number with the first parameter of the negative binomial distribution set as 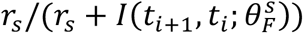 where *r*_*S*_ is the resampled *r* which is also a free parameter in 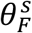 and is inferred in the MCMC. We repeat the resampling procedures and use the generated samples to compute the percentile of the past history and future prediction.

Our aim is to perform the Bayesian inference daily as soon as a new regional confirmed case number is reported. Although the Bayesian framework allows a sequential analysis, that the previous derived posterior distribution is used as a prior and the new inference problem involves only one new data point, in practice, such an analysis is difficult if the posterior distribution cannot be emulated or interpolated from the discrete posterior chain. Our analysis shows that in some regions, the posterior is far from Gaussian, making accurate interpolation or emulation difficult. Thus, we are not adopting this workflow, and instead, we perform the inference with all the data points collected up to the time of inference. Nevertheless, we warm-start the MCMC chain from the maximum a posteriori estimator estimated from the previous derived chain, and we also use the previously derived chain for estimating the optimal covariance matrix for the proposal of the normal symmetric random-walk Metropolis sampler. This approach allows an online learning of the optimal proposal which significantly reduces the mixing time.

Technical Details of Approach and Numerical Methods Used in Bayesian Inference

Because the variability of the data due to the regional and temporal differences, it is difficult to identify a universal sampling strategy (the proposal kernel). Thus, we adopted an adaptive Metropolis algorithm, specifically Algorithm 4 in Andrieu and Thoms *(21)* to accommodate the regional and temporal differences.

For all the model parameters, we assume their priors are uniformly distributed. Denote the 0:00 of the calendar date of the first confirmed case in a specific region by *t*_first_ and the total population of that region by *S*_0_. For the curve-fitting model, we assume the parameters are bounded by *N* ∈ (0, *S*_0_), *t*_0_ ∈ (*t*_first,_ − 21, *t*_first,_), *μ* ∈ (0,10^6^), *k* ∈ (0,10^6^), *θ* ∈ (0,10^6^). For the compartmental model with *n* = 0, we assume that the parameters are bounded by *t*_0_ ∈ (0, *t*^⋆^), *σ* ∈ (*t*_0_, *t*^⋆^ − *t*_0_), *β* ∈ (0,10^*6*^), *λ*_0_ ∈ (0,10), *p*_0_ ∈ (0,1), and *f*_*D*_ ∈ (0,1), where *t*^⋆^ is the time at which inference is performed. We assume *r* ∈ (0,10^6^) for both models. The limits on parameter values reflect feasibility constraints or bounds determined by trial-and-error so as to ensure that the posterior mass is contained within the hypercube defined by the limits. We adopted rejection-based sampling to assure the parameter values are sampled in the hypercube.

We start inference with an isotropic proposal kernel, that randomly perturbs the parameter values by independent Gaussian proposals whose standard deviations are set to be 5% of the parameter values. We carry out the standard Metropolis–Hastings algorithm for 5 × 10^4^ iterations first to identify local minimum. Then, we turned on the adaptive Metropolis algorithm to calculate the covariance matrix on-the-fly, for another 5 × 10^4^ iteration, when we turned on an on-the-fly learning for optimal proposed increment, i.e., *λ* in Algorithm 4 of Andrieu and Thoms *(21)*. Because the weight for learning the proposed increment decays 1/Iteration *(21)*, the proposed increment stabilizes after about 10^3^ more iterations. We began to collect the statistics from 1.5 × 10^5^ iteration, until the simulation finishes at 6 × 10^5^ iterations.

Except for the first time of the procedure (i.e., online learning and day-to-day operation), we warm start the simulation from the previously obtained best-fit (maximum a posteriori (MAP) estimator) and with the previously obtained covariance matrix and proposed increment. We carry out standard Metropolis–Hastings algorithms for 2.5 × 10^4^ iterations first to identify a local minimum, noting that it is often not far away from the previously identified MAP. We then turn on the adaptive MCMC algorithm to calculate the covariance matrix on-the-fly, again for another 5 × 10^4^ iteration. We then use another 2.5 × 10^4^ iterations to calculate the optimal proposed increment. We start to collect statistic from 10^5^ iteration to 4 × 10^5^ iteration when the simulation finishes.

#### Model Selection

A heuristic model-selection procedure is used to select the most parsimonious value of *n*, which determines the structure of the compartmental model (i.e., the number of social distancing periods considered) and the number of adjustable parameters describing social distancing (three for each social-distancing period). The procedure for deciding between *n* = 0 and *n* = 1 is as follows. We calculate Aikake information criterion (AIC) *(40)* values for the *n* = 0 and *n* = 1 versions of the compartmental model. Similarly, we calculate Bayesian information criterion (BIC) *(40)* values for the two versions of the model. We define Δ*AIC* as *AIC*^n=0^ − *AIC*^n=1^and Δ*BIC* as *BIC*^n=0^− *BIC* ^n=1^, where *AIC* ^n=0^ (*AIC* ^n=1^) is the AIC value calculated for the *n* = 0 (*n* = 1) version of the compartmental model and, similarly, *BIC*^n=0^ (*BIC*^n=1^) is the BIC value calculated for the *n* = 0 (*n* = 1) version of the compartmental model. The recommendations of Burnham and Anderson *(40)* indicate that we can interpret Δ*AIC*>10 to mean that the evidence strongly supports the *n* = 1 version of the model. In other words, we are justified in using *n* = 1 (the more complex version of the model) instead of *n* = 0 when Δ*AIC*>10. However, we take a more conservative approach. We adopt *n* = 1 over *n* = 0 only when both Δ*AIC*>10 and Δ*BIC*>10. We are using the same approach described above to decide between *n* = 1 and *n* = 2. Prediction Updates

Daily predictions based on region-specific parameterizations of the compartmental model are being archived at a GitHub repository (https://github.com/lanl/COVID-19-Predictions). It should be noted that the model’s structural parameter *n*—which determines the number of distinct social-distancing periods considered in the model—varies from region to region and can change over time.

#### Forecasting Accuracy

We performed out-of-sample testing of forecasting accuracy for inferences conditioned on the compartmental model with the value of *n* chosen through model selection. For predictions 1, 4, and 7 days ahead, we determined the empirical coverage, meaning the frequencies characterizing how often the empirical data fell below predicted quantiles. In this analysis, we considered surveillance data available between 14-July-2020 and 9-September-2020 (inclusive dates). The results obtained using the combined out-of-sample data and predictions for all 15 MSAs of interest are shown in Appendix Figure 5. As can be seen, overall, predictions are (weakly) biased toward underprediction of new case detection.

### Evaluation of a Stochastic Version of the Compartmental Model

A compartmental model may be formulated as either a deterministic model or a stochastic model. Use of a deterministic model is a potential liability, because there are situations where the assumptions of a deterministic model break down, necessitating use of a stochastic model. For this reason, we performed an analysis to determine which of the two model types (deterministic or stochastic) is better suited for analysis of the data of interest here.

We constructed an individual-based continuous-time Markov chain with consideration of the same processes as those in our deterministic compartmental model. The intrinsic stochasticity arises from the discreteness of populations. We followed the standard procedure to construct a set of stochastic differential equations (SDEs), by first formulating a master equation, then performing the Kramers-Moyal expansion to obtain the approximate continuum-limit Fokker-Planck equation in the large-population limit, and formulating the corresponding SDEs. The procedure has been described, for example, by Lin et al. *(41, 42)*.

In simulations, we used the Euler-Maruyama integrator to evolve the SDEs with a time step of 0.05 (d). We checked to make sure the timestep was sufficiently small.

We adopted a standard particle filter technique to identify the Maximum Likelihood Estimator of the parameters of the stochastic model, noting that our process is not time-homogeneous due to different episodes of distinct social-distancing practices. We used the data from the New York City and Miami MSAs from 21-Jan-2020 to 29-Aug-2020 to parametrize the stochastic model.

After identifying the point estimator, the predicted quantiles were calculated similarly as for the deterministic model. Finally, we quantified the in-sample empirical coverage, i.e., the empirical frequencies quantifying when training-data points fall below the predicted quantiles. When the noise model is correct, one would expect that such frequencies should coincide with the prediction.

For the New York City MSA, we obtained the results shown in Appendix Figure 6. For the Miami MSA, we obtained the results shown in Appendix Figure 7. By visually inspecting the predictive posteriors, we can see that the stochastic models lead to higher uncertainty than the deterministic models for New York City and Miami. The right-most panels, the in-sample calibration curves, show that the SDE model in both cases is farther away from the diagonal than the deterministic model. These results indicate that the stochastic models assert more uncertainty than the data distribution. Thus, it seems more appropriate to use a deterministic version of the model for forecasting than a stochastic version, at least at this time.

### Evaluation of a Curve-Fitting Model

Various fitting functions, or curve-fitting models, have been used to reproduce COVID-19 incidence data and to make forecasts of new cases *(43–50)*. We decided to evaluate this approach using the fitting function described below, which is a discretized convolution of two integrals. This approach is related to an approach used to analyze acquired immunodeficiency syndrome (AIDS) data *(51, 52)*. Use of a curve-fitting model has the advantage of not requiring insights into disease transmission mechanisms, which may be particularly advantageous during outbreaks of new/emerging diseases. A drawback of a curve-fitting model is that it may be limited in its ability to reproduce empirical epidemic curves. We considered a fitting function that is able to generate an asymmetric curve (Appendix Figure 8), i.e., an epidemic curve with two timescales, as when there is fast growth and slow decay in new daily cases. Like many such models used in epidemiology, the curve-fitting model considered here can only generate single-peaked curves. Because the MSAs of interest have experienced multiple-wave disease-transmission dynamics (i.e., >1 period of increasing disease incidence), we are no longer using the curve-fitting model in forecasting. We present the curve-fitting model and results obtained with it to illustrate how a curve-fitting model can be combined with Bayesian inference to generate forecasts with uncertainty quantification.

For each metropolitan statistical area (MSA) of interest, we assume that there is an infection curve *Q*(*t*) describing the number of individuals who become infected at time *t* with SARS-CoV-2 and who will later be detected in local COVID-19 surveillance efforts. Furthermore, we assume that this curve has a shape that can be generated/reproduced by *ρ*_Γ_(*t, k, θ*), the probability density function (PDF) of a gamma distribution Γ(*k, θ*). In other words, we assume that *Q*(*t*) = *Nρ*_Γ_ (*t, k, θ*), where *N* is a scaling factor that we can identify as the number of individuals who will be detected over the entire course of the local epidemic. The shape of a gamma distribution is flexible and determined by the values of its two parameters: *k*, which is called the shape parameter, and *θ*, which is called the scale parameter. The functional form that we assume for *Q*(*t*) allows the curve-fitting model to reproduce the shape of an epidemic curve having two timescales. Early in the pandemic, many empirical COVID-19 epidemic curves appeared to have two timescales: an initial period during which new case reports increase relatively quickly from day to day followed by a period during which new case reports decrease relatively slowly from day to day.

We do not take the infection curve *Q*(*t*) to correspond directly to the number of new COVID-19 cases reported on the date encompassing time *t*, because only symptomatic individuals are likely to be detected in COVID-19 surveillance testing (to a first approximation). This situation complicates our model as there is known to be a potentially lengthy, variable delay in the onset of symptoms after infection *(12)*. We assume that the waiting time *τ* − *t* for the onset of COVID-19 symptoms after SARS-CoV-2 infection at time *t* is distributed according to a log-normal distribution. Let us use *ρ*_*LN*_(*τ* − *t*; *μ*_*LN*_, *σ*_*LN*_) to denote the PDF of the waiting-time distribution modeled by a log-normal distribution with parameters *μ*_*LN*_ and *σ*_*LN*_ set to the values estimated by Lauer et al. *(12)*. Let us use *I*(*t*_*i*_, *t*_*i*+1_) to denote the predicted number of new COVID-19 cases reported within a period beginning at time *t*_*i*_ ≡ *t*_0_+ *i d* and ending at time *t* _*i*+1_, where *t*_0_ > 0 is the start time of the local epidemic. We assume that surveillance testing for SARS-CoV-2 infection starts prior to time *t*_0_, and we take time *t* = 0 to correspond to 0000 hours on 21-January-2020, the date on which detection of the first US COVID-19 case was widely reported *(3)*. Under the aforementioned assumptions, *I*(*t*_*i*_,*t* _*i*+1_) is given by a convolution of integral functions. Namely, *I*(*t*_*i*_, *t*_*i*+1_) is given by the following expression:

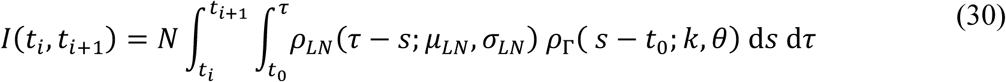

It should be noted that *s* in this expression is a dummy variable of integration. Equation (30) is a special case of the general model proposed by Brookmeyer and Gail *(51)* for predicting future AIDS cases.

Equation (30) can be evaluated through numerical quadrature, but this procedure is computationally expensive. To overcome this limitation, we replace the double integral in Equation (30) with a sum, and we calculate *I*(*t*_*i*_, *t*_*i*+1_) using the following expression instead of Equation (30):

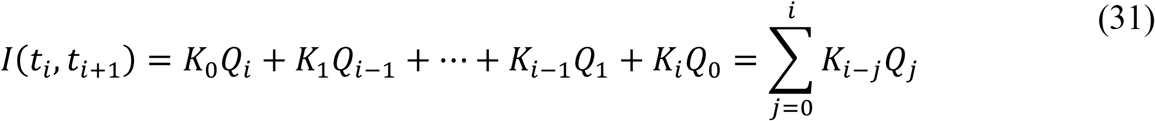

where

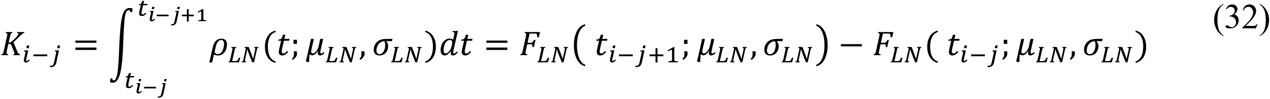

and

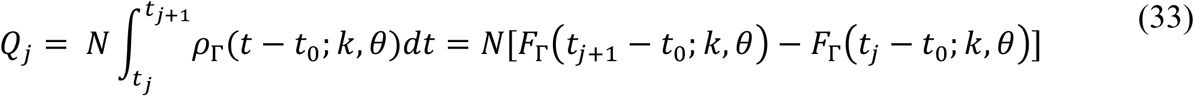

In Equation (31), the *K*_*i*−*j*_ terms are weighting functions (i.e., kernels) that account for the variable duration of the incubation period, and the *Q* _*j*_ terms represent cumulative numbers of new detectable infections occurring over discrete 1-d periods. In Equation (32), each *F*_*LN*_ term denotes a cumulative distribution function (CDF) of a log-normal distribution, and in Equation (33), each *F*_Γ_ term denotes a CDF of a gamma distribution. In other words, *Q*_*j*_ is the cumulative number of individuals infected in the period (*t*_*j*_, *t*_*j*+1_) who will eventually be detected, and *K*_*i*−*j*_ is the probability that one of these individuals becomes symptomatic and is detected in the period (*t*_*i*_, *t*_*i*+1_), where *t*_*i*_ ≥ *t*_*j*_.

The functional form of our curve-fitting model is defined by Equations (31)–(33), which are derived from Equation (30). As can be seen by inspecting Equation (30), the curve-fitting model has six parameters: *N, t*_0_ (which is hidden in the definition of *t*_*i*_), *k, θ, μ*_*LN*_, and *σ*_*LN*_. As noted earlier, estimates are available for *μ*_*LN*_ and *σ*_*LN*_ from Lauer et al. *(12)*. These parameters characterize the variable duration of the incubation period, which starts at infection and ends at the onset of symptoms. Thus, we take *μ*_*LN*_ and *σ*_*LN*_ to have fixed region-independent values. We take the remaining parameters—*N* (a population size/scaling factor), *t*_0_ (the start time of the local epidemic), and *k* and *θ* (the parameters that determine the shape of the infection curve *Q*(*t*))—to have adjustable region-specific values. In our daily inferences, we consider one additional region-specific adjustable parameter, the dispersal parameter of the likelihood function (see Equation (27) above). The value of this parameter, *r*, is inferred jointly with the values of *N, t*_0_, *k*, and *θ*.

For the period from January to June, 2020, for each of the 15 MSAs of interest, we parameterized on a daily basis a curve-fitting model for consistency with all daily reports of new confirmed cases available at the time. The methodology used was the same as that used for inferences conditioned on the compartmental model unless otherwise noted. For each MSA, the curve-fitting model was taken to have four adjustable parameters (Appendix Table 2): *N*, the total number of infected individuals who will be detected over the entire course of the local epidemic; *t*_0_, the start time of the local epidemic; and *k* and *θ*, the shape and scale parameters of a gamma (Γ) distribution. Inference of adjustable parameter values is based on a negative binomial likelihood function, which is given by Equation (27). The dispersal parameter *r* of the likelihood was taken to be adjustable; its value was jointly inferred with those of *N, t*_0_, *k*, and *θ*. Inferences are conditioned on fixed parameter estimates for *μ*_*LN*_ and *σ*_*LN*_ (Appendix Table 2). These parameters, the mean and standard deviation of a log-normal distribution, characterize the incubation period (i.e., the waiting time from infection to onset of symptoms) *(12)*. For any given (1-day) surveillance period and specified settings for parameter values, a prediction of the curve-fitting model for expected new cases detected was generated by evaluating the sum in Equation (31). A prediction of the actual number of new cases detected was obtained by entering the predicted expected number of new cases (according to either the curve-fitting or compartmental model) into Equation (29).

Predictive inferences conditioned on the curve-fitting model for all 15 MSAs of interest conditioned are shown in Appendix Figure 9. These results demonstrate that, for the timeframe of interest, the curve-fitting model was capable of reproducing many but not all of the MSA-specific empirical epidemic curves. The limitations of curve fitting can be seen by examining predictions for the Atlanta MSA, where there is high variability in the daily number of new cases detected. Although there is no clear downward trend in the data, the curve-fitting model nevertheless predicts a peak in late-April/early-May and a downward trend thereafter. This prediction is obtained because the model, by design, is only capable of generating single-peaked epidemic curves that rise and then fall.

## Footnotes

1 For the metropolitan statistical areas (MSAs) of interest, the number of political units (counties and independent cities) comprising an MSA ranges from 2 (for the Los Angeles and Riverside MSAs) to 29 (for the Atlanta MSA); the median (mean) number of counties is 7 (10). The number of States encompassing an MSA ranges from 1 (for eight of the 15 MSAs) to 4 (for Philadelphia); the median (mean) number of encompassing States is 1 (2).

2 Through mid-2020, we considered a curve-fitting model alongside the compartmental model, but we abandoned use of curve fitting after the MSAs of interest all experienced multiple-wave dynamics. The curve-fitting model is only able to generate single-peak epidemic curves. See the Appendix for details.

3 It should be noted that some related methods, typified by approximate Bayesian computation (ABC), do not rely on a likelihood.

4 This conclusion is supported by the Akaike information criterion (AIC) and Bayesian information criterion (BIC) values we calculated for the two scenarios (Appendix Table 1). Although AIC and BIC are crude model-selection tools because the posteriors here are non-normal, we deem them to be adequately discriminatory. Each strongly indicates that the model with two social-distancing periods is more explanatory of the data than the model with just one social-distancing period.

5 The 95% credible interval places the start date within the period beginning on 20-May-2020 and ending on 28-May-2020.

6 Unfortunately, our study sheds no light on which social-distancing practices are effective at slowing COVID-19 transmission.

## References

1. Gorbalenya AE, Baker SC, Baric RS, de Groot RJ, Drostetn C, Gulyaeva AA, et al.; Coronaviridae Study Group of the International Committee on Taxonomy of Viruses. The species Severe acute respiratory syndrome-related coronavirus: classifying 2019-nCoV and naming it SARS-CoV-2. Nat Microbiol. 2020;5:536–44.

2. Ghinai I, McPherson TD, Hunter JC, Kirking HL, Christiansen D, Joshi K, et al. Severe acute respiratory syndrome coronavirus 2 (SARS-CoV-2) in the USA. Lancet 2020;395:1137–44.

3. Holshue ML, DeBolt C, Lindquist S, Lofy KH, Wiesman J, Bruce H, et al. First case of 2019 novel coronavirus in the United States. N Engl J Med 2020;382:929–36.

4. The COVID Tracking Project at The Atlantic. https://covidtracking.com/data/us-daily

5. Silverman JD, Hupert N, Washburne AD. Using influenza surveillance networks to estimate state-specific prevalence of SARS-CoV-2 in the United States. Sci Transl Med 2020; 10.1126/scitranslmed.abc1126

6. Sanche S, Lin YT, Xu C, Romero-Severson E, Hengartner N, Ke R. High contagiousness and rapid spread of severe acute respiratory syndrome coronavirus 2. Emerg Inf Dis 2020;26:1470–7.

7. Coronavirus Resource Center, Johns Hopkins University. Timeline of COVID-19 policies, cases, and deaths in your state: a look at how social distancing measures may have influenced trends in COVID-19 cases and deaths. https://coronavirus.jhu.edu/data/state-timeline

8. Courtemanche C, Garuccio J, Le A, Pinkston J, and Yelowitz A. Strong social distancing measures in the United States reduced the COVID-19 growth rate. Health Aff 2020; https://doi.org/10.1377/hlthaff.2020.00608

9. Hsing S, Allen D, Annan-Phan S, Bell K, Bolliger I, Chong T, et al. The effect of large- scale anti-contagion policies on the COVID-19 pandemic. Nature 2020; https://doi.org/10.1038/s41586-020-2404-8

10. Vought RT. Revised delineations of metropolitan statistical areas, micropolitan statistical areas, and combined statistical areas, and guidance on uses of the delineations of these areas. US Office of Management and Budget Bulletin 2020;20–01

11. The New York Times. Coronavirus (Covid-19) data in the United States. https://github.com/nytimes/covid-19-data

12. Lauer SA, Grantz KH, Bi Q, Jones FK, Zheng Q, Meredith HR, et al. The incubation period of coronavirus disease 2019 (COVID-19) from publicly reported confirmed cases: estimation and application. Ann Intern Med 2020;172:577–82. https://doi.org/10.7326/M20-0504

13. United States Census Bureau. Metropolitan and micropolitan statistical areas population totals and components of change: 2010–2019. https://www.census.gov/data/tables/time-series/demo/popest/2010s-total-metro-and-micro-statistical-areas.html

14. Arons MM, Hatfield KM, Reddy SC, Kimball A, James A, Jacobs JR, et al. Presymptomatic SARS-CoV-2 infections and transmission in a skilled nursing facility. N Engl J Med 2020;382:2081–90.

15. Nguyen VVC, Vo TL, Nguyen TD, Lam MY, Ngo NQM, L.MH, et al. The natural history and transmission potential of asymptomatic SARS-CoV-2 infection. Clin Infect Dis 2020;ciaa711 https://doi.org/10.1093/cid/ciaa711

16. Ministry of Health, Labour and Welfare of Japan. Official report on the cruise ship Diamond Princess, May 1, 2020. https://www.mhlw.go.jp/stf/newpage_11146.html

17. Sakurai A, Sasaki T, Kato S, Hayashi M, Tsuzuki SI, Ishihara T, et al. Natural history of asymptomatic SARS-CoV-2 infection. N Engl J Med 2020; https://doi.org/10.1056/NEJMc2013020

18. Perez-Saez J, Lauer SA, Kaiser L, Regard S, Delaporte E, Guessous I, et al. Serology- informed estimates of SARS-COV-2 infection fatality risk in Geneva, Switzerland. 2020; https://osf.io/wdbpe/

19. Richardson S, Hirsch JS, Narasimhan M, Crawford JM, McGinn T, Davidson KW, et al. Presenting characteristics, comorbidities, and outcomes among 5700 patients hospitalized with COVID-19 in the New York City area. JAMA 2020;323:2052–9.

20. Wölfel R, Corman VM, Guggemos W, Seilmaier M, Zange S, Müller MA, et al. Virological assessment of hospitalized patients with COVID-2019. Nature 2020;581:465–9.

21. Andrieu C, Thoms J. A tutorial on adaptive MCMC. Stat Comput 2008;18:343–73.

22. Hindmarsh AC. ODEPACK, a systematized collection of ODE solvers. In Scientific Computing (Stepleman RS, Ed) 1983;pp 55–64. North-Holland, Amsterdam.

23. U.S. Department of Energy. COVID-19 pandemic modeling and analysis (https://covid19.ornl.gov) and Los Alamos COVID-19 City Predictions (https://github.com/lanl/COVID-19-Predictions).

## Appendix References

24. . Anderson SC, Edwards AM, Yerlanov M, Mulberry N, Stockdale J, Iyaniwura SA, Falcao RC, Otterstatter MC, Irvine MA, Janjua NZ, Coombs D, and Colijn C. Estimating the impact ofCOVID-19 control measures using a Bayesian model of physical distancing 2020. https://www.medrxiv.org/content/10.1101/2020.04.17.20070086v1

25. Sia SF, Yan LM, Chin AWH, Fung K, Choy KT, et al. Pathogenesis and transmission of SARS-CoV-2 in golden hamsters. Nature. 2020;583:834–838.

26. Kucirka LM, Lauer SA, Laeyendecker O, Boon D, Lessler J. Variation in false-negative rate of reverse transcriptase polymerase chain reaction-based SARS-CoV-2 tests by time since exposure. Ann Intern Med 2020;173:262–7.

27. Sekine T, Perez-Potti A, Rivera-Ballesteros O, Strålin K, Gorin JB, et al. Robust T cell immunity in convalescent individuals with asymptomatic or mild COVID-19. Cell. 2020; https://doi.org/10.1016/j.cell.2020.08.017

28. Addetia A, Crawford KHD, Dingens A, Zhu H, Roychoudhury P, et al. Neutralizing antibodies correlate with protection from SARS-CoV-2 in humans during a fishery vessel outbreak with high attack rate. J Clin Microbiol 2020; https://doi.org/10.1128/JCM.02107-20

29. To KKW, Hung IFN, Ip JD, Chu AWH, Chan WM, et al. COVID-19 re-infection by a phylogenetically distinct SARS-coronavirus-2 strain confirmed by whole genome sequencing. Clin Infect Dis 2020; https://doi.org/10.1093/cid/ciaa1275

30. Papst I, Li M, Champredon D, Bolker BM, Dushoff J, Earn DJ. Age-dependence of healthcare interventions for SARS-CoV-2 infection in Ontario, Canada. 2020; https://www.medrxiv.org/content/10.1101/2020.09.01.20186395v2

31. Elderd BD, Dukic VM, Dwyer G. Uncertainty in predictions of disease spread and public health responses to bioterrorism and emerging diseases. Proc Natl Acad USA 2006;103:15693–7.

32. He X, Lau EHY, Wu P, Deng X, Wang J, Hao X, et al. Temporal dynamics in viral shedding and transmissibility of COVID-19. Nat Med 2020;26:672–5.

33. Benefield AE, Skrip LA, Clement A, Althouse RA, Chang S, Althouse BM. SARS-CoV- 2 viral load peaks prior to symptom onset: a systematic review and individual-pooled analysis of coronavirus viral load from 66 studies. 2020; https://doi.org/10.1101/2020.09.28.20202028

34. Böhmer MM, Buchholz U, Corman VM, Hoch M, Katz K, Marosevic DV, et al. Investigation of a COVID-19 outbreak in Germany resulting from a single travel-associated primary case: a case series. The Lancet 2020; https://doi.org/10.1016/S1473-3099(20)30314-5

35. Emery JC, Russell TW, Liu Y, Hellewell J, Pearson CAB, CMMID COVID-19 Working Group, Knight GM, Eggo RM, Kucharski AJ, Funk S, Flasche S, Houben RMGJ. The contribution of asymptomatic SARS-CoV-2 infections to transmission on the Diamond Princess cruise ship. eLife 2020;9:e58699.

36. Expert Taskforce for the COVID-19 Cruise Ship Outbreak. Epidemiology of the COVID- 19 outbreak on cruise ship quarantined at Yokohama, Japan, February 2020. Emerg Inf Dis 2020;26:2591–7.

37. Lavezzo E, Franchin E, Ciavarella C, Cuomo-Dannenburg G, Barzon L, Del Vecchio C, et al. Suppression of COVID-19 outbreak in the municipality of Vo, Italy. 2020; https://www.medrxiv.org/content/10.1101/2020.04.17.20053157v1

38. Gudbjartsson DF, Helgason A, Jonsson H, Magnusson OT, Melsted P, Norddahl GL, et al. Spread of SARS-CoV-2 in the Icelandic population. N Engl J Med 2020;282:2302–15.

39. Grewelle RE, De Leo GA. Estimating the global infection fatality rate of COVID-19. 2020; https://www.medrxiv.org/content/10.1101/2020.05.11.20098780v1.full.pdf

40. Burnham KP and Anderson DR. Multimodal inference: understanding AIC and BIC in model selection. Sociological Methods Research 2004;33:261–304.

41. Lin YT, Kim H, Doering CR. Demographic stochasticity and evolution of dispersion. I. Spatially homogeneous environments. J Math Biol 2015;70:647–78.

42. Lin YT, Feng S, Hlavacek WS. Scaling methods for accelerating kinetic Monte Carlo simulations of chemical reaction networks. J Chem Phys 2019;150:244101.

43. Chen DG, Chen X, Chen JK. Reconstructing and forecasting the COVID-19 epidemic in the United States using a 5-parameter logistic growth model. Glob Health Res Policy. 2020;5:25.

44. IHME COVID-19 Health Service Utilization Forecasting Team, Murray CJL. Forecasting COVID-19 impact on hospital bed-days, ICU-days, ventilator-days and deaths by US State in the next 4 months. 2020;medRxiv preprint: https://doi.org/10.1101/2020.03.27.20043752

45. Moreau VH. Forecast predictions for the COVID-19 pandemic in Brazil by statistical modeling using the Weibull distribution for daily new cases and deaths. Braz J Microbiol. 2020;51:1109-15.

46. . Nishimoto Y, Inoue K. Curve-fitting approach for COVID-19 data and its physical background. 2020;medRxiv preprint: https://doi.org/10.1101/2020.07.02.20144899

47. Roosa K, Lee Y, Luo R, Kirpich A, Rothenberg R, et al.; Short-term forecasts of the COVID-19 epidemic in Guangdong and Zhejiang, China: February 13-23, 2020. J Clin Med. 2020;9:596.

48. Rypdal K, Rypdal M. A parsimonious description and cross-country analysis of COVID- 19 epidemic curves. Int J Environ Res Public Health. 2020;17:E6487.

49. Wu K, Darcet D, Wang Q, Sornette D. Generalized logistic growth modeling of the COVID-19 outbreak: comparing the dynamics in the 29 provinces in China and the rest of the world. Nonlinear Dyn. 2020;https://doi.org/10.1007/s11071-020-05862-6

50. Xu J, Cheng Y, Yuan X, Li WV, Zhang L. Trends and prediction in daily incidence of novel coronavirus infection in China, Hubei Province and Wuhan City: an application of Farr’s law. Am J Transl Res. 2020;12:1355–61.

51. Brookmeyer R, Gail MH. Minimum size of the acquired immunodeficiency syndrome (AIDS) epidemic in the United States. Lancet. 1986;2:1320–2.

52. Brookmeyer R. Reconstruction and future trends of the AIDS epidemic in the United States. Science. 1991;253:37–42.

